# Modeling the genomic architecture of adiposity and anthropometrics across the lifespan

**DOI:** 10.1101/2024.08.14.24312003

**Authors:** Christopher H. Arehart, Meng Lin, Raine A. Gibson, Sridharan Raghavan, Christopher R. Gignoux, Maggie A. Stanislawski, Andrew D. Grotzinger, Luke M. Evans

## Abstract

Obesity-related conditions are among the leading causes of preventable death and are increasing in prevalence worldwide. Body size and composition are complex traits that are challenging to characterize due to environmental and genetic influences, longitudinal variation, heterogeneity between sexes, and differing health risks based on adipose distribution. We constructed a 4-factor genomic structural equation model using 18 measures and unveiled shared and distinct genetic architectures underlying birth size, abdominal size, adipose distribution, and adiposity. Multivariate genome-wide associations revealed the adiposity factor was enriched specifically in neural tissues and pathways, while adipose distribution was enriched across widespread physiological systems. In addition, polygenic scores for the adiposity factor predicted many adverse health outcomes, while body size and composition predicted a more limited subset. Finally, we characterized the factors’ genetic correlations with obesity-related traits and examined the druggable genome through constructing a bipartite drug-gene network to identify viable therapeutic targets.

## Introduction

Human body size and body composition vary throughout an individual’s lifecourse and across individuals in a population. The strong associations linking excess fat stores with a constellation of morbidities have highlighted the importance of understanding how various anthropometric traits are connected to the broad and multifaceted biological systems underpinning human health. Obesity prevalence has increased markedly in the United States between 1999 and 2020 from 30.5% to 41.9%.^1^ On a global scale, the increasing rates of obesity observed among children and adults are a widespread source of concern;^2^ obesity-related conditions such as heart disease, stroke, type 2 diabetes (T2D), and some cancers are among the leading causes for preventable death.^3^ Although family-based^2^ and genome-wide association studies (GWASs)^4^ point to substantive genetic influences on obesity, the broader landscape of what characterizes this genetic signal across different measures of adiposity remains poorly understood.

The phenotypic and genetic signal of adiposity traits is remarkably difficult to characterize due to heterogeneity between sexes and longitudinal variation across the lifespan. The genetic architecture for adipose distribution is notably different between males and females,^5^ and women exhibit a greater ratio of subcutaneous-to-visceral adipose tissue than men.^6^ Moreover, the amount of visceral adipose tissue tends to increase with age for both males and females, but men tend to lose relatively more visceral adipose tissue due to calorie restriction than women.^6–8^ Body mass index (BMI) – an easily obtainable clinical measure (diagnosing obesity as BMI ≥ 30 kg/m^2^) – falls short when differentiating between masses of visceral adipose, subcutaneous adipose, muscle, or bone, leading to its criticism as a misleading metric of body composition and cardiometabolic health.^9–11^ Waist circumference adjusted for BMI (WCadjBMI), hip circumference adjusted for BMI (HCadjBMI), and waist-to-hip circumference ratio adjusted for BMI (WHRadjBMI)^5^ are proxy measures of body fat distribution. Notably, the genetic drivers of BMI and WHRadjBMI are distinct: genetic associations for BMI and obesity are linked to enriched gene expression in the central nervous system (CNS), implicating a relationship between obesity and the brain,^2,5,12,13^ whereas genes associated with WHRadjBMI demonstrate less enrichment for tissue-specific expression in the CNS and more with gene expression in preadipocytes and adipocytes.^5,14^ Similarly, the genetic contributors to metabolic syndrome (MetSyn) – a cluster of often comorbid risk factors (e.g., hypertension) that link adiposity with cardiovascular disease and T2D – strongly overlap with the genetic associations for waist circumference (WC).^15^ However, the alleles associated with a higher subcutaneous-to-visceral adipose distribution (increased capacity for adipose tissue expansion)^16^ are protective for T2D, heart disease, and high blood pressure.^17,18^ These findings highlight the complexity of body composition and genetic influences, with sometimes contrasting effects on health outcomes.

Given this complex and intertwined landscape of anthropometric measurements, we speculated that the genetic associations for human body size and body composition would be more suitably represented as latent variables in a genomic structural equation modeling (Genomic SEM) framework.^19^ Genomic SEM estimates how strongly the genetic associations of various observed traits are related to a number of underlying and unobserved genetic constructs (latent factors). It does so by estimating the strengths of the relationships (loadings) of each trait with the factors, which themselves can be related to one another (genetic correlations). A primary characteristic of Genomic SEM is its ability to include different sets of traits from various participant samples; this enabled us to incorporate a diverse range of anthropometric traits from across the lifespan and stratified by biological sex into the same statistical model. Through this modeling process, we balanced model complexity and parsimony to unveil the shared and distinct genetic components underlying differences in birth size, abdominal size, weight distribution, and adiposity. We found the enrichment of biological pathways and tissue types to be distinct among the 4 genetic factors in the model, and the factors showed different associations with adverse health outcomes in an independent dataset with electronic health records. In addition, we contextualized the genome-wide signal for each of the factors by identifying differing genetic correlations with obesity-related traits. Together, our results particularly highlighted the adiposity factor for its distinct enrichment in nervous systems, genetic correlations with related traits, and prediction of adverse health outcomes across broad phenotypic domains. Finally, we examined the druggable genome and constructed a bipartite drug-gene network to identify possible mechanistic explanations for weight-related side effects and the potential for repurposing therapeutics to address adiposity.

## Results

### A four-factor model of anthropometric and adiposity genetics

We began by bringing together GWAS summary statistics for 18 adiposity and anthropometric measures from different points in the lifespan and stratified by sex (Supplementary Table 1). The Genomic SEM model in Fig. 1 revealed an overall structure with 4 latent genetic factors referred to as F1-F4 and had adequate model fit^20,21^ (comparative fit index [CFI] = 0.94 and a standardized root mean square residual [SRMR] = 0.11). The genetic covariance and correlation matrices are shown in Supplementary Figs. 1-2 along with further description of the modeling techniques and considerations in the online methods. The model estimated differing strengths of relationships between the 18 genetic indicator variables and their underlying latent constructs, as represented by their factor loadings (Fig. 1 one-directional arrows). F1 included 3 loadings for traits related to birth size, F2 included 3 loadings for traits relating to abdominal size, F3 included 7 loadings for traits relating to body size and adipose distribution, and F4 included 7 loadings for traits relating to adiposity. The 4 factors generally exhibited small genetic correlations (Fig. 1; r_g_ ≤ 0.15 indicated by two-directional arrows representing standardized covariance relationships between the factors). The only sizable genetic correlation was for F1 and F3 (r_g_ = 0.44), likely reflecting the shared genetic effects of birth length and adult height (r_g_ = 0.49). Together, this emphasized the unique subclusters of genetic signal across traits that are often thought of as similar proxy measurements of anthropometry and adiposity.

**Figure 1:**
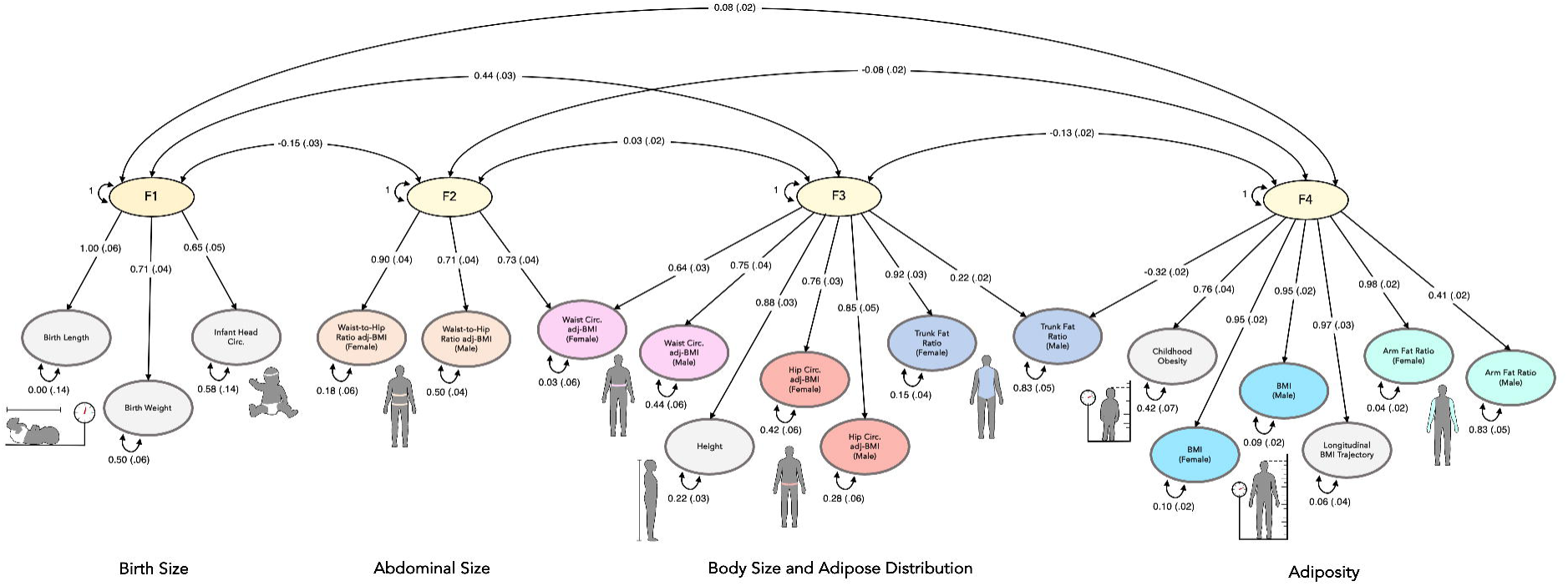
Genomic structural equation model of adiposity and anthropometrics across the lifespan. The standardized measurement model derived using genomic structural equation modeling (SEM) comprised of 4 latent genetic factors and 18 indicator variables. The 4 genetic factors are shaded yellow, the traits with combined males and females are shaded gray, and the traits stratified by males and females are shaded in color-matched pairs. The one-directional arrows signify standardized factor loadings and describe the strength and direction of the relationships between genetic indicators and their underlying latent constructs. Standardized covariance relationships (i.e., correlations) between the factors are represented by two-directional arrows, and the two-directional arrows pointing from a variable to itself to denote the standardized residuals (the unique genetic variance not reflected through other paths in the model).

Among the 6 sex-stratified traits, each male-female pair generally loaded onto the same factor, highlighting the largely shared genetic associations within males and females. HCadjBMI male and female had similar loadings on F3, and BMI male and female had similar loadings on F4 – however, across the other sex-stratified traits there were some notable differences. More genetic variance of WHRadjBMI was explained by F2 in females relative to males (see loadings in Fig. 1), and F4 explained more variance of female than male arm fat ratio (AFR). In addition, the variance in female trunk fat ratio (TFR) was mostly explained by F3, but male TFR had modest cross-loadings between F3 and F4, with substantial residual genetic variance (0.83) and generally low genetic covariance (Supplementary Fig. 1) with other anthropometric traits, suggesting a more divergent genetic influence on male TFR. WCadjBMI female cross-loaded substantially onto both F2 and F3, while WCadjBMI male only loaded on F3. One primary advantage of our SEM model is its ability to estimate these sex-specific differences and relationships within the landscape of anthropometric traits across the lifespan. The 4 factors in our model provide latent constructs that are less prone to measurement error and can discern the genetic components relating to body size and body composition; as such, this novel genetic representation goes beyond any single indicator variable, such as BMI.

We subsequently used our 4-factor Genomic SEM to perform multivariate GWASs, which leveraged improved power over the constituent indicator GWASs. We identified multiple genome-wide significant (GWS; p < 5 x 10^-8^) variants that were unique to each factor and not identified in the underlying GWASs after removing SNPs with heterogeneous effects (Q_SNPs_; Supplementary Table 2). F1, F2, F3, and F4 respectively uncovered 103; 1,318; 8; and 6,206 novel GWS SNPs that were not identified in the indicator GWASs for each factor, and Manhattan plots for each multivariate factor-GWAS are shown in Supplementary Figs. 3-6. We characterized these multivariate GWASs in multiple downstream analyses. First, we implemented DEPICT^22^ to identify significantly prioritized genes (FDR < 0.05) from the 88; 344; 1,173; and 675 independent GWS loci for F1, F2, F3, and F4 respectively, and assessed the enrichment of those loci across functional gene sets (p < 4.56×10^-6^, the Bonferroni-corrected significance threshold) and tissue-specific expression profiles (FDR < 0.05). Next, we used FOCUS^23^ to perform transcription-level analyses for each of the latent factor GWASs, and we extracted genes with posterior inclusion probability (PIP) > 0.1 that were fine-mapped to non-null 90% credible sets (CSs). We additionally used the factor multivariate GWASs’ effect estimates and LDpred2^24^ to calculate 4 polygenic risk scores (PRSs) in an external dataset (N = 25,240) which were then tested for association (FDR < 0.10, due to the highly correlated structure of the phecodes) with 1,591 phecode-based phenotypes in a phenome-wide association study (pheWAS). Next, we estimated the genetic correlations with comorbidity-related traits to contextualize each factor within a broader genomic landscape using LDSC.^25,26^ Finally, we constructed drug-gene interaction networks for the factors’ DEPICT- and FOCUS-identified genes to advance existing, proposed, and novel therapeutic targets for adiposity-related conditions.

### F1 – birth size

F1 characterized the genetic signal underlying size at birth with loadings from 3 indicator variables (Fig. 2a). The DEPICT analysis highlighted 88 independent GWS loci with 24 significantly prioritized genes and 3 enriched gene sets including ‘incomplete somite formation’ and ‘decreased embryo size’ gene sets. The GWS loci for F1 were not enriched for expression profiles across physiological systems, cell types, or tissue types (Fig. 2b). In a tissue-agnostic transcriptome wide association study (TWAS) analysis using FOCUS, however, there were 158 finemapped genes across 69 CSs (Supplementary Fig. 11). These putatively causal gene-expression mediated effects consisted of SNP-expression weights from 27 general tissues including the brain (43 genes), adipose (17 genes), and esophagus (16 genes). The F1 PRSs that were validated in an external dataset (N = 25,240) were negatively associated with acute sinusitis, insomnia, renal failure, T2D, and hypertension (Fig. 2c).

**Figure 2:**
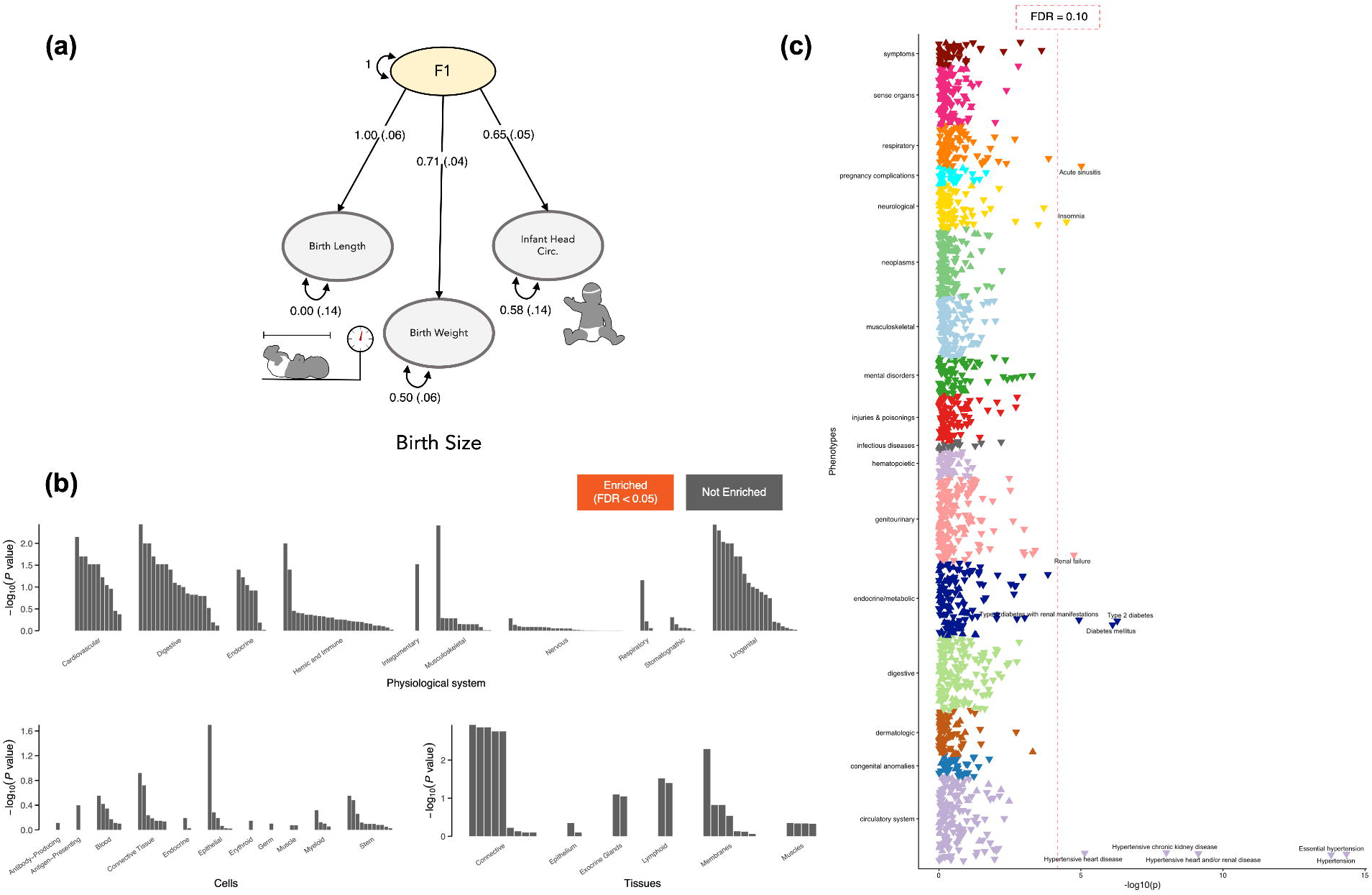
Characterizing F1 – the genetics of birth size. The 3 indicator variables relating to birth size and their standardized loadings on F1, the 1^st^ latent genetic factor, are shown in (a). This genetic factor did not have any genetic enrichment across physiological systems, cell types, or tissue types (FDR < 0.05) in the DEPICT analysis (b). The polygenic risk score (PRS) weights for F1 were validated in an external sample (CCPM Biobank, N = 25,240) and implemented in a phenome wide association study (pheWAS); The significant pheWAS associations between F1 PRS and phenotypes are shown in (c), with phenotype labels for the points to the right of the vertical dashed red line denoting FDR < 0.10, and triangle direction (up/down) indicating F1 PRS direction of effect (+/-).

### F2 – abdominal size

F2 had 3 loadings from indicator variables relating to adult abdominal size (Fig. 3a) and 319 significant DEPICT-prioritized genes from 344 independent GWS loci. We observed significant physiological system enrichment across 7 of the 10 categories (Fig. 3b), including adipocytes, subcutaneous adipose tissue, and abdominal adipose tissue. Beyond those adipose-related tissues, F2’s genetic signal was broadly enriched throughout the body including the musculoskeletal, urogenital, cardiovascular, digestive, and endocrine systems. Using tissue-agnostic FOCUS TWAS we identified 676 finemapped genes across 243 CSs (Supplementary Fig. 12). These prioritized TWAS associations spanned 28 general tissues but primarily consisted of brain (160 genes) and adipose tissue weights (78 genes). F2 PRS-pheWAS showed positive associations with T2D, peripheral angiopathy, and hypertension (Fig. 3c) suggesting a genetic propensity for larger abdominal size was predictive of these circulatory and metabolic health outcomes. These phenotypic associations were aligned with the DEPICT gene-set analysis which identified 185 significantly enriched gene sets relating to insulin resistance and organ development/morphology (particularly within the cardiovascular system).

**Figure 3:**
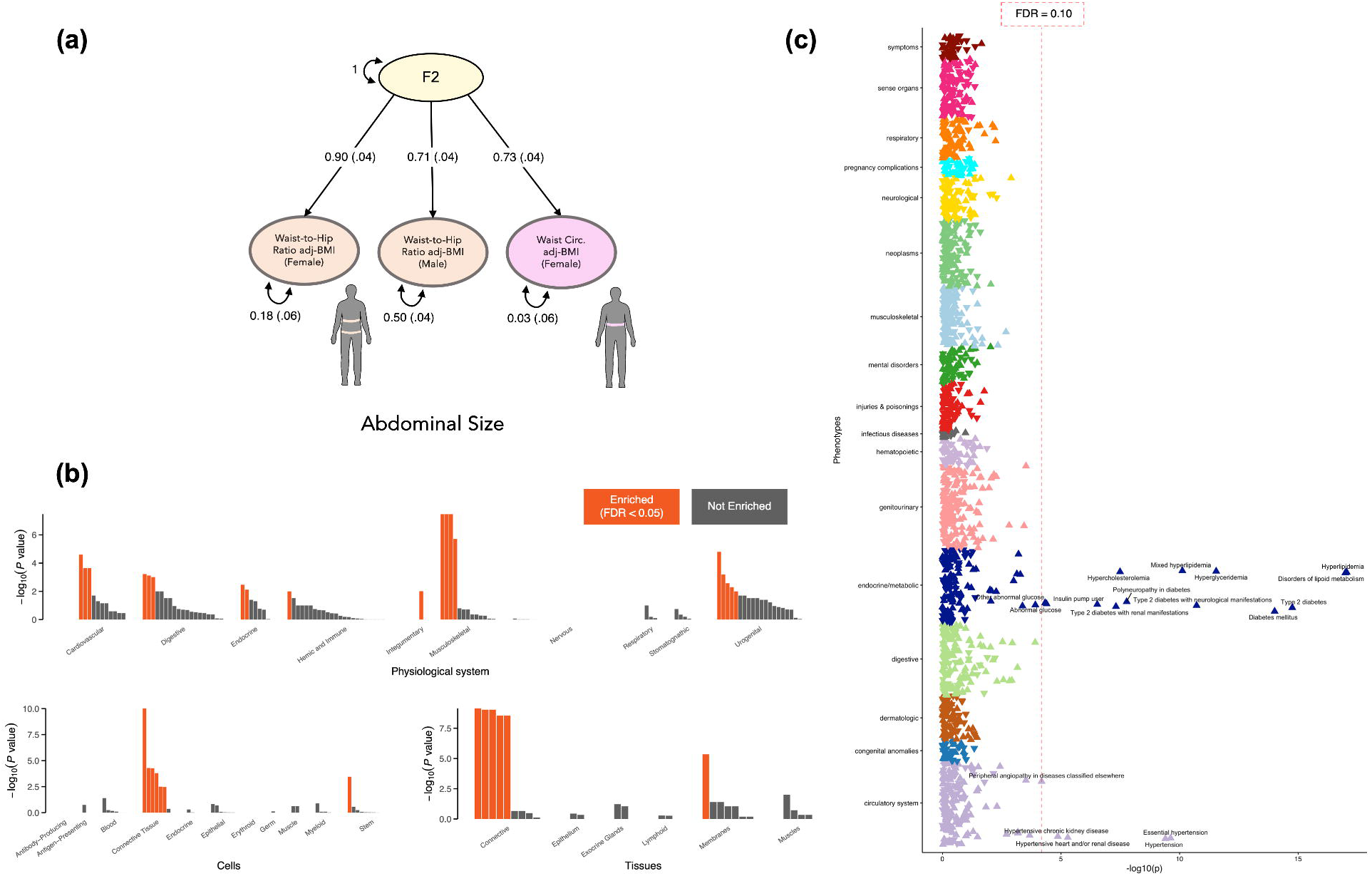
Characterizing F2 – the genetics of abdominal size. The 3 indicator variables relating to abdominal size and their standardized loadings on F2, the 2^nd^ latent genetic factor, are shown in (a). This genetic factor showed gene expression enrichment across a variety of physiological systems, cell types, and tissue types (orange coloring, FDR < 0.05) in the DEPICT analysis (b). The polygenic risk score (PRS) weights for F2 were validated in an external sample (CCPM Biobank, N = 25,240) and implemented in a phenome wide association study (pheWAS); The significant pheWAS associations between F2 PRS and phenotypes are shown in (c), with phenotype labels for the points to the right of the vertical dashed red line denoting FDR < 0.10, and triangle direction (up/down) indicating F2 PRS direction of effect (+/-).

### F3 – body size and adipose distribution

The third genetic factor, F3, captured the shared variance among 7 indicator variables describing body size and adipose distribution (Fig. 4a), with notable differences between the loadings for male and female traits, especially for TFR (described above). The DEPICT analysis for F3 identified 1,864 significantly prioritized genes for 1,173 independent GWS loci and enrichment in 8 of the 10 physiological system categories (Fig. 4b; musculoskeletal, urogenital, cardiovascular, endocrine, digestive, respiratory, hemic and immune, integumentary), exemplifying the multifaceted physiology underlying variation in adult body size and adipose distribution. We found 1,127 gene sets significantly enriched for F3, including many gene sets relating to embryonic development and protein-protein interaction subnetworks. In a tissue-agnostic FOCUS TWAS, we identified 2,266 finemapped genes across 689 CSs (Supplementary Fig. 13), spanning 28 general tissues, particularly brain (571 genes), esophagus (242 genes), adipose (218 genes), and artery (202 genes). Interestingly, *APOE*, a gene linked to Alzheimer’s disease and catabolism of lipoprotein constituents, was significantly associated via prostate expression weights (Z-score = -5.35, PIP = 0.61). The PRS-pheWAS analysis revealed that F3 was predictive of a few health outcomes including negative associations with abdominal pain, hyperlipidemia, and hypertension, but a positive association with atrial fibrillation.

**Figure 4:**
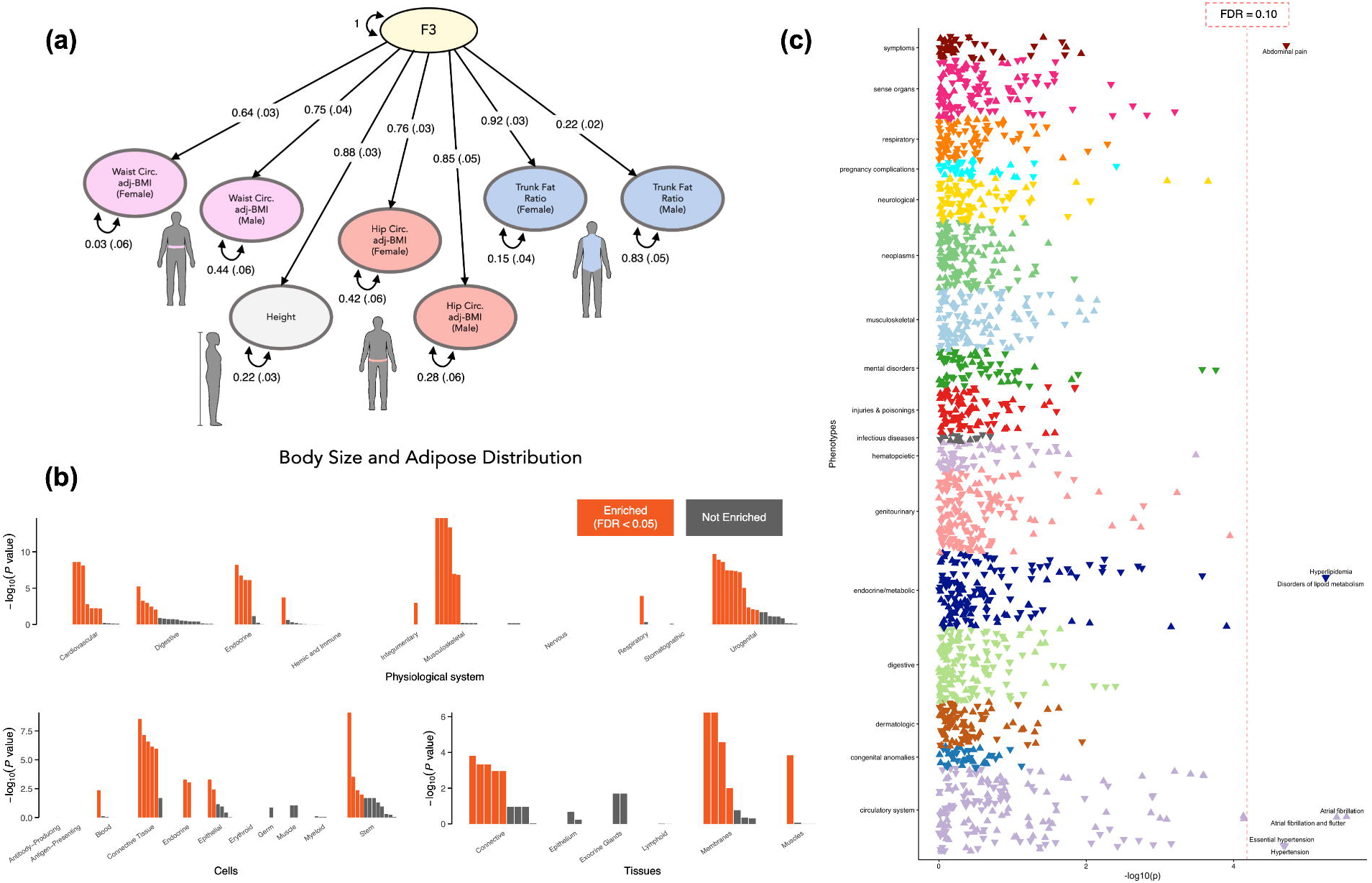
Characterizing F3 – the genetics of body size and adipose distribution. The 7 indicator variables relating to body size and adipose distribution and their standardized loadings on F3, the 3^rd^ latent genetic factor, are shown in (a). This genetic factor showed gene expression enrichment across a variety of physiological systems, cell types, and tissue types (FDR < 0.05) in the DEPICT analysis (b). The polygenic risk score (PRS) weights for F3 were validated in an external sample (CCPM Biobank, N = 25,240) and implemented in a phenome wide association study (pheWAS); The significant pheWAS associations between F3 PRS and phenotypes are shown in (c), with phenotype labels for the points to the right of the vertical dashed red line denoting FDR < 0.10, and triangle direction (up/down) indicating F3 PRS direction of effect (+/-).

### F4 – adiposity

F4 had 7 adiposity-related indicator variables loading onto it relating to excess fat tissue and obesity (Fig. 5a). The associated loci were enriched only in one physiological system (nervous; Fig. 5b). Broad regions across the CNS were enriched, including the hindbrain (cerebellum) and the forebrain (cerebral cortex, temporal lobe, occipital lobe, frontal lobe, parietal lobe, basal ganglia) – regions responsible for complex perceptual, cognitive, and behavioral processes involving learning, emotion, and memory. The F4 DEPICT analysis identified 437 significantly prioritized genes for the 675 independent GWS loci and 62 enriched gene sets; upon comparing these gene sets to the other 3 factors, they were much more specific to the CNS, relating to brain development, neurons, synaptosomes, and dendrites. In a brain-tissue-prioritized FOCUS TWAS, we identified 850 finemapped genes across 335 CSs (Supplementary Fig. 14). These prioritized TWAS associations spanned 28 general tissues but the majority corresponded to brain tissue weights (498 genes). The PRS-pheWAS analysis for F4 uncovered many more associations with adverse health outcomes, spanning a wide range of domains (Fig. 5c): chronic pain, fatigue, asthma, shortness of breath, sleep apnea, benign skin neoplasm, cancer of kidney and renal pelvis, osteoarthritis, substance use disorders, anxiety, depression, sepsis, allergy to medications, skin/nail fungal infections, anemia, renal disease/failure, obesity, T2D, liver disease/cirrhosis, bariatric surgery, esophageal diseases, acid reflux, cellulitis, long-term anticoagulants, and hypertension.

**Figure 5:**
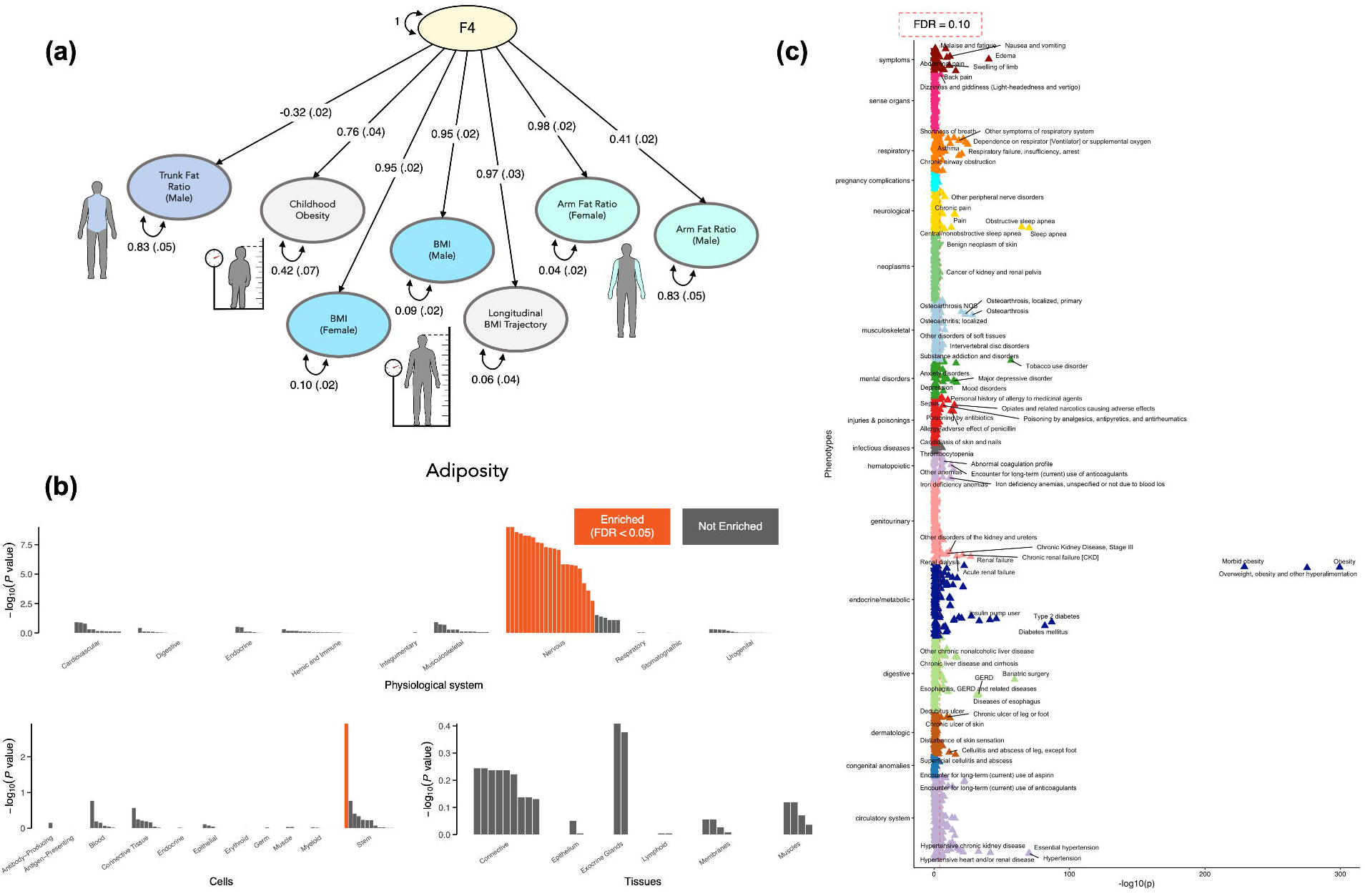
Characterizing F4 – the genetics of adiposity. The 7 indicator variables relating to adiposity and their standardized loadings on F4, the 4^th^ latent genetic factor, are shown in (a). This genetic factor showed gene expression enrichment only in nervous physiological systems and cell types (FDR < 0.05) in the DEPICT analysis (b). The polygenic risk score (PRS) weights for F4 were validated in an external sample (CCPM Biobank, N = 25,240) and implemented in a phenome wide association study (pheWAS); The significant pheWAS associations between F4 PRS and phenotypes are shown in (c), with phenotype labels for the points to the right of the vertical dashed red line denoting FDR < 0.10, and triangle direction (up/down) indicating F4 PRS direction of effect (+/-).

### Comparison of F4 and BMI genetic signals

The male and female BMI indicator variables both had large standardized loadings of 0.95 with F4; therefore, we explored the shared versus novel aspects of the genetic signals for F4 (a highly predictive latent factor) compared to BMI. There were 6,578 GWS SNPs common between the F4 and BMI GWASs, but 6,206 SNPs that were novel to F4 (i.e., not GWS in any of the indicator GWASs loading onto F4, including BMI). Overall, the GWS SNPs for F4 and BMI (combined males and females) resided in 675 and 1,035 independent significant loci, respectively, which only partially overlapped (624 of the 675 F4 loci had genomic positional overlap with the BMI loci [Extended Data Fig. 1, Supplementary Figs. 6-7]). Notably, while 392 DEPICT-prioritized genes were common to BMI and F4, 45 genes were unique to only F4 (Supplementary Table 18, Extended Data Fig. 2). In addition, while 339 putatively causal genes with expression mediated effects (FOCUS-identified genes) were common to BMI and F4, 511 genes were unique to F4. Only 21 genes were common to all 4 analyses (identified by DEPICT and FOCUS for both F4 and BMI). Beyond these distinguishing overlaps at the gene-level, the DEPICT tissue enrichment analyses (Supplementary Fig. 9) pinpointed a key difference between F4 and BMI: the BMI-associated genetic loci were distinctively enriched for the hypothalamus and the hypothalamo hypophyseal system – the brain’s control center for hunger and satiety. The BMI-associated loci were therefore enriched in the canonical energy homeostasis-related areas of the brain whereas the F4-associated loci were not. Thus, F4 was characterized by a novel partitioning of the genetic architecture of adiposity; F4 disentangles a neural and behavioral component of adiposty that is rooted in sensory processing, learning, memory, and experience.

The genetic differences between F4 and BMI motivated us to perform an additional pheWAS controlling for BMI to investigate the conditionally idependent associations of F4’s polygenic risk with heath outcomes (Extended Data Fig. 3). We observed an attenuation of the F4-pheWAS associations, as expected, after conditioning on BMI (Supplementary Fig. 10); several health outcomes including chronic pain, sleep apnea, depression, and acid reflux dropped below the significance threshold, implicating BMI as a potential mediator for some disease associations. However, F4 clearly captured additional and unique contributions to health outcomes beyond BMI alone, with F4 still positively and significantly predicting adverse health outcomes for the vast majority of associations (Fig. 5c, Extended Data Fig. 3). These results illustrate the utility of F4 as a polygenic predictor beyond BMI, and they showcase the added value of our model for disentangling the genetics of adiposity and anthropometrics across the lifespan.

### Genetic correlations with related traits

Following the characterization of each of the 4 factors with regard to their genome-, transcriptome-, and phenome-wide associations, we estimated LDSC-based genetic correlations between each factor and 75 adiposity-related traits (Supplementary Tables 3 and 48), including metabolism, substance use, psychopathology, neuroticism, risk tolerance, diet, sleep, exercise, pain, frailty, dementia, inflammatory disease, autoimmune disease, and cardiovascular disease. The full genetic covariance and correlation matrices are shown in Supplementary Figs. 16-17 and Supplementary Tables 49-52, and pairwise genetic correlations with F1, F2, F3, and F4 are shown in Extended Data Fig. 4 and Supplementary Figs. 18-26. Fig. 6 depicts the prominent genetic correlations (> 0.15) with each of the factors in our Genomic SEM; F1 and F3 were the only factors with a notable inter-factor genetic correlation (r_g_ = 0.44). The genetic link between F3 and atrial fibrillation recapitulates the F3 pheWAS result (Fig. 4c) highlighting the shared genetics underlying an association between taller stature and increased risk of atrial fibrillation.^27^ F2 had positive genetic correlations with the components of MetSyn and smaller genetic correlations relating to substance use, but weak correlations otherwise. F4 was again the most central factor in terms of the strength and quantity of genetic correlations, including positive correlations with metabolic disorders, pain, internalizing disorders, general risk-tolerance, attention-deficit hyperactivity disorder, substance use disorders, frailty, adult-onset asthma, coronary artery disease, and gout. F4 was negatively correlated with measures of fitness/exercise, compulsive disorders, HDL cholesterol, alcohol consumption frequency, and sleep efficiency. Interesting and nuanced relationships emerged between adipose genetic factors and mental health traits: general neuroticism was more genetically correlated with F2 (r_g_ = 0.18) compared to F4 (-0.01), but the depressed affect and worry subtypes of neuroticism were more genetically correlated with F4 (0.20 and -0.21, respectively) compared with F2 (0.12 and 0.10, respectively). Thus, we found opposite directionality of the genetic correlation between F4 and the neuroticism subtypes and also between F4 and compulsive disorders (e.g., obsessive compulsive disorder [-.25] and anorexia nervosa [-.27]) versus internalizing disorders (e.g., anxiety disorders [.12] and major depressive disorder [.14]). Together, this suggests that the relationship of adiposity and mental health outcomes depends in part on which aspect of body composition is evaluated, and in turn, the possible physiological and neurological systems involved.

**Figure 6:**
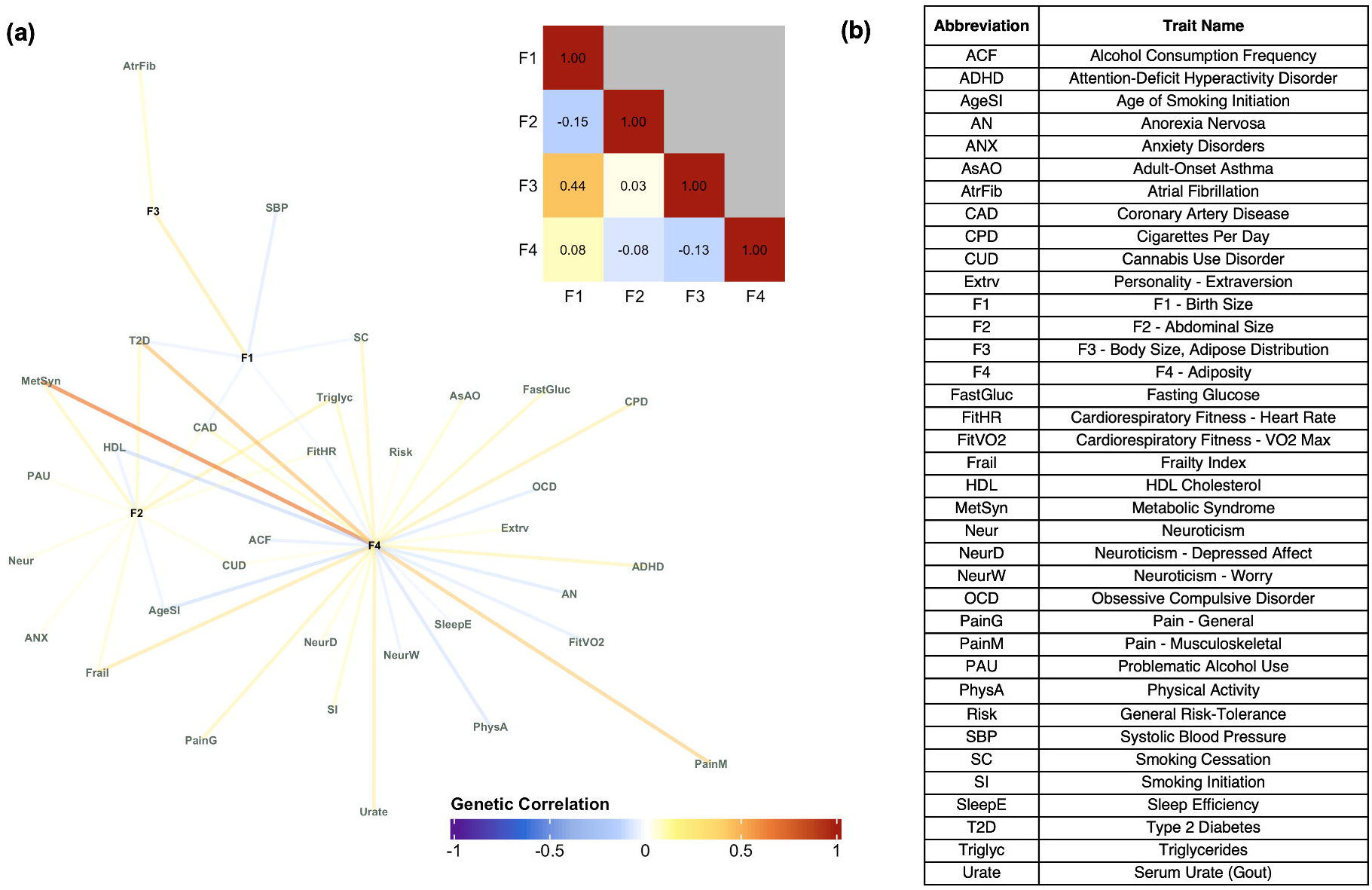
Network of genetic correlations with the 4 factors. A network describing the genetic correlations between the 4 Genomic SEM factors and a variety of genetically related traits is shown in panel (a); pairwise correlations ≤ 0.15 were pruned from the network. An inset correlation matrix for the 4 factors illustrates the mostly distinct genetic components represented by the factors. Abbreviations corresponding to the full trait names are described in panel (b) to assist with interpretation.

### Drug-gene network

Our final downstream analysis aimed to identify potential therapeutics that might ameliorate or prevent adipostiy by querying the significantly associated genes across two drug-gene interaction databases (Drug Repurposing Hub [DRH] and Drug-Gene Interaction Database [DGIdb]). We constructed a bipartite drug-gene network for each of the latent factors to assess the druggable genome in the context of our 4-factor model. Given the extensive phenotypic associations we observed for the PRS trained on the 4^th^ factor (Fig. 5c), we primarily focused on F4’s 1,239 DEPICT- or FOCUS-identified genes (results for the other three factor-identified genes are presented in Supplementary Tables 53-60). Our bipartite network for F4 included 733 drug-gene pairs (451 from the DRH, 192 identified by the DGIdb, and 90 identified by both), consisting of 151 genes and 529 drugs with regulatory approval. Of these 529 drugs, a substantial number (148) had prior descriptions of weight-related adverse drug events (wADEs) in the OnSIDES database.^28^ The 381 drugs without wADEs typically interacted with genes that were connected to drugs with known wADEs (Extended Data Figs. 5-10).

This network had groups of drugs clustered around high-degree genes, and drugs that served as links between different modules. Upon annotation of these drug clusters, we identified parts of the network that were specific to psychiatry, neurology, cardiology, oncology, endocrinology, and gastroenterology illustrating the diversity of therapeutics with potential wADEs based on interactions with F4-associated target genes. This analysis identified drug-gene pairs for serotonergic (e.g., trazodone) and dopaminergic agents (e.g., quetiapine) – well-known psychiatric medication classes with wADEs, sulfonylureas – diabetes medications with known wADEs, and tirzepatide – a potent weight loss and diabetes medication that interacts with *GIPR*. In addition, the drug-gene network for F3 recapitulated the function of fenofibrate as a therapeutic for MetSyn components^15^ via interactions with two significant genes (*SCARB1* and *GCKR*). These confirmatory results support the utility of our approach to identify novel and salient drug targets or existing drugs that might be repurposed to target adiposity. Moreover, genes interacting with drugs with known wADEs – e.g., antihistamines interacting with *HRH1* – frequently interacted with numerous other medications of the same drug class, suggesting weight-related drug effects may be under-recognized among medications with a common mechanism of action. Our bipartite network results can also be used to explore direct mechanisms for the drug-induced bodyweight changes that are commonly listed as adverse side effects of treatment and are observed in routine clinical care. For example, olanzapine (a psychiatric drug for schizophrenia and bipolar disorder), interacts with the same gene target as tirzepatide – *GIPR* – and this could explain the adverse weight gain often associated with olanzapine administration.^29–32^ In addition, the DEPICT GWAS identified muscarinic cholinergic receptor gene *CHRM4* and the FOCUS TWAS identified histamine receptor gene *HRH1* for F4 – these genes provide potential explanations for the wADEs of drugs that are used to treat mental disorders^33^ and antihistamine medications.^34^ Similarly, the identification of several receptor tyrosine kinases as having potentially causal effects on adiposity from the DEPICT and FOCUS analyses provides a mechanistic explanation for the wADEs of tyrosine kinase inhibitors.^35^ We also uncovered potentially novel drug-gene pairs that may inform studies of drug repurposing. One of the 45 genes that was identified by our DEPICT analyses for F4 but not for BMI was *PDE5A* on chromosome 12; this gene is targeted by dipyridamole (a medication used to prevent blood clots), which has been implicated as a potential therapeutic for weight loss via stimulating brown fat energy expenditure.^36^

## Discussion

Our 4-factor structural equation model serves as an informative and parsimonious representation of the genetic relationships among anthropometrics and adiposity across the lifespan. While many different measurements aim to quantify aspects of body size and body composition, our approach using correlated latent factors is less prone to the measurement error introduced by a singular phenotype definition, such as BMI. Furthermore, our modeling approach leveraged the combined power across indicator GWASs to identify novel genomic associations and provided a comprehensive mapping of the genetic architecture underlying birth size, abdominal size, adipose distribution, and adiposity. Our model highlighted differing genetic effects and loadings between males and females, and we characterized the distinct polygenic signals underlying each of the 4 genetic factors through various downstream analyses: multivariate GWASs, SNP-to-gene mapping, gene set enrichment, tissue enrichment, fine-mapped TWASs, PRS-based pheWASs, genetic correlations, and drug-gene interaction networks.

All of these analyses recapitulated the importance of F4, the adiposity factor, as the primary genetic culprit predisposing individuals to adverse health outcomes. Compared to the other 3 factors, F4 showed distinct enrichment for neuronal tissues and gene sets, stronger genetic correlations with related traits, broad health associations across numerous phenotypic domains, and relevant drug-gene pairings across diverse fields of medicine. Furthermore, F4 showed distinct genetic signal compared to BMI. The link between F4 and substance use traits is further accentuated by our identification of *GIPR* and tirzepatide in the drug-gene network because of the growing evidence for GIP and GLP-1 receptor agonists as potential anti-addiction treatments (beyond their primary indication for diabetes and weight loss).^37,38^ In the context of our ongoing search for more effective treatments, F4 provided possible mechanistic explanations for weight-related side effects across many medications and identified the potential for repurposed therapeurics to address adiposity (e.g., dipyridamole, an antiplatelet medication, which has been shown to target inosine as a stimulant of energy expenditure in brown adipocytes).^36,39^ The findings from our downstream analyses triangulated F4’s close relationship with behavioral traits through disentangling the genetic architecture of adiposity; the neuronal and behavioral context of F4 emphasized that the genetic loci associated with increased adiposity are underlain by complex relationships with environmental and lifestyle influences. F4 implicates a broad and cascading network of adiposity-mediated diseases^40^ and the underlying physiology of excess fat storage,^41^ adipokines (e.g., leptin and adiponectin),^42,43^ chronic inflammation from adipocyte apoptosis,^44^ MetSyn,^15^ and diabetes subtypes.^45,46^

Anthropometrics and adiposity across the lifespan have important health implications amidst a complex landscape of various patterns of inheritance (e.g., rare-vs-common genetic variants, high-vs-low penetrance, large-vs-small effect sizes)^2^ and diverse environmental contexts (e.g., food availability, physical activity, exposure to pollutants).^47–49^ The present analyses were limited to individuals of European ancestry, and future work will aim to characterize anthropometrics for additional ancestry groupings. In addition, our analyses share the strengths, assumptions, and limitations of the underlying methods including Genomic SEM,^19^ LDSC,^25,26^ DEPICT,^22^ and FOCUS.^23^ Extending our genetic insights into multi-ancestry, longitudinal, and multi-omics^50^ frameworks will enable the identification of biological markers beyond the genome and further disentangle the etiology of adipose-related diseases. While F4 had the strongest and most widespread health implications, the other three genetic factors characterized important aspects of body size and adipose distribution, reflecting unique influences on additional health outcomes, including respiratory illness,^51^ renal failure,^52^ hypertension,^53^ kidney stones^54^, T2D, and hyperlipidemia.^55–57^ Future directions might involve further exploration of the negative pheWAS association for F3 with hyperlipidemia, especially in the context of F3’s evidence for sex differences regarding depot-specific genetic architectures of adipose distribution.^58^

Our model describing the genetic associations for variation in human body size and body composition across the lifespan recapitulates the notion that food intake is not merely an unconditioned response to an energy deficiency, nor is it restricted to the canonical energy homeostasis areas in the brain (e.g., the hypothalamus).^59^ Instead, the involvement of brain areas performing the functions of sensory processing, learning, emotion, and memory indicates a broader neuro-centric genetic relationship with obesity. In this context, this neural component carries significant influence on health outcomes; and from a personalized medicine perspective, F4 has the promising capability to improve the prediction, diagnosis, treatment, and prevention of morbidities such as obesity, diabetes, adult persistent asthma, heart disease, chronic pain, substance use, and mental disorders.

## Online Methods

### Genomic structural equation modeling

Structural equation modeling is a widely used methodology for understanding the correlation and covariance patterns of interconnected variables. The resulting models are useful for explaining the variance of measurable variables, latent variables, and the relationships between those latent variables.^60^ We constructed an SEM describing the genetic associations of body size and body composition using a set of publicly available GWASs for various anthropometric traits. The measurement model that we constructed consisted of 18 individual GWAS summary statistics for 12 different phenotypes (described in Supplementary Table 1).^14,61–69^ Given our interest in investigating the sex-specific genetic architecture of body size and body composition, we included male and female GWASs independently for 6 of the 12 traits. The GWAS summary statistics were formatted using the `munge` function in the `GenomicSEM` R package after specifying the effect alleles, the effect sizes, standard errors, and sample sizes for each dataset. All 18 GWASs passed heritability-based quality control (QC) with heritability Z-statistics greater than 4, signifying they were well powered and had measurable effects across 954,086 overlapping genetic variants. These GWASs were comprised of European ancestry populations and the corresponding SNP reference file and linkage disequilibrium (LD) scores and were downloaded from the Genomic SEM data repository. The Ethics Board at the University of Colorado Boulder deemed that institutional review board approval was not necessary for our analyses as GWAS summary data do not include individual-level results; the studies that published the incorporated summary statistics obtained written informed consent from participants and were approved by local ethics committees.

The only binary trait included in the analysis was childhood obesity, which consisted of 9,116 cases and 13,292 controls; because this GWAS was a meta-analysis of multiple cohorts, the sum of effective sample sizes was used along with a sample prevalence of 0.5 (per the Genomic SEM multivariable LDSC function guidelines) and a population prevalence of 0.20 for liability scale conversion.^62^ We implemented the standard parameters for Genomic SEM, and QC criteria ensured the included SNPs were common (`maf.filter` = 0.01) and that the SNPs with lower imputation quality were removed from the analysis (`info.filter` = 0.9). When initially attempting to include all 3 bio-electrical impedance fat distribution GWASs (arm-fat-ratio [AFR], leg-fat-ratio [LFR], and trunk-fat-ratio [TFR]), the model showed poor fit and spurious standardized loadings greater than 1. This was due to the linear dependency among these 3 traits (the ratios of AFR, LFR, and TFR sum to 1, and therefore one ratio is predictable by the other two) which was problematic when inverting the sample covariance matrix in the process of computing the model estimates. We omitted LFR from the analysis since, LFR has many redundant genetic associations with TFR,^61^ and TFR was more relevant given the relationship between visceral adipose tissue and adverse health outcomes.

We implemented `Genomic` `SEM` in a 2-stage modeling process to fit an SEM to the genetic association estimates.^19^ We used multivariate linkage disequilibrium score regression (LDSC)^25,26^ to construct the genetic covariance (S_LDSC_) and sampling covariance (*V_SLDSC_*) matrices for the 18 GWAS summary statistics. Then, we fit an SEM using diagonally weighted least squares (DWLS) estimation. An important feature of Genomic SEM is that it is designed to handle varying degrees of sample overlap among the incorporated GWASs.

We first performed an exploratory factor analysis (EFA) by using odd chromosomes then a confirmatory factor analysis (CFA) using the even chromosomes to serve as a hold-out sample and protect from model overfitting. We used the Kaiser rule,^70^ the acceleration factor, and optimal coordinates criteria^71^ to assess the EFA and determine which eigenvalues of the genetic covariance matrix were most pronounced; all 3 criteria indicated that 4 latent factors was a judicious choice for the SEM. The `factanal` R package was used to perform a promax (i.e., correlated factor) rotation preceding the estimation of the unstandardized and standardized loadings from the nearest positive definite genetic covariance matrix via the `nearPD` function from the `matrix` R package. Variables with standardized loadings greater than 0.3 were specified to load onto each of the 4 latent factors, and the model structure was notably consistent for any threshold choice between 0.3 and 0.5. Heywood cases were handled for indicator variables with loadings close to 1 by constraining the residuals to be greater than 0.0001. The resulting fit of the SEM was evaluated using the comparative fit index (CFI) and the standardized root mean square residual (SRMR). Generally, CFI > 0.9 and SRMR < 0.1 are indicative of acceptable model fit for Genomic SEM models.^20,21^ WCadjBMI females and TFR males showed notable genetic correlations with indicator variables loading onto the 3^rd^ and 4^th^ factors respectively; including these cross-loadings improved model fit and resolved warnings regarding the covariance matrix of the residuals of the observed variables being non-positive definite. Ultimately, the CFA showed consistent factor structure with the EFA, and the overall measurement model achieved a reasonable balance between model fit and model parsimony. The resulting Genomic SEM model contained 4 factors and 127 degrees of freedom with a CFI = 0.94 and an SRMR = 0.11. After observing the generally distinct signals exhibited by these 4 factors and the poor model fit from a common factor model, we refrained from fitting a hierarchical factor model to the data.

### Genetic factors: multivariate genome wide association study

After defining the measurement model, we estimated SNP effects for the 4 genetic factors. This analysis was run in parallel for 954,086 SNPs that were common across the indicator GWASs and passed QC criteria. For each factor, we fit an independent pathways model for each SNP to test for heterogeneity of effect sizes among the indicator variables loading onto the same factor. The Genomic SEM Q_SNP_ methods included a `fix_measurement` parameter which was used to specify that the measurement model should be fixed across all SNPs, and we used the differences in the two models’ χ^2^ test statistics and degrees of freedom to identify SNPs with evidence for significant differences in model fit (Q_SNP_ p < 5×10^-8^).^19^ While these Q_SNPs_ are of interest because their indicator-specific effects might explain phenotypic divergence, for the purposes of constructing latent genetic factors that represent shared variance we removed these Q_SNPs_ along with nearby SNPs in LD. A European ancestry LD reference panel from the thousand genomes project (TGP)^72^ consisting of 503 unrelated individuals and 13.6 million genetic variants was implemented with `PLINK`^73,74^ to identify and filter variants within 1 mega-base and LD r^2^ ≥ 0.2 with the Q_SNPs_. F1, F2, F3, and F4 respectively had 23; 335; 1,525; and 969 significant Q_SNPs_, and after considering LD structure 79; 1,284; 6,909; and 4,183 SNPs were removed. The allele frequencies and the standard errors of the effect estimates were used to estimate the effective sample size for each of the 4 latent factors via the method described in the supplement of Mallard et al., 2022.^75^ F1, F2, F3, and F4 had estimated effective sample sizes of 52,404; 176,820; 690,110; and 393,268 respectively.

We used DEPICT^22^ v1.194 to identify independent, associated genomic loci using default parameters of p < 5×10^-8^, LD pairwise r^2^ < 0.1, and physical distance < 1 Mb (Supplementary Tables 19, 24, 29, and 34). These significantly associated independent loci were used as input for the following analyses included in the DEPICT framework. First, we performed DEPICT SNP-to-gene mapping to identify likely causal genes based on the assumption that genes within an associated locus have functional similarity to genes from other associated loci. This consisted of a scoring step (to quantify the similarity of gene set membership of genes near associated loci), a bias adjustment step (to control for gene length and data structure), and a false discovery rate (FDR) estimation step. Significantly prioritized genes with FDR < 0.05 were retained as likely causal genes for our downstream analyses and are included in Supplementary Tables 4, 7, 10, and 13 for each factor GWAS. Next, DEPICT was used to identify functional or phenotypic gene sets that are enriched in genes within associated loci. This was performed using DEPICT’s 10,968 reconstituted gene sets that are representative of a broad spectrum of biological annotations. Gene sets with enrichment nominal p-values less than the Bonferroni-corrected significance threshold (p < 4.56×10^-6^) are listed in Supplementary Tables 21, 26, 31 and 36 for each factor GWAS. Finally, DEPICT was implemented to test for enrichment (FDR < 0.05) of tissues or cell types for the associated loci which is described included in Supplementary Tables 22, 27, 32, and 37.

### Genetic factors: multivariate transcriptome wide association study

TWAS methods provided a natural extension of the multivariate GWASs to highlight genes with predicted expression that are putatively causal for the latent factors. We implemented TSEM and FOCUS softwares to perform transcription-level analyses of the previously discussed latent factor GWASs.^23,76,77^ Ultimately, the FOCUS framework was prioritized over TSEM in our TWAS analysis because the software’s fine-mapping approach handled the induced correlation structure for predicted gene expression and provided PIPs and credible sets of putatively causal genes. Although we do not discuss the TSEM results here, they are included in Supplementary Tables 61-72. Using FOCUS, we identified 90%-credible gene sets that excluded the null model (i.e., regions with strong evidence for modeled gene expression associating with the phenotype; Supplementary Tables 23, 28, 33, and 38). These credible sets were estimated using SNP LD structure, prediction eQTL weights, and the factor GWAS summary statistics. We used the FOCUS repository’s recommended European ancestry reference LD plink-formatted files from LDSC and the FOCUS repository’s multiple tissue, multiple eQTL reference panel weight database. First, the FOCUS munge functionality was used to format the factors’ GWAS summary statistics, and then each chromosome was run in parallel using independent genomic regions across European ancestry identified by LDetect^78^ and the prior probability for a gene to be causal as 0.001. The tissue-enrichment results from DEPICT revealed that the 4^th^ factor was the only factor with enrichment in a singular physiological system (enriched only for nervous tissues and cell types); thus, FOCUS was run tissue-agnostic for the first 3 factors (F1, F2, and F3), and was run tissue-prioritized for the ‘brain’ for the 4^th^ factor (F4). The 4 factors respectively had 86, 290, 737, and 562 LD blocks with identified 90%-credible gene sets, and 69, 243, 690, and 335 of those did not contain the null model, respectively. Among those gene sets that did not contain the null model, we retained genes with PIP > 0.1 to filter out low probability genes from our downstream analyses. This thresholding step resulted in 158; 676; 2,266; and 850 respective genes with putatively causal predicted gene expression effects for each of the 4 factors (Supplementary Tables 5, 8, 11, and 14).

### Genetic factors: genetic correlations

We evaluated the genetic correlations of the four factors with a broad range of obesity-related traits, using multivariate LDSC. Given the far-reaching spectrum of obesity-related health outcomes, we compiled a list of traits relating to psychopathology, risky behavior, neuroticism, diet, sleep, exercise, substance use, pain, frailty, dementia, inflammatory disease, autoimmune disease, cardiovascular disease, and metabolism. The full set of considered traits is described in Supplementary Tables 3 and 48 along with sample sizes, population prevalence, and heritability Z-statistics. The multivariate LDSC function in the `Genomic` `SEM` R package was used to estimate genetic covariances and correlations.

### Genetic factors: phenome wide association study of genetic risk scores

Polygenic risk score (PRS) SNP weights were estimated for the 4 factor GWASs using `LDpred2`.^24^ A random subset of 5,000 unrelated individuals of European ancestry from the UK Biobank were used as an LD reference panel. This LD reference panel was > 1,000 individuals per the `LDpred2` guidelines, and we ensured the individuals were unrelated via `gcta64` `-- grm-singleton` 0.05.^79^ Standard QC processes involved filtering SNPs based on Hardy-Weinberg equilibrium p > 1 × 10^−6^, genotyping rate > 0.99, minor allele frequency (MAF) > 1%, and filtering individuals based on heterozygosity within 3 standard deviations of the mean and sample missingness < 0.02. The ancestry matched remarkably well between the LD panel and the summary statistics and nearly all SNPs were retained when applying the LDpred2 standard deviation filter on SNPs (Supplementary Fig. 28). The `snp_ldpred2_auto` function in the `bigsnpr` package was used to generate LD-adjusted PRS weights for a sequence of causal variant thresholds (30 evenly spaced values on a logarithmic scale ranging from 1×10^-4^ to 0.9). The average of the betas for the models that converged were used for the PRSs resulting in 710,801; 710,489; 709,195; and 709,830 SNP weights for F1, F2, F3, and F4 respectively. Visualization of the raw GWAS effect sizes compared to the attenuated `LDpred2` adjusted PRS weights are included in (Supplementary Fig. 27).

These PRS weights were validated in an external dataset with no sample overlap with the included GWASs. We conducted 4 phenome-wide association studies (pheWASs) to investigate the associations between each of the 4 PRSs and all 1,591 phecode-based phenotypes in a cohort of unrelated Europeans from the Colorado Center for Personalized Medicine (CCPM) Biobank freeze2 (N = 25,240). Ancestry information was inferred based on the grouping of individuals’ genetic proximity to reference populations via PCA-UMAP (Principal Component Analysis-Uniform Manifold Approximation and Projection) projection and k-nearest neighbors methods. We excluded related individuals identified through KING-robust kinship estimates greater than 2×10^-3.5^, using the `bigsnpr` package in R.^80^ Details regarding the recruitment of CCPM Biobank participants, data processing, and the inference of population structures are described in Wiley et al., 2024.^81^

Our association model corrected for age, sex, batch, and the first 10 genetic principal components. To achieve unbiased estimates in the presence of case-control imbalance, we utilized the Saddlepoint approximation method from the *`SPAtest`* package in R.^82^ Due to the highly correlated structure of the phecodes, we considered associations with an FDR < 0.10 significant for characterizing the predictive signal of the factor PRSs. To evaluate the predictive utility of the F4 PRS conditioned on BMI, we ran an auxiliary pheWAS with BMI included as an additional covariate. To estimate BMI for each participant, we used the median of BMI measurements across the electronic health record. For each encounter with documented height (measured in inches) and weight (measured in ounces) we performed unit conversions and calculated the BMI as height/weight^2^. BMI values less than 13 kg/m^2^ or greater than 60 kg/m^2^ were removed before finding the median.

### Genetic factors: drug-gene network

We queried two large drug repurposing databases (Drug Repurposing Hub [DRH; 3/24/2020 version]^83^ and the Drug-Gene Interaction Database and the [DGIdb; 12/2023 version]) for the genes that were significantly prioritized by DEPICT for the independent GWAS loci (FDR < 0.05), or in non-null 90% credible sets identified by FOCUS (and PIP > 0.1) from the fine-mapped TWAS. There were 24; 319; 1,864; and 437 significantly prioritized DEPICT genes and 215; 862; 2,944; and 864 FOCUS fine-mapped genes for F1, F2, F3, and F4, respectively. The DGIdb contained drug-gene interaction scores reflecting strength of supporting publications and the relative drug-gene specificity. We filtered out drug-gene pairs with low interaction scores (< 0.50) based on the QC procedures described in similar studies.^84,85^ To map gene identifiers between datasets, we used the custom download feature from https://www.genenames.org/ to map the official gene symbol approved by the HGNC to the manually curated Ensembl Gene IDs. There were 14,472 drug-gene pairs for 6,798 drugs in the DRH and 19,819 drug-gene pairs for 8,037 drugs in the DGIdb. For visualization^86^ of the drug-gene network for F4 we removed drugs that did not have ‘launched’ clinical phase in the DRH or ‘approved’ status in the DGIdb. Drug indications were extracted from the ensemble MEDication Indication resource (MEDI-C)^87^ containing 38,378 high precision drug-indication pairs. The PheWAS R package^88^ was used to map the indication ICD10CM codes to phecodes and their corresponding phenotype domains. The ON-label SIDE effectS resource (OnSIDES, v2.0.0_20231113)^28^ was used to identify wADEs for the drugs in the network. This database contained 2,020 ingredients and 4,302 unique adverse reactions that were assigned using natural language processing models of drug labels. We considered drug-ADE pairs for which the adverse reaction was extracted from at least 75% of labels, and defined wADEs based on the following list of drug events: ‘Obesity’, ‘Central obesity’, ‘Weight increased’, ‘Weight decreased’, ‘Weight fluctuation’, ‘Abnormal loss of weight’, ‘Abnormal weight gain’, ‘Weight loss poor’, ‘Decreased appetite’, ‘Increased appetite’, ‘Appetite disorder’, ‘Hunger’, ‘Early satiety’, ‘Binge eating’, ‘Sleep-related eating disorder’, ‘Eating disorder’. Of these 16 terms, ‘Decreased appetite’, ‘Weight increased’, ‘Weight decreased’, and ‘Increased appetite’ were the most prominent and frequently observed (comprising 98% of wADE instances).

## Data Availability

All data included in this study were publicly available and citations linking downloads to the GWAS summary statistic files are included in the supplementary information. Multiavariate GWAS summary statistics for the 4 genetic factors will be made publicly available with the publication of this manuscript along with the LDpred2-derived PRS weights.

## Acknowledgements

We thank the generous feedback from members of the Institute for Behavioral Genetics (IBG) statistical genetics group. Data storage for this project was supported by the PetaLibrary and computational analysis was supported by the Blanca and Alpine high performance computing resources at the University of Colorado Boulder (funded by the University of Colorado Boulder, the University of Colorado Anschutz, and Colorado State University). CHA was supported by the IBG NIMH T32MH016880 training grant and by the Interdisciplinary Quantitative Biology program. RAG was supported by the University of Colorado Boulder’s Summer Multicultural Access to Research Training program, part of the Colorado Diversity Initiative, which is funded internally by the University of Colorado Boulder Graduate School. MAS was supported by grant K01 HL157658. ADG was supported by NIH Grants R01MH120219 and RF1AG073593. LME was supported by R01AG046938.

## Ethics declarations - Competing interests

There are no conflicts of interest related to this study.

## Data Availability

All data included in this study were publicly available and citations linking downloads to the GWAS summary statistic files are included in the supplementary information. Multiavariate GWAS summary statistics for the F1, F2, F3, and F4 can be downloaded from [data_repository_url] along with the LDpred2-derived PRS weights.

Code Availability [GitHub_repository_url]

---

**Extended Data Figure 1:**
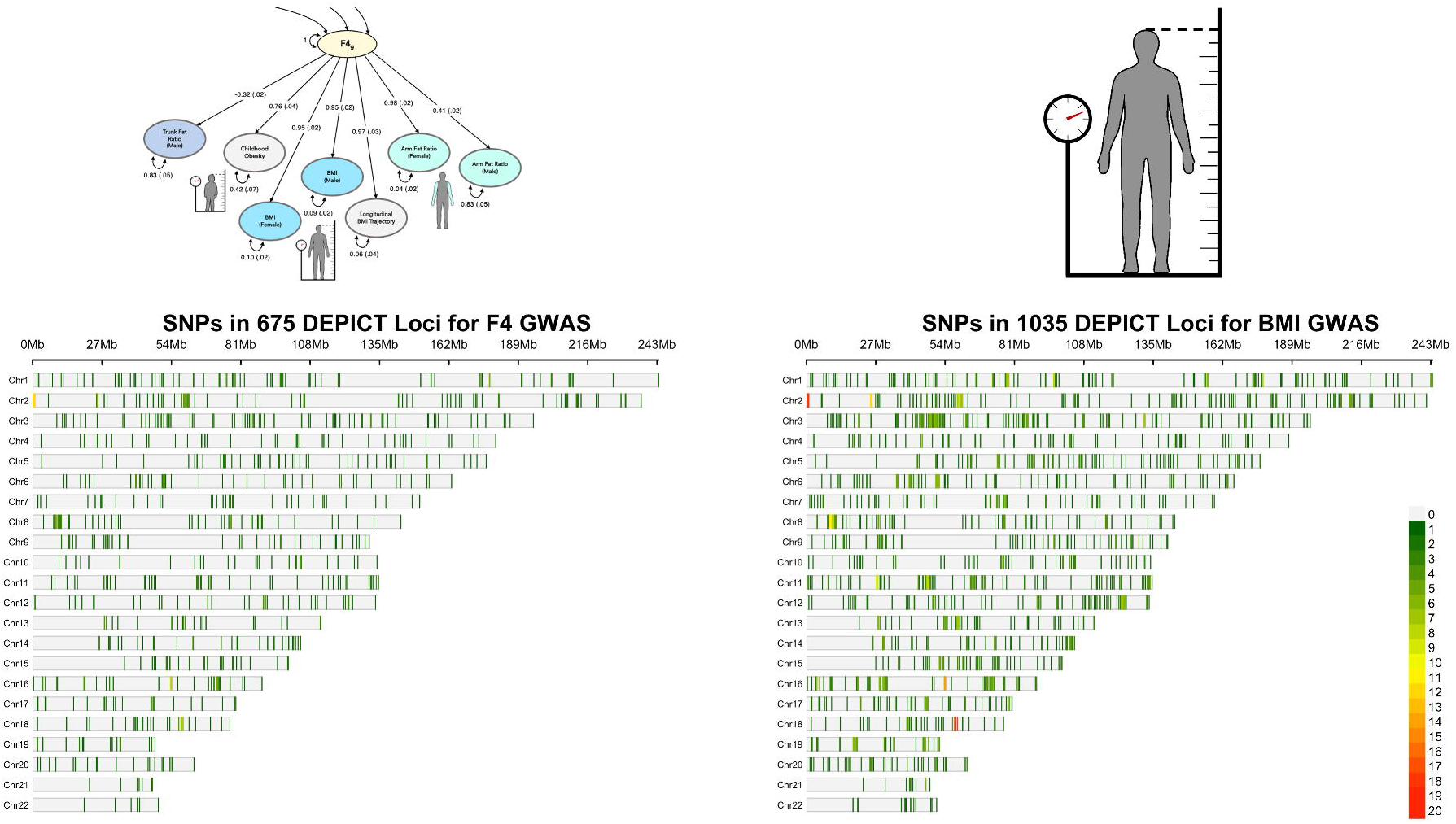
Comparison of F4 and BMI GWAS loci. A side-by-side chromosomal-map comparison of genome-wide significant (GWS; p < 5 x 10^-8^) genetic variants illustrating the shared and discordant associations across the 675 independent loci for F4 (left) and the 1,035 independent loci for BMI (right). 624 of the 675 F4 loci had genomic positional overlap with the BMI loci. The green-to-red shading describes density of significant GWAS SNPs in these loci across the 22 autosomes (bottom right color-scale).

**Extended Data Figure 2:**
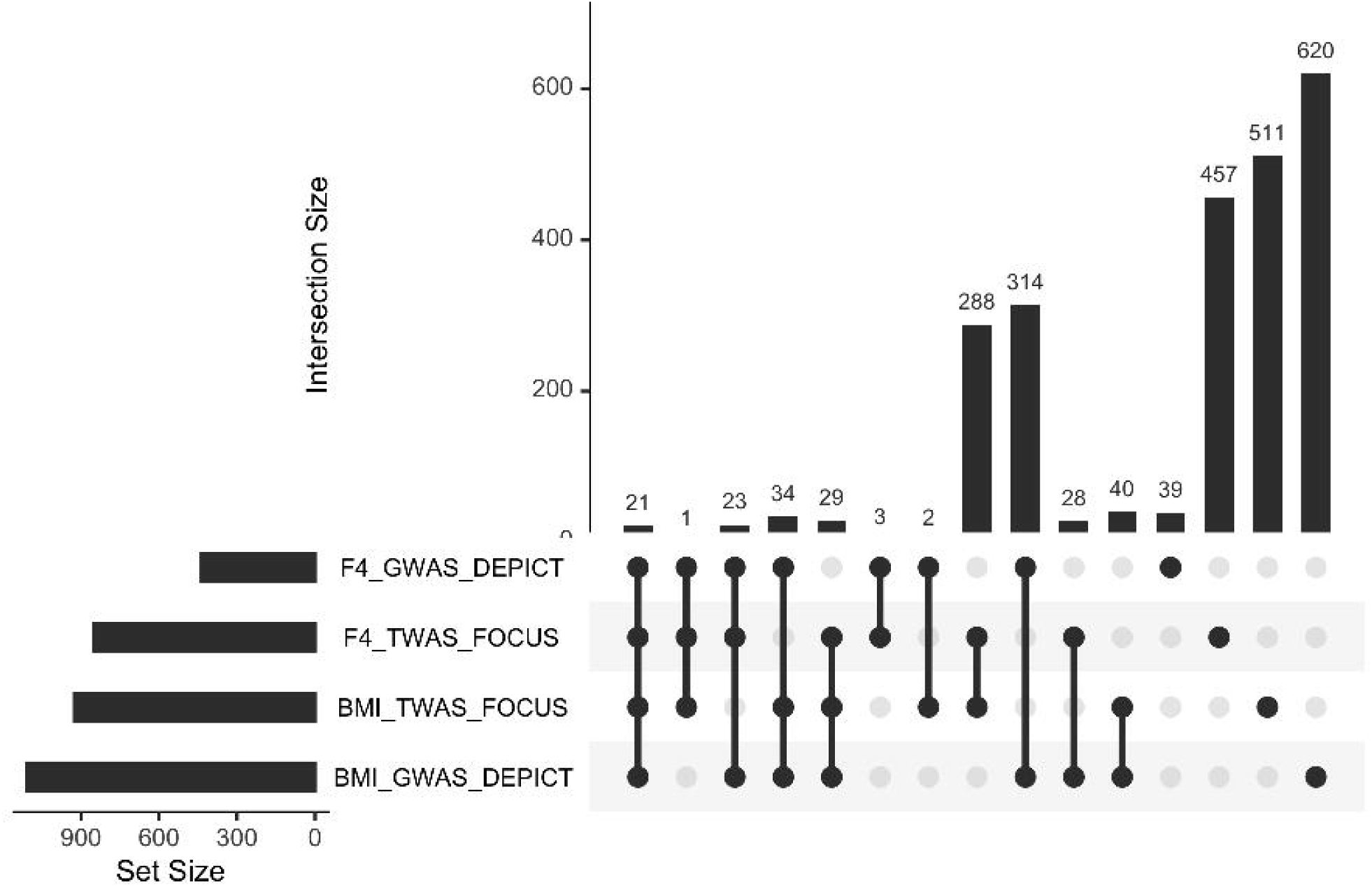
Comparison of identified genes for F4 and BMI. UpSet plot visualization^89^ of the set membership for identified genes in the DEPICT and FOCUS analyses of the F4 and BMI genome wide association studies (GWASs). Bars depicting set size in the bottom left show the number of genes identified by DEPICT (significantly prioritized genes from DEPICT with FDR < 0.05) and FOCUS (fine-mapped genes with PIP > 0.1 across non-null 90% credibles sets). The implicated genes from DEPICT describe variant-to-gene mapping from the GWAS, and the implicated genes from FOCUS describe genes with predicted expression that are putatively causal for the phenotype in a brain-tissue-prioritized transcriptome wide association study (TWAS). Genes identified by only 1 of the analyses are represented on the right, and genes with more overlap between sets are shown on the left.

**Extended Data Figure 3:**
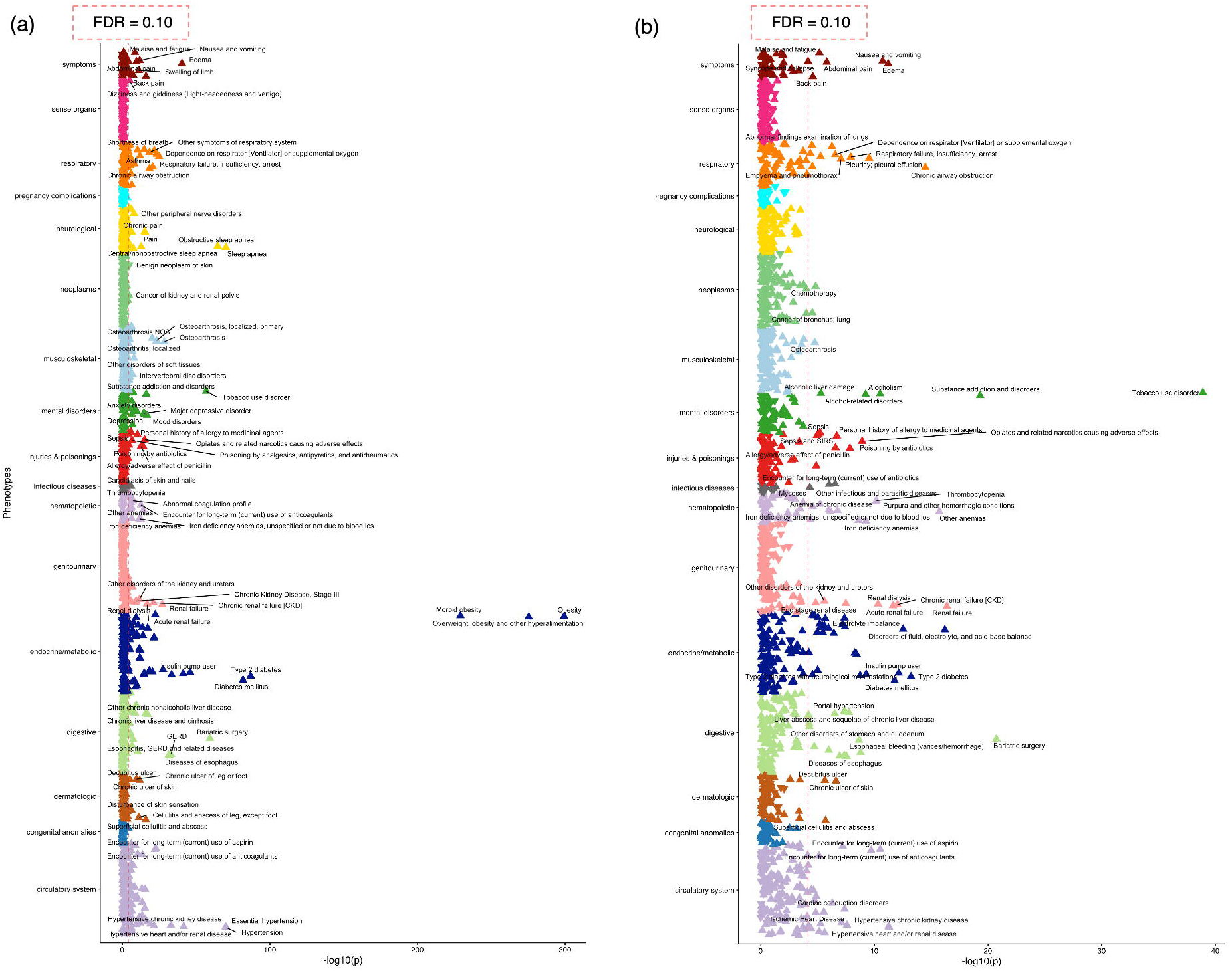
F4 pheWAS adjusted for BMI. The polygenic risk score (PRS) weights for F4 were validated in a phenome wide association study (pheWAS) in an external sample (N = 25,240); Panel (a) matches Figure 5c and is included alongside panel (b) for comparison to illustrate the pheWAS associations before (a) and after (b) adjusting for participant BMI. The significant associations between the PRS and phenotypes are labeled to the right of the vertical dashed red line denoting FDR < 0.10. Upward pointing triangles indicate a positive effect size between PRS and phenotype, and downward pointing triangles indicate a negative effect size.

**Extended Data Figure 4:**
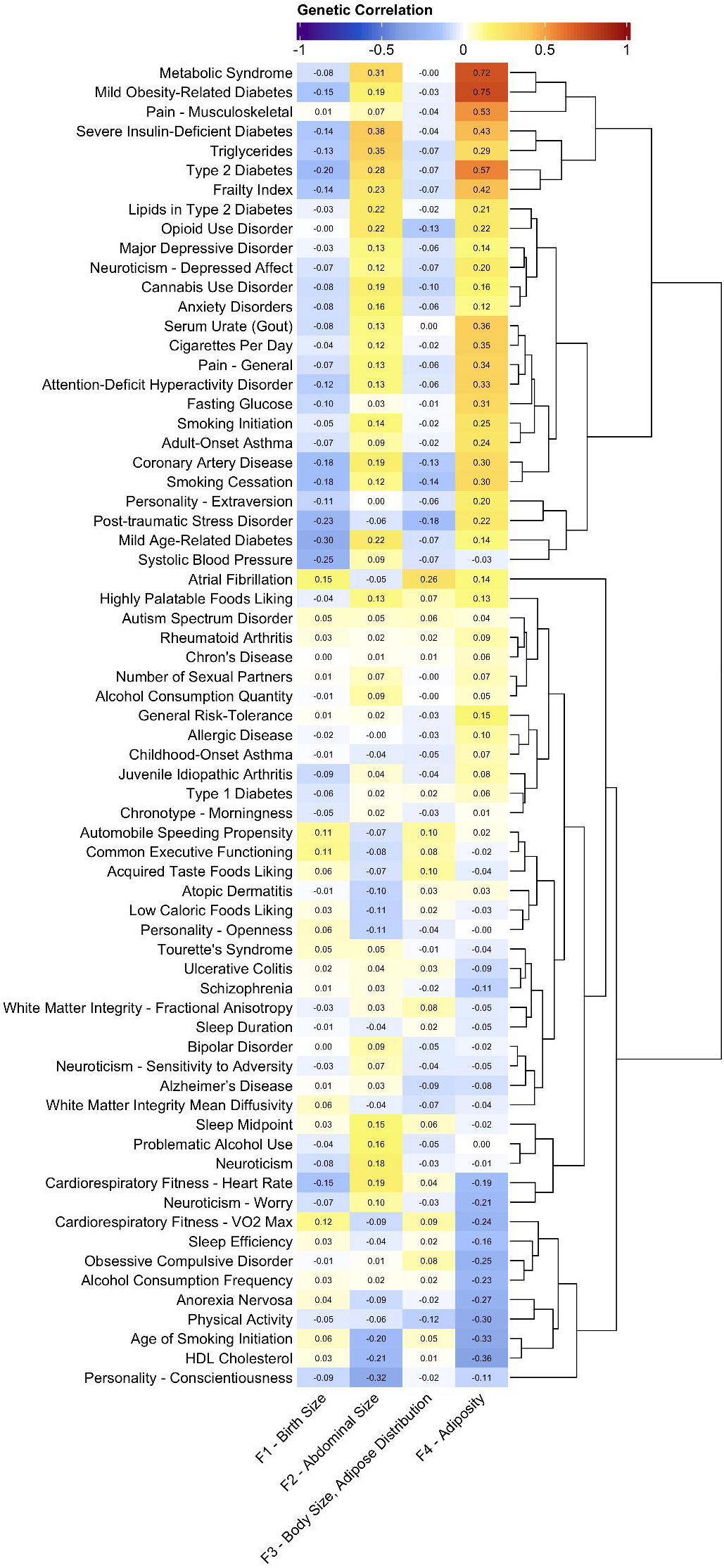
Genetic correlations with related traits. The genetic correlations between F1, F2, F3, and F4 and a variety of comorbidity-related traits. Rows of the heatmap^90^ are ordered in correspondence to the dendrogram along the right to cluster traits with similar relationships across the 4 factors. Genetic correlations greater than 0.15 are shown in Figure 6a, the full correlation matrix is shown in Supplementary Figure 49, standard errors are included in Supplementary Table 50, and information on the summary statistics is included in Supplemental Tables 3 and 48.

**Extended Data Figure 5:**
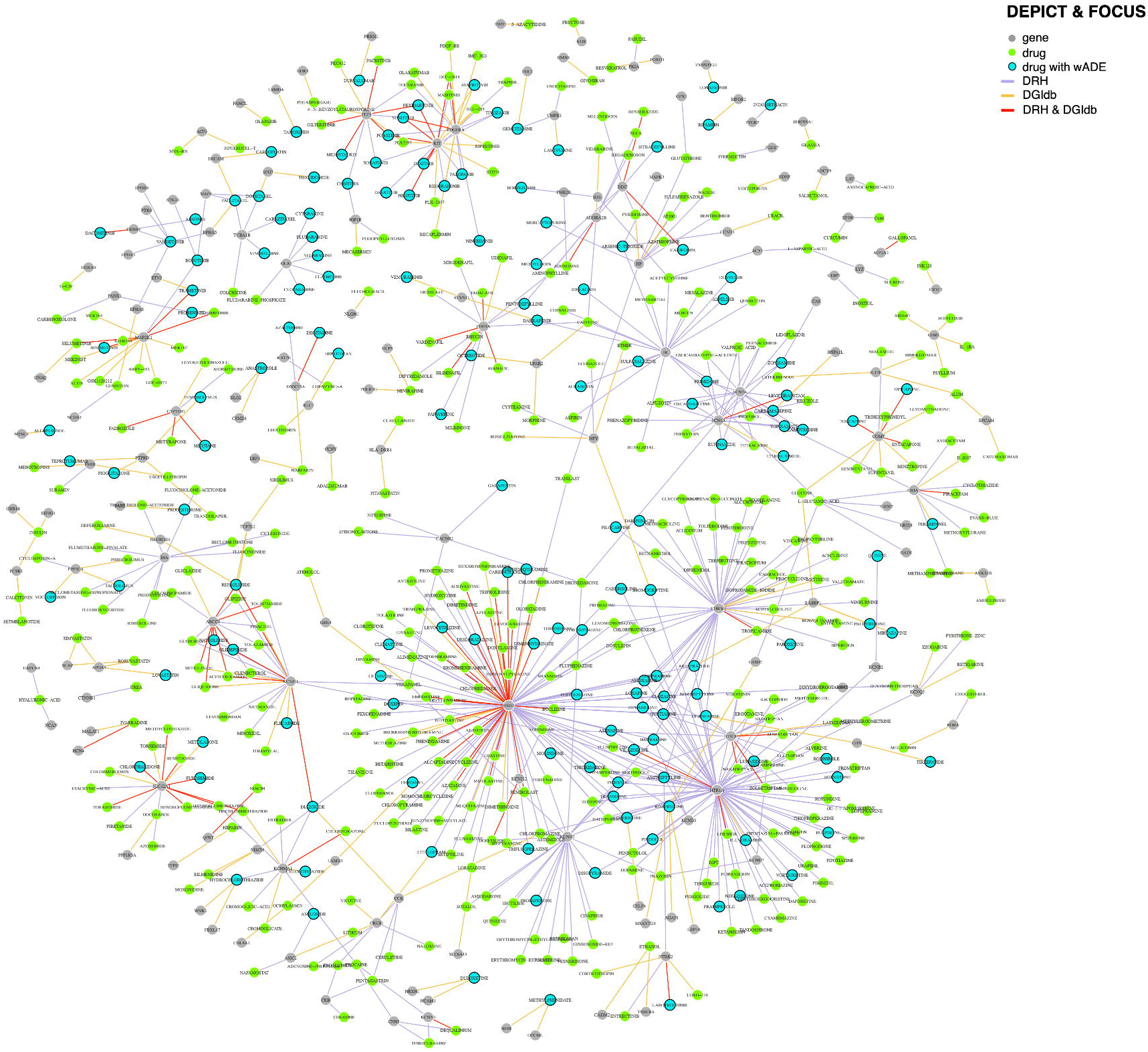
Drug-gene network for F4. The bipartite approved drug-gene network for significant genes in the GWAS DEPICT gene prioritization analysis or the FOCUS TWAS fine mapping analysis for F4 (the latent genetic factor relating to adiposity). For visualization of this network we removed drugs that did not have ‘launched’ clinical phase in the Drug Repurposing Hub (DRH) or ‘approved’ status in the Drug-Gene Interaction Database (DGIdb), for a total of 733 drug-gene pairs (451 identified in the DRH [purple edges], 192 identified in the DGIdb [orange edges], and 90 identified by both [red edges]) between 529 drugs and 151 genes significant for F4. The gene vertices are colored grey, and the drug vertices are colored green unless they had a weight-related adverse drug events (wADEs) listed in the ON-label SIDE effectS resource (OnSIDES) database (in which case they are colored blue with a black border).

**Extended Data Figure 6:**
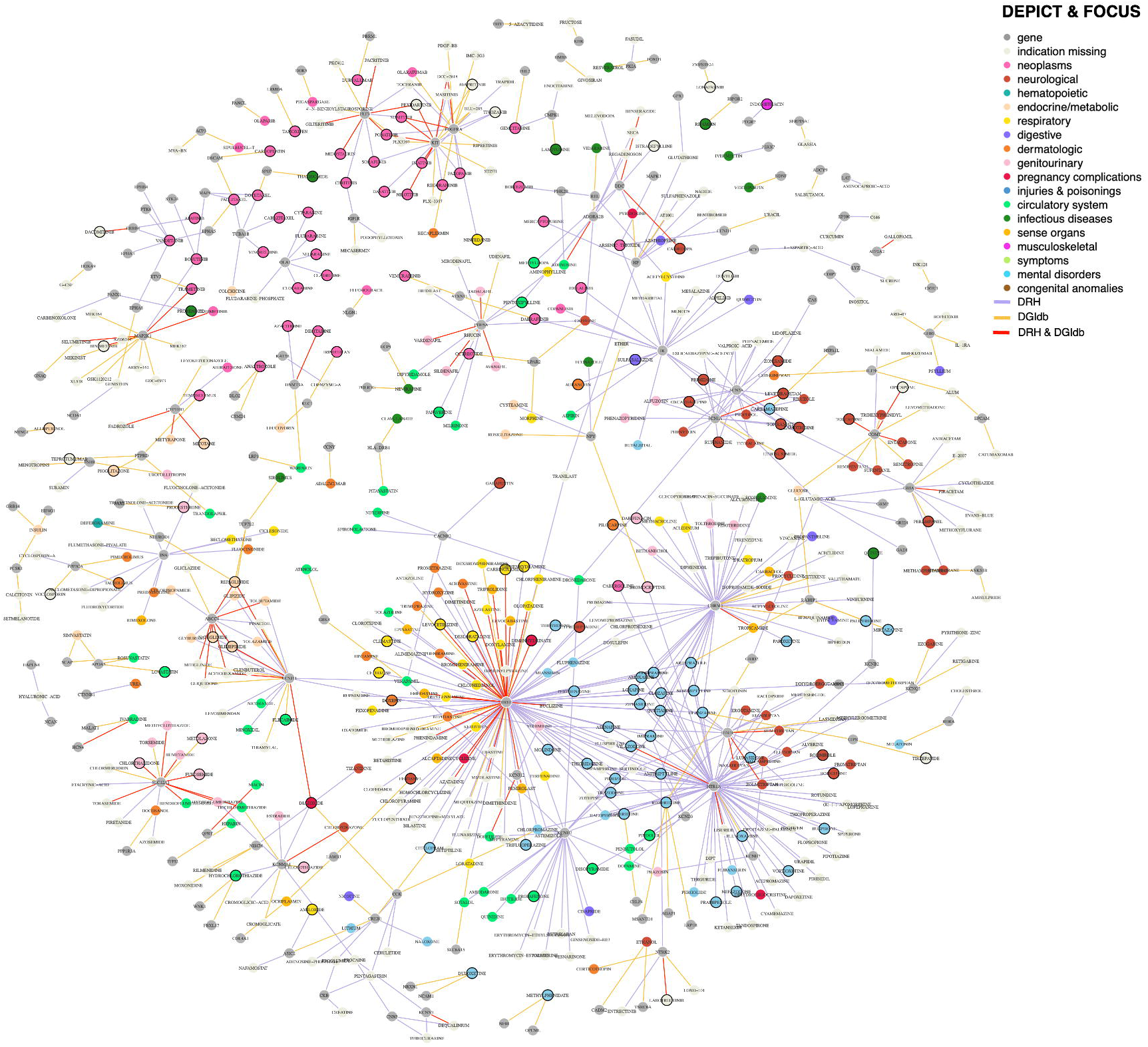
Drug-gene network for F4 with indications. The bipartite approved drug-gene network for significant genes in the GWAS DEPICT gene prioritization analysis or the FOCUS TWAS fine mapping analysis for F4 (the latent genetic factor relating to adiposity). For visualization of this network we removed drugs that did not have ‘launched’ clinical phase in the Drug Repurposing Hub (DRH) or ‘approved’ status in the Drug-Gene Interaction Database (DGIdb), for a total of 733 drug-gene pairs (451 identified in the DRH [purple edges], 192 identified in the DGIdb [orange edges], and 90 identified by both [red edges]) between 529 drugs and 151 genes significant for F4. The gene vertices are colored grey, and the drug vertices are colored by their most frequent indication category in the MEDI-C database. Drugs vertices with weight-related adverse drug events (wADEs) listed in the OnSIDES database have a black border.

**Extended Data Figure 7:**
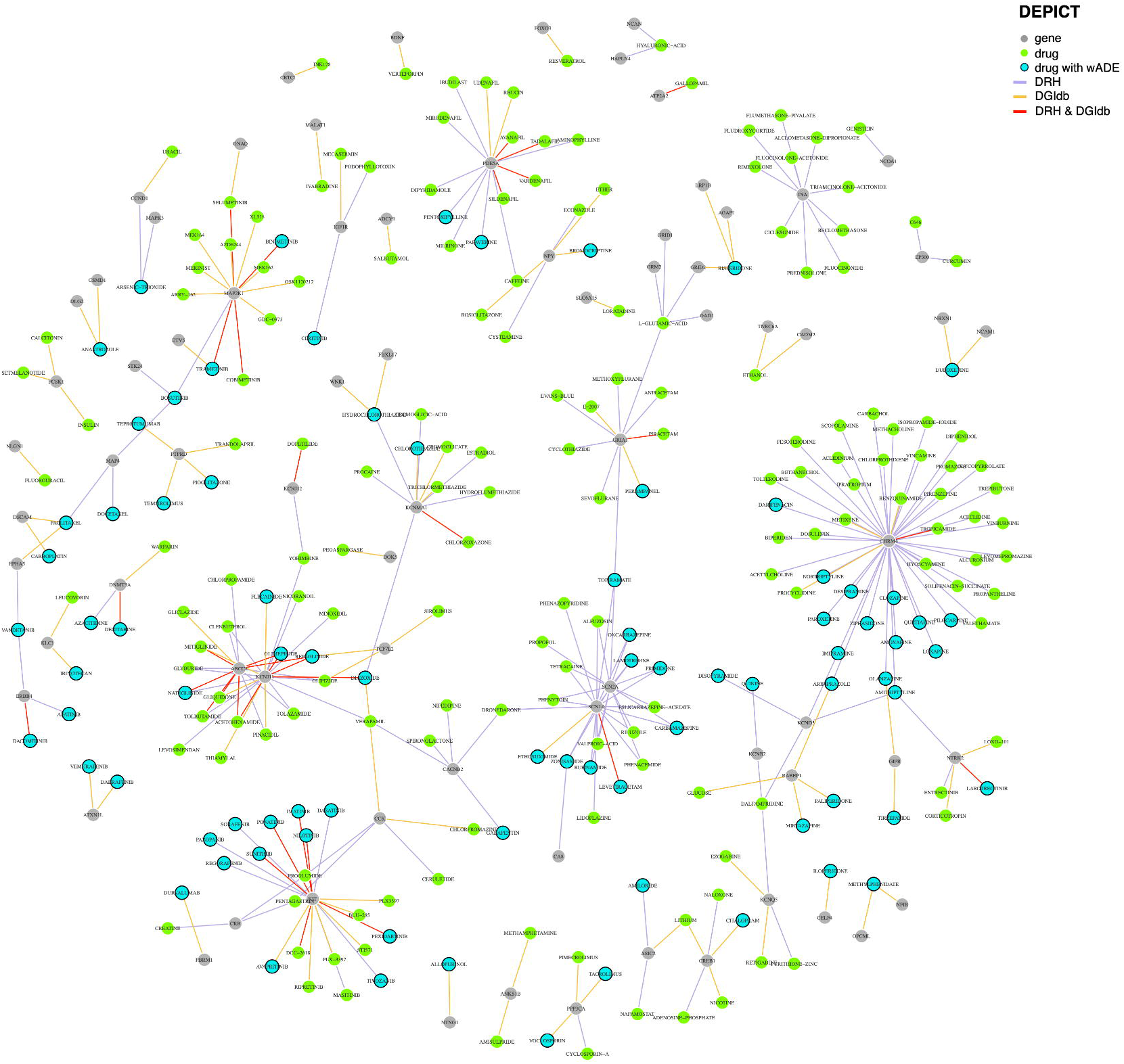
Drug-gene network for F4 (GWAS-identified genes). The bipartite approved drug-gene network for significant genes in the GWAS DEPICT gene prioritization analysis for F4 (the latent genetic factor relating to adiposity). For visualization of this network we removed drugs that did not have ‘launched’ clinical phase in the Drug Repurposing Hub (DRH) or ‘approved’ status in the Drug-Gene Interaction Database (DGIdb), for a total of 314 drug-gene pairs (179 identified in the DRH [purple edges], 101 identified in the DGIdb [orange edges], and 34 identified by both [red edges]) between 248 drugs and 75 genes significant for F4. The gene vertices are colored grey, and the drug vertices are colored green unless they had a weight-related adverse drug events (wADEs) listed in the ON-label SIDE effectS resource (OnSIDES) database (in which case they are colored blue with a black border).

**Extended Data Figure 8:**
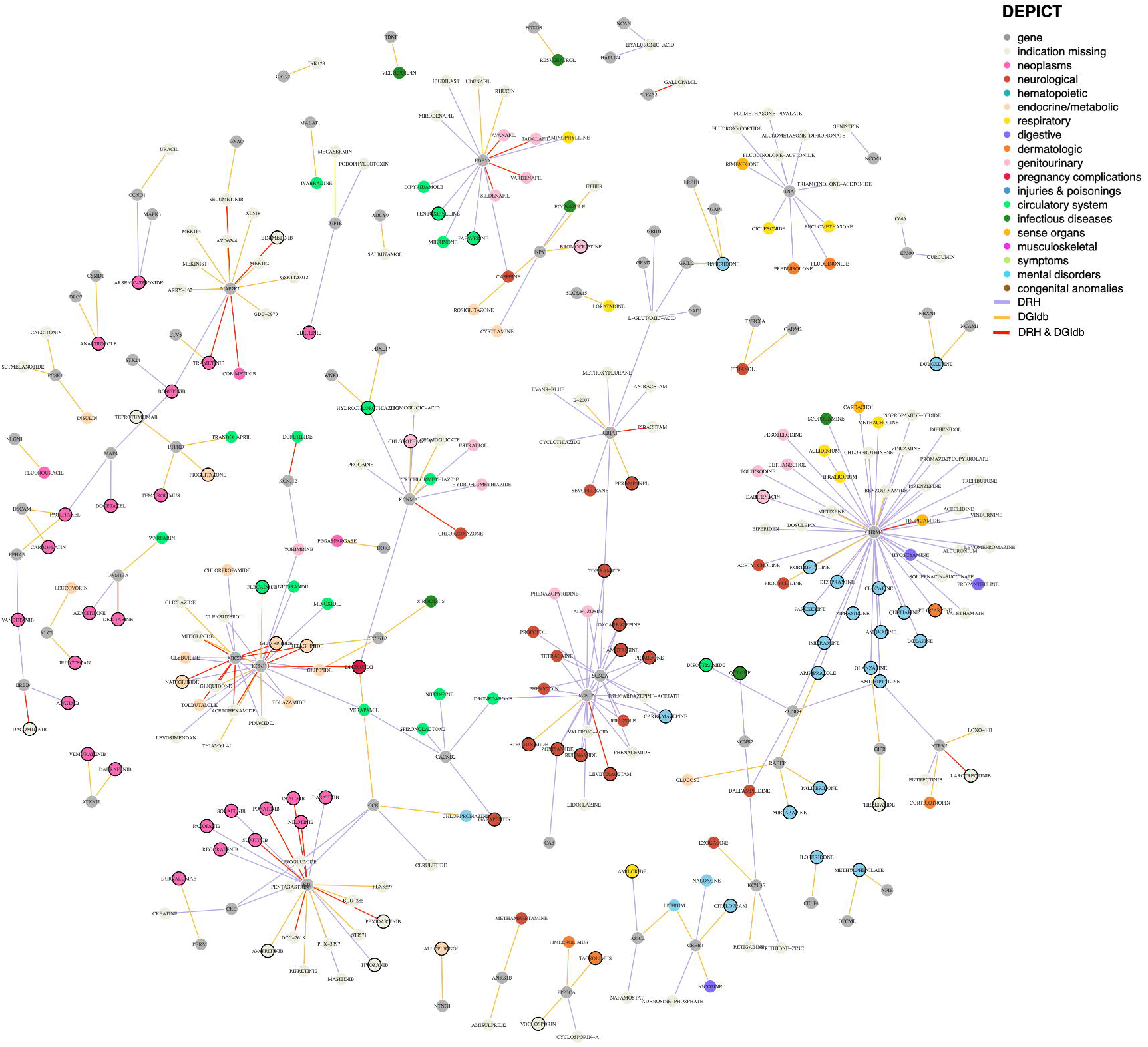
Drug-gene network for F4 with indications (GWAS-identified genes). The bipartite approved drug-gene network for significant genes in the GWAS DEPICT gene prioritization analysis for F4 (the latent genetic factor relating to adiposity). For visualization of this network we removed drugs that did not have ‘launched’ clinical phase in the Drug Repurposing Hub (DRH) or ‘approved’ status in the Drug-Gene Interaction Database (DGIdb), for a total of 314 drug-gene pairs (179 identified in the DRH [purple edges], 101 identified in the DGIdb [orange edges], and 34 identified by both [red edges]) between 248 drugs and 75 genes significant for F4. The gene vertices are colored grey, and the drug vertices are colored by their most frequent indication category in the MEDI-C database. Drugs vertices with weight-related adverse drug events (wADEs) listed in the OnSIDES database have a black border.

**Extended Data Figure 9:**
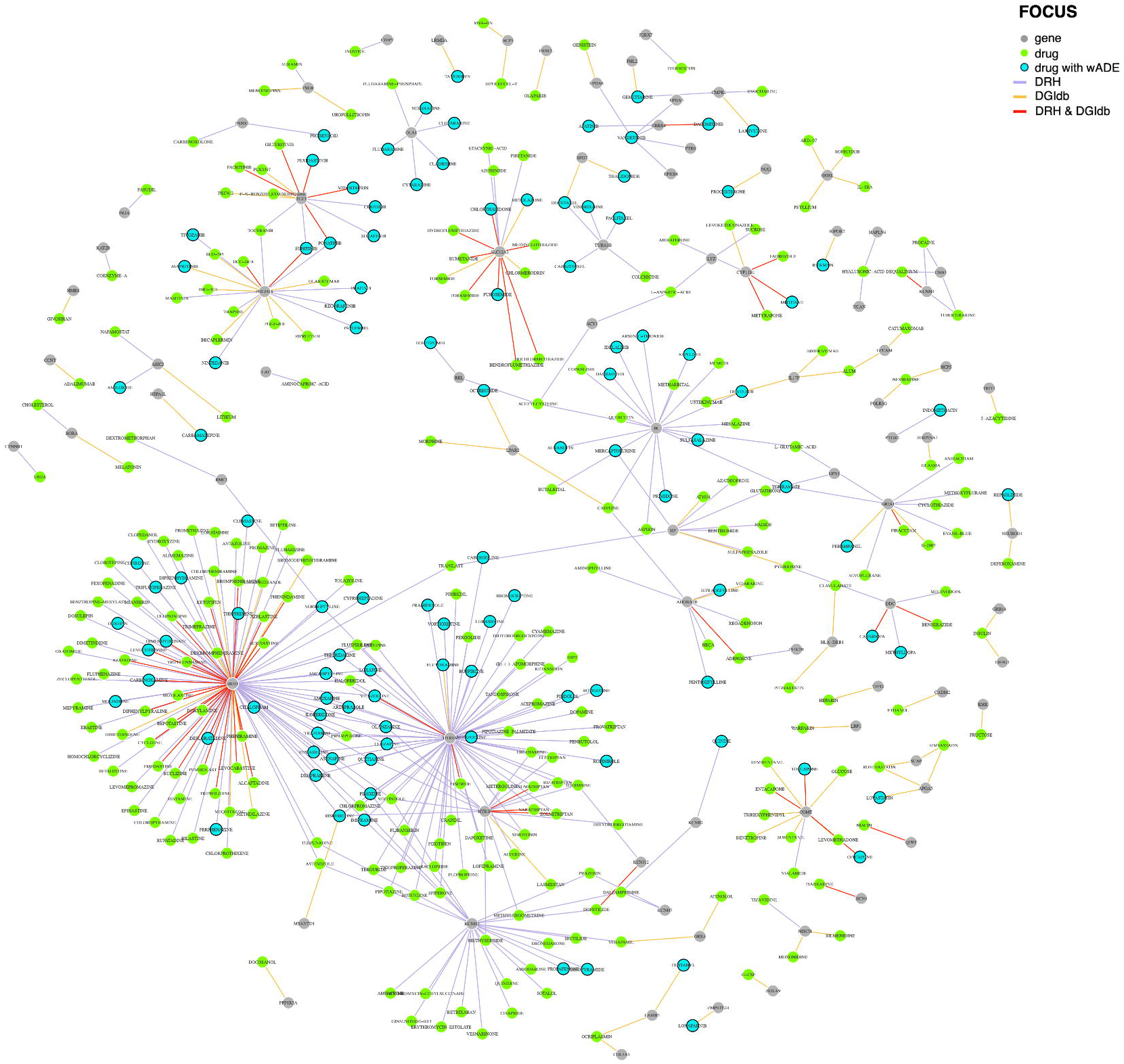
Drug-gene network for F4 (TWAS-identified genes). The bipartite approved drug-gene network for significant genes in the FOCUS TWAS fine mapping analysis for F4 (the latent genetic factor relating to adiposity). For visualization of this network we removed drugs that did not have ‘launched’ clinical phase in the Drug Repurposing Hub (DRH) or ‘approved’ status in the Drug-Gene Interaction Database (DGIdb), for a total of 442 drug-gene pairs (288 identified in the DRH [purple edges], 95 identified in the DGIdb [orange edges], and 59 identified by both [red edges]) between 360 drugs and 84 genes significant for F4. The gene vertices are colored grey, and the drug vertices are colored green unless they had a weight-related adverse drug events (wADEs) listed in the ON-label SIDE effectS resource (OnSIDES) database (in which case they are colored blue with a black border).

**Extended Data Figure 10:**
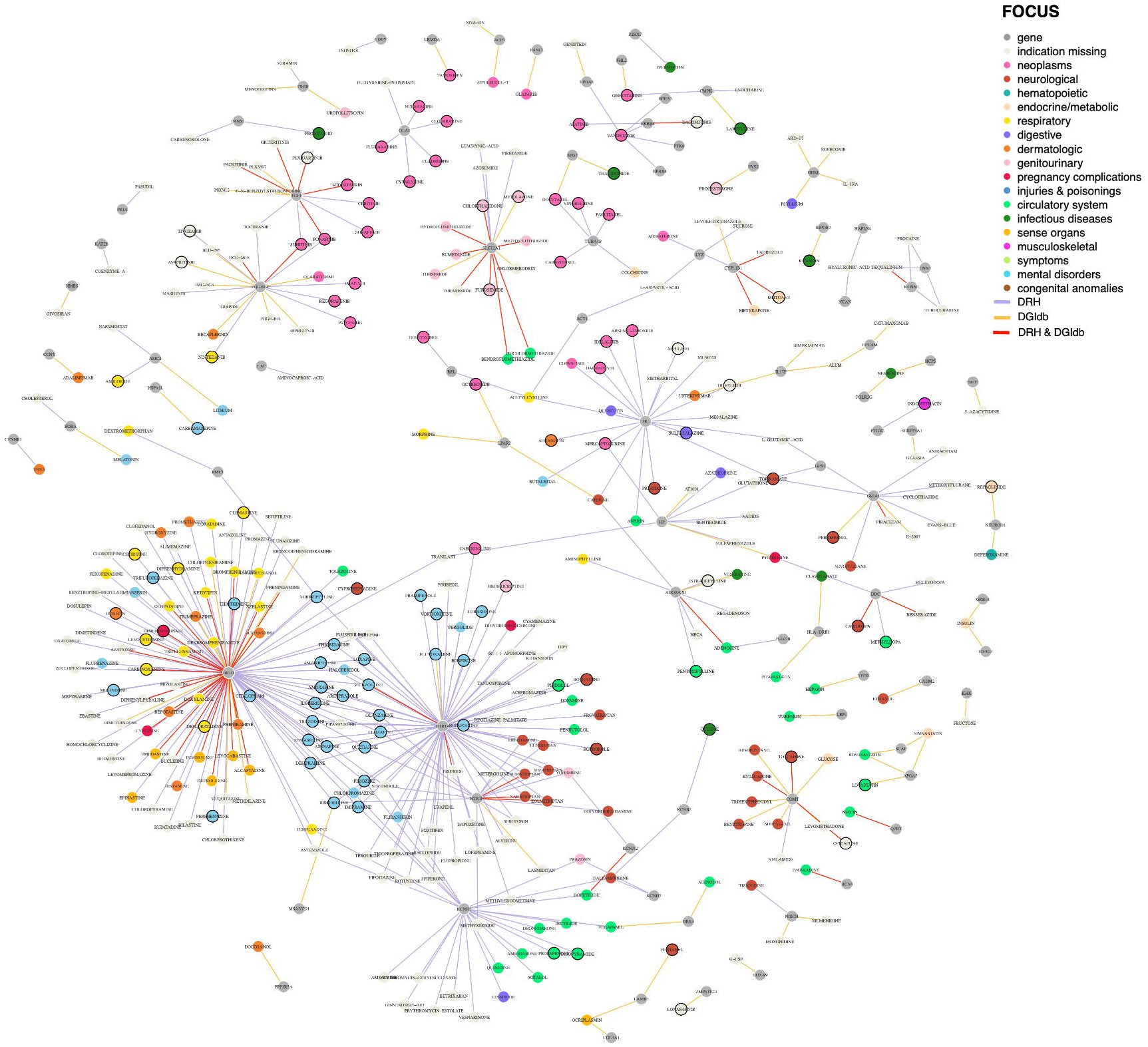
Drug-gene network for F4 with indications (TWAS-identified genes). The bipartite approved drug-gene network for significant genes in the FOCUS TWAS fine mapping analysis for F4 (the latent genetic factor relating to adiposity). For visualization of this network we removed drugs that did not have ‘launched’ clinical phase in the Drug Repurposing Hub (DRH) or ‘approved’ status in the Drug-Gene Interaction Database (DGIdb), for a total of 442 drug-gene pairs (288 identified in the DRH [purple edges], 95 identified in the DGIdb [orange edges], and 59 identified by both [red edges]) between 360 drugs and 84 genes significant for F4. The gene vertices are colored grey, and the drug vertices are colored by their most frequent indication category in the MEDI-C database. Drugs vertices with weight-related adverse drug events (wADEs) listed in the OnSIDES database have a black border.

## Notes

### Competing Interest Statement

The authors have declared no competing interest.

### Author Declarations

The Ethics Board at the University of Colorado Boulder deemed that institutional review board approval was not necessary for our analyses as GWAS summary data do not include individual-level results; the studies that published the incorporated summary statistics obtained written informed consent from participants and were approved by ethics committees.

## References

1. Bryan, S., et al. NHSR 158. National Health and Nutrition Examination Survey 2017–March 2020 Pre-Pandemic Data Files. https://stacks.cdc.gov/view/cdc/106273 (2021) doi:10.15620/cdc:106273.

2. Loos, R. J. F. & Yeo, G. S. H. The genetics of obesity: from discovery to biology. Nat. Rev. Genet. 23, 120–133 (2022).

3. CDC. Obesity is a Common, Serious, and Costly Disease. Centers for Disease Control and Prevention https://www.cdc.gov/obesity/data/adult.html (2022).

4. Khera, A. V. et al. Polygenic Prediction of Weight and Obesity Trajectories from Birth to Adulthood. Cell 177, 587–596.e9 (2019).

5. Hansen, G. T. et al. Genetics of sexually dimorphic adipose distribution in humans. Nat. Genet. 55, 461–470 (2023).

6. Ibrahim, M. M. Subcutaneous and visceral adipose tissue: structural and functional differences. Obes. Rev. 11, 11–18 (2010).

7. Aronica, L., Rigdon, J., Offringa, L. C., Stefanick, M. L. & Gardner, C. D. Examining differences between overweight women and men in 12-month weight loss study comparing healthy low-carbohydrate vs. low-fat diets. Int. J. Obes. 45, 225–234 (2021).

8. Doucet, E. et al. Reduction of visceral adipose tissue during weight loss. Eur. J. Clin. Nutr. 56, 297–304 (2002).

9. Okorodudu, D. O. et al. Diagnostic performance of body mass index to identify obesity as defined by body adiposity: a systematic review and meta-analysis. Int. J. Obes. 34, 791–799 (2010).

10. Tomiyama, A. J., Hunger, J. M., Nguyen-Cuu, J. & Wells, C. Misclassification of cardiometabolic health when using body mass index categories in NHANES 2005–2012. Int. J. Obes. 40, 883–886 (2016).

11. De Lorenzo, A. et al. Normal-weight obese syndrome: early inflammation? Am. J. Clin. Nutr. 85, 40–45 (2007).

12. Abdellaoui, A. & Verweij, K. J. H. Dissecting polygenic signals from genome-wide association studies on human behaviour. Nat. Hum. Behav. 5, 686–694 (2021).

13. Morys, F. et al. Neuroanatomical correlates of genetic risk for obesity in children. Transl. Psychiatry 13, 1 (2023).

14. The ADIPOGen Consortium et al. New genetic loci link adipose and insulin biology to body fat distribution. Nature 518, 187–196 (2015).

15. van Walree, E. S. et al. Disentangling Genetic Risks for Metabolic Syndrome. Diabetes 71, 2447–2457 (2022).

16. Virtue, S. & Vidal-Puig, A. Adipose tissue expandability, lipotoxicity and the Metabolic Syndrome--an allostatic perspective. Biochim. Biophys. Acta 1801, 338–349 (2010).

17. Yaghootkar, H. et al. Genetic Evidence for a Link Between Favorable Adiposity and Lower Risk of Type 2 Diabetes, Hypertension, and Heart Disease. Diabetes 65, 2448–2460 (2016).

18. Yaghootkar, H. et al. Genetic evidence for a normal-weight ‘metabolically obese’ phenotype linking insulin resistance, hypertension, coronary artery disease, and type 2 diabetes. Diabetes 63, 4369–4377 (2014).

19. Grotzinger, A. D. et al. Genomic structural equation modelling provides insights into the multivariate genetic architecture of complex traits. Nat. Hum. Behav. 3, 513–525 (2019).

20. Grotzinger, A. D. et al. Multivariate genomic architecture of cortical thickness and surface area at multiple levels of analysis. Nat. Commun. 14, 946 (2023).

21. Kaplan, D. Structural Equation Modeling (2nd Ed.): Foundations and Extensions. (SAGE Publications, Inc., 2009). doi:10.4135/9781452226576.

22. Pers, T. H. et al. Biological interpretation of genome-wide association studies using predicted gene functions. Nat. Commun. 6, 5890 (2015).

23. Mancuso, N. et al. Probabilistic fine-mapping of transcriptome-wide association studies. Nat. Genet. 51, 675–682 (2019).

24. LDpred2: better, faster, stronger | Bioinformatics | Oxford Academic. https://academic.oup.com/bioinformatics/article/36/22-23/5424/6039173.

25. Bulik-Sullivan, B. K. et al. LD Score regression distinguishes confounding from polygenicity in genome-wide association studies. Nat. Genet. 47, 291–295 (2015).

26. Bulik-Sullivan, B. et al. An atlas of genetic correlations across human diseases and traits. Nat. Genet. 47, 1236–1241 (2015).

27. Levin, M. G. et al. Genetics of height and risk of atrial fibrillation: A Mendelian randomization study. PLOS Med. 17, e1003288 (2020).

28. Tanaka, Y. et al. OnSIDES (ON-label SIDE effectS resource) Database: Extracting Adverse Drug Events from Drug Labels using Natural Language Processing Models. 2024.03.22.24304724 Preprint at 10.1101/2024.03.22.24304724 (2024).

29. Komossa, K. et al. Olanzapine versus other atypical antipsychotics for schizophrenia. Cochrane Database Syst. Rev. CD006654 (2010) doi:10.1002/14651858.CD006654.pub2.

30. Praharaj, S. K., Jana, A. K., Goyal, N. & Sinha, V. K. Metformin for olanzapine-induced weight gain: a systematic review and meta-analysis. Br. J. Clin. Pharmacol. 71, 377–382 (2011).

31. Ono, S. et al. Association between the GIPR gene and the insulin level after glucose loading in schizophrenia patients treated with olanzapine. Pharmacogenomics J. 12, 507–512 (2012).

32. Ono, S. et al. GIPR Gene Polymorphism and Weight Gain in Patients With Schizophrenia Treated With Olanzapine. J. Neuropsychiatry Clin. Neurosci. 27, 162–164 (2015).

33. Jeon, W. J., Dean, B., Scarr, E. & Gibbons, A. The Role of Muscarinic Receptors in the Pathophysiology of Mood Disorders: A Potential Novel Treatment? Curr. Neuropharmacol. 13, 739–749 (2015).

34. Simons, F. E. R. & Simons, K. J. Histamine and H1-antihistamines: Celebrating a century of progress. J. Allergy Clin. Immunol. 128, 1139–1150.e4 (2011).

35. Zhao, M., Jung, Y., Jiang, Z. & Svensson, K. J. Regulation of Energy Metabolism by Receptor Tyrosine Kinase Ligands. Front. Physiol. 11, (2020).

36. Niemann, B. et al. Apoptotic brown adipocytes enhance energy expenditure via extracellular inosine. Nature 609, 361–368 (2022).

37. Klausen, M. K., Thomsen, M., Wortwein, G. & Fink-Jensen, A. The role of glucagon-like peptide 1 (GLP-1) in addictive disorders. Br. J. Pharmacol. 179, 625–641 (2022).

38. Tsermpini, E. E., Goričar, K., Kores Plesničar, B., Plemenitaš Ilješ, A. & Dolžan, V. Genetic Variability of Incretin Receptors and Alcohol Dependence: A Pilot Study. Front. Mol. Neurosci. 15, 908948 (2022).

39. Willemsen, N., Kotschi, S. & Bartelt, A. Fire up the pyre: inosine thermogenic signaling for obesity therapy. Signal Transduct. Target. Ther. 7, 1–2 (2022).

40. Huang, J. et al. Genomics and phenomics of body mass index reveals a complex disease network. Nat. Commun. 13, 7973 (2022).

41. Emdin, C. A. et al. Genetic Association of Waist-to-Hip Ratio With Cardiometabolic Traits, Type 2 Diabetes, and Coronary Heart Disease. JAMA 317, 626–634 (2017).

42. Nakao, K. Adiposcience and adipotoxicity. Nat. Clin. Pract. Endocrinol. Metab. 5, 63–63 (2009).

43. Whitehead, J. P., Richards, A. A., Hickman, I. J., Macdonald, G. A. & Prins, J. B. Adiponectin--a key adipokine in the metabolic syndrome. Diabetes Obes. Metab. 8, 264–280 (2006).

44. Röszer, T. Adipose Tissue Immunometabolism and Apoptotic Cell Clearance. Cells 10, 2288 (2021).

45. Fuchsberger, C. et al. The genetic architecture of type 2 diabetes. Nature 536, 41–47 (2016).

46. Kim, H. et al. High-throughput genetic clustering of type 2 diabetes loci reveals heterogeneous mechanistic pathways of metabolic disease. Diabetologia 66, 495–507 (2023).

47. Qasim, A. et al. On the origin of obesity: identifying the biological, environmental and cultural drivers of genetic risk among human populations. Obes. Rev. 19, 121–149 (2018).

48. Alderete, T. L. et al. Ambient and Traffic-Related Air Pollution Exposures as Novel Risk Factors for Metabolic Dysfunction and Type 2 Diabetes. Curr. Epidemiol. Rep. 5, 79–91 (2018).

49. Alderete, T. L. et al. Prenatal traffic-related air pollution exposures, cord blood adipokines and infant weight. Pediatr. Obes. 13, 348–356 (2018).

50. Watanabe, K. et al. Multiomic signatures of body mass index identify heterogeneous health phenotypes and responses to a lifestyle intervention. Nat. Med. (2023) doi:10.1038/s41591023-02248-0.

51. Walter, E. C., Ehlenbach, W. J., Hotchkin, D. L., Chien, J. W. & Koepsell, T. D. Low Birth Weight and Respiratory Disease in Adulthood. Am. J. Respir. Crit. Care Med. 180, 176–180 (2009).

52. Lillås, B. S., Qvale, T. H., Richter, B. K. & Vikse, B. E. Birth Weight Is Associated With Kidney Size in Middle-Aged Women. Kidney Int. Rep. 6, 2794–2802 (2021).

53. Eriksson, J., Forsén, T., Tuomilehto, J., Osmond, C. & Barker, D. Fetal and childhood growth and hypertension in adult life. Hypertens. Dallas Tex 1979 36, 790–794 (2000).

54. Abufaraj, M. et al. Association Between Body Fat Mass and Kidney Stones in US Adults: Analysis of the National Health and Nutrition Examination Survey 2011-2018. Eur. Urol. Focus 8, 580–587 (2022).

55. Després, J. P. The insulin resistance-dyslipidemic syndrome of visceral obesity: effect on patients’ risk. Obes. Res. 6 **Suppl 1**, 8S–17S (1998).

56. Sironi, A. M. et al. Visceral fat in hypertension: influence on insulin resistance and beta-cell function. Hypertens. Dallas Tex 1979 44, 127–133 (2004).

57. Goswami, B., Reang, T., Sarkar, S., Sengupta, S. & Bhattacharjee, B. Role of body visceral fat in hypertension and dyslipidemia among the diabetic and nondiabetic ethnic population of Tripura—A comparative study. J. Fam. Med. Prim. Care 9, 2885–2890 (2020).

58. Agrawal, S. et al. Inherited basis of visceral, abdominal subcutaneous and gluteofemoral fat depots. Nat. Commun. 13, 3771 (2022).

59. Timshel, P. N., Thompson, J. J. & Pers, T. H. Genetic mapping of etiologic brain cell types for obesity. eLife 9, e55851 (2020).

60. Kline, R. B. Principles and Practice of Structural Equation Modeling. (The Guilford Press, New York, 2023).

61. Rask-Andersen, M., Karlsson, T., Ek, W. E. & Johansson, Å. Genome-wide association study of body fat distribution identifies adiposity loci and sex-specific genetic effects. Nat. Commun. 10, 339 (2019).

62. Bradfield, J. P. et al. A trans-ancestral meta-analysis of genome-wide association studies reveals loci associated with childhood obesity. Hum. Mol. Genet. 28, 3327–3338 (2019).

63. Vogelezang, S. et al. Genetics of early-life head circumference and genetic correlations with neurological, psychiatric and cognitive outcomes. BMC Med. Genomics 15, 124 (2022).

64. van der Valk, R. J. P. et al. A novel common variant in DCST2 is associated with length in early life and height in adulthood. Hum. Mol. Genet. 24, 1155–1168 (2015).

65. EGG Consortium et al. Maternal and fetal genetic effects on birth weight and their relevance to cardio-metabolic risk factors. Nat. Genet. 51, 804–814 (2019).

66. Pulit, S. L. et al. Meta-analysis of genome-wide association studies for body fat distribution in 694 649 individuals of European ancestry. Hum. Mol. Genet. 28, 166–174 (2019).

67. Yengo, L. et al. A saturated map of common genetic variants associated with human height. Nature 610, 704–712 (2022).

68. The Electronic Medical Records and Genomics (eMERGE) Consortium et al. Defining the role of common variation in the genomic and biological architecture of adult human height. Nat. Genet. 46, 1173–1186 (2014).

69. Ko, S. et al. GWAS of longitudinal trajectories at biobank scale. Am. J. Hum. Genet. 109, 433–445 (2022).

70. Kaiser, H. F. The Application of Electronic Computers to Factor Analysis. Educ. Psychol. Meas. 20, 141–151 (1960).

71. Ruscio, J. & Roche, B. Determining the number of factors to retain in an exploratory factor analysis using comparison data of known factorial structure. Psychol. Assess. 24, 282–292 (2012).

72. The 1000 Genomes Project Consortium et al. A global reference for human genetic variation. Nature 526, 68–74 (2015).

73. Purcell, S. et al. PLINK: a tool set for whole-genome association and population-based linkage analyses. Am. J. Hum. Genet. 81, 559–575 (2007).

74. Chang, C. C. et al. Second-generation PLINK: rising to the challenge of larger and richer datasets. GigaScience 4, 7 (2015).

75. Mallard, T. T. et al. Multivariate GWAS of psychiatric disorders and their cardinal symptoms reveal two dimensions of cross-cutting genetic liabilities. Cell Genomics 2, 100140 (2022).

76. Lu, Z. et al. Multi-ancestry fine-mapping improves precision to identify causal genes in transcriptome-wide association studies. Am. J. Hum. Genet. 109, 1388–1404 (2022).

77. Grotzinger, A. D., de la Fuente, J., Davies, G., Nivard, M. G. & Tucker-Drob, E. M. Transcriptome-wide and stratified genomic structural equation modeling identify neurobiological pathways shared across diverse cognitive traits. Nat. Commun. 13, 6280 (2022).

78. Berisa, T. & Pickrell, J. K. Approximately independent linkage disequilibrium blocks in human populations. Bioinformatics 32, 283–285 (2016).

79. Yang, J., Lee, S. H., Goddard, M. E. & Visscher, P. M. GCTA: a tool for genome-wide complex trait analysis. Am. J. Hum. Genet. 88, 76–82 (2011).

80. Privé, F., Aschard, H., Ziyatdinov, A. & Blum, M. G. B. Efficient analysis of large-scale genome-wide data with two R packages: bigstatsr and bigsnpr. Bioinformatics 34, 2781– 2787 (2018).

81. Wiley, L. K. et al. Building a vertically integrated genomic learning health system: The biobank at the Colorado Center for Personalized Medicine. Am. J. Hum. Genet. 111, 11–23 (2024).

82. Dey, R., Schmidt, E. M., Abecasis, G. R. & Lee, S. A Fast and Accurate Algorithm to Test for Binary Phenotypes and Its Application to PheWAS. Am. J. Hum. Genet. 101, 37–49 (2017).

83. Corsello, S. M. et al. The Drug Repurposing Hub: a next-generation drug library and information resource. Nat. Med. 23, 405–408 (2017).

84. Olla, S. et al. Combining Human Genetics of Multiple Sclerosis with Oxidative Stress Phenotype for Drug Repositioning. Pharmaceutics 13, 2064 (2021).

85. Grotzinger, A. D. et al. Transcriptome-Wide Structural Equation Modeling of 13 Major Psychiatric Disorders for Cross-Disorder Risk and Drug Repurposing. JAMA Psychiatry 80, 811–821 (2023).

86. Csárdi, G., et al. igraph for R: R interface of the igraph library for graph theory and network analysis. Zenodo 10.5281/ZENODO.7682609 (2024).

87. Zheng, N. S. et al. An updated, computable MEDication-Indication resource for biomedical research. Sci. Rep. 11, 18953 (2021).

88. Carroll, R. J., Bastarache, L. & Denny, J. C. R PheWAS: data analysis and plotting tools for phenome-wide association studies in the R environment. Bioinformatics 30, 2375–2376 (2014).

89. Lex, A., Gehlenborg, N., Strobelt, H., Vuillemot, R. & Pfister, H. UpSet: Visualization of Intersecting Sets. IEEE Trans. Vis. Comput. Graph. 20, 1983–1992 (2014).

90. Gu, Z., Eils, R. & Schlesner, M. Complex heatmaps reveal patterns and correlations in multidimensional genomic data. Bioinformatics 32, 2847–2849 (2016).

