## Supplementary_Materials for "Modeling the genomic architecture of adiposity and anthropometrics across the lifespan"

Supplementary Table 1: GWAS summary statistics included in the 4-factor genomic structural equation model. Each GWAS was conducted with samples of European ancestry. The median N column refers to the median sample size used to estimate effect sizes across the SNPs retained after quality control (QC).

| Trait | Continuous or Binary | $N_{SNPs}$ retained after QC | Median N | Download Source |
|---|---|---|---|---|
| Childhood obesity[1] | Binary | 6,094,124 | 21,224 | http://egg-consortium.org/ |
| Infant head circumference[2] | Continuous | 7,919,055 | 20,904 | http://egg-consortium.org/ |
| Birth length[3] | Continuous | 2,144,566 | 21,929 | http://egg-consortium.org/ |
| Birth weight[4] | Continuous | 7,944,591 | 281,141 | http://egg-consortium.org/ |
| Waist to hip ratio adjusted for BMI females[5] | Continuous | 7,531,295 | 262,759 | https://portals.broadinstitute.org/collaboration/giant/index.php/GIANT_consortium_data_files |
| Waist to hip ratio adjusted for BMI males[5] | Continuous | 7,531,295 | 221,804 | https://portals.broadinstitute.org/collaboration/giant/index.php/GIANT_consortium_data_files |
| Waist circumference adjusted for BMI females[6] | Continuous | 2,376,028 | 91,298 | https://portals.broadinstitute.org/collaboration/giant/index.php/GIANT_consortium_data_files |
| Waist circumference adjusted for BMI males[6] | Continuous | 2,233,846 | 61,075 | https://portals.broadinstitute.org/collaboration/giant/index.php/GIANT_consortium_data_files |
| Hip circumference adjusted for BMI females[6] | Continuous | 2,372,868 | 86,748 | https://portals.broadinstitute.org/collaboration/giant/index.php/GIANT_consortium_data_files |
| Hip circumference adjusted for BMI males[6] | Continuous | 2,133,665 | 56,944 | https://portals.broadinstitute.org/collaboration/giant/index.php/GIANT_consortium_data_files |
| BMI females[5] | Continuous | 7,533,667 | 262,817 | https://portals.broadinstitute.org/collaboration/giant/index.php/GIANT_consortium_data_files |
| BMI males[5] | Continuous | 7,533,667 | 221,863 | https://portals.broadinstitute.org/collaboration/giant/index.php/GIANT_consortium_data_files |

| | | | | |
|---|---|---|---|---|
| Height[7,8] | Continuous | 2,284,352 | 704,526 | https://www.joelhirschhornlab.org/giant-consortium-results |
| BMI mean trajectory (beta)[9] | Continuous | 5,871,415 | 144,414 | https://ucla.app.box.com/v/trajgwassummary |
| Bio-electrical impedance arm fat ratio females[10] | Continuous | 7,798,339 | 191,163 | https://myfiles.uu.se/ssf/s/readFile/share/3993/1270878243748486898/publicLink/GWAS_summary_stats_ratios.zip |
| Bio-electrical impedance arm fat ratio males[10] | Continuous | 7,800,166 | 163,201 | https://myfiles.uu.se/ssf/s/readFile/share/3993/1270878243748486898/publicLink/GWAS_summary_stats_ratios.zip |
| Bio-electrical impedance trunk fat ratio females[10] | Continuous | 7,798,337 | 191,132 | https://myfiles.uu.se/ssf/s/readFile/share/3993/1270878243748486898/publicLink/GWAS_summary_stats_ratios.zip |
| Bio-electrical impedance trunk fat ratio males[10] | Continuous | 7,800,166 | 163,158 | https://myfiles.uu.se/ssf/s/readFile/share/3993/1270878243748486898/publicLink/GWAS_summary_stats_ratios.zip |
| Bio-electrical impedance leg fat ratio females[10] | Continuous | 7,798,337 | 191,226 | https://myfiles.uu.se/ssf/s/readFile/share/3993/1270878243748486898/publicLink/GWAS_summary_stats_ratios.zip |
| Bio-electrical impedance leg fat ratio males[10] | Continuous | 7,800,166 | 163,250 | https://myfiles.uu.se/ssf/s/readFile/share/3993/1270878243748486898/publicLink/GWAS_summary_stats_ratios.zip |

Supplementary Table 2: Descriptive statistics for the multivariate factor GWASs and the downstream DEPICT and FOCUS analyses.

| GSEM Factor | F1: Birth Size | F2: Abdominal Size | F3: Body Size and Adipose Distribution | F4: Adiposity |
|---|---|---|---|---|
| Estimated effective sample size | 52,404 | 176,820 | 690,110 | 393,268 |
| N significant $Q_{SNPs}$ ($Q_{SNP}$ p < $5x10^{-8}$) | 23 | 335 | 1,525 | 969 |
| N SNPs filtered after accounting for LD with significant $Q_{SNPs}$ (LD $r^2 \geq 0.2$) | 79 | 1,284 | 6,909 | 4,183 |
| N SNPs after liftover to GRCh38 and filtering of $Q_{SNPs}$ | 953,681 | 953,287 | 951,752 | 952,497 |
| N SNPs novel relative to the indicator GWASs (p < $5x10^{-8}$) | 103 | 1,318 | 8 | 6,206 |
| N DEPICT independent GWAS loci | 88 | 344 | 1,173 | 675 |
| N DEPICT significantly prioritized genes (FDR < 0.05) | 24 | 319 | 1,864 | 437 |
| N FOCUS non-null 90% credibles sets | 69 | 243 | 689 | 335 |
| N FOCUS finemapped genes with PIP > 0.1 in non-null 90% credibles sets | 158 | 676 | 2,266 | 850 |

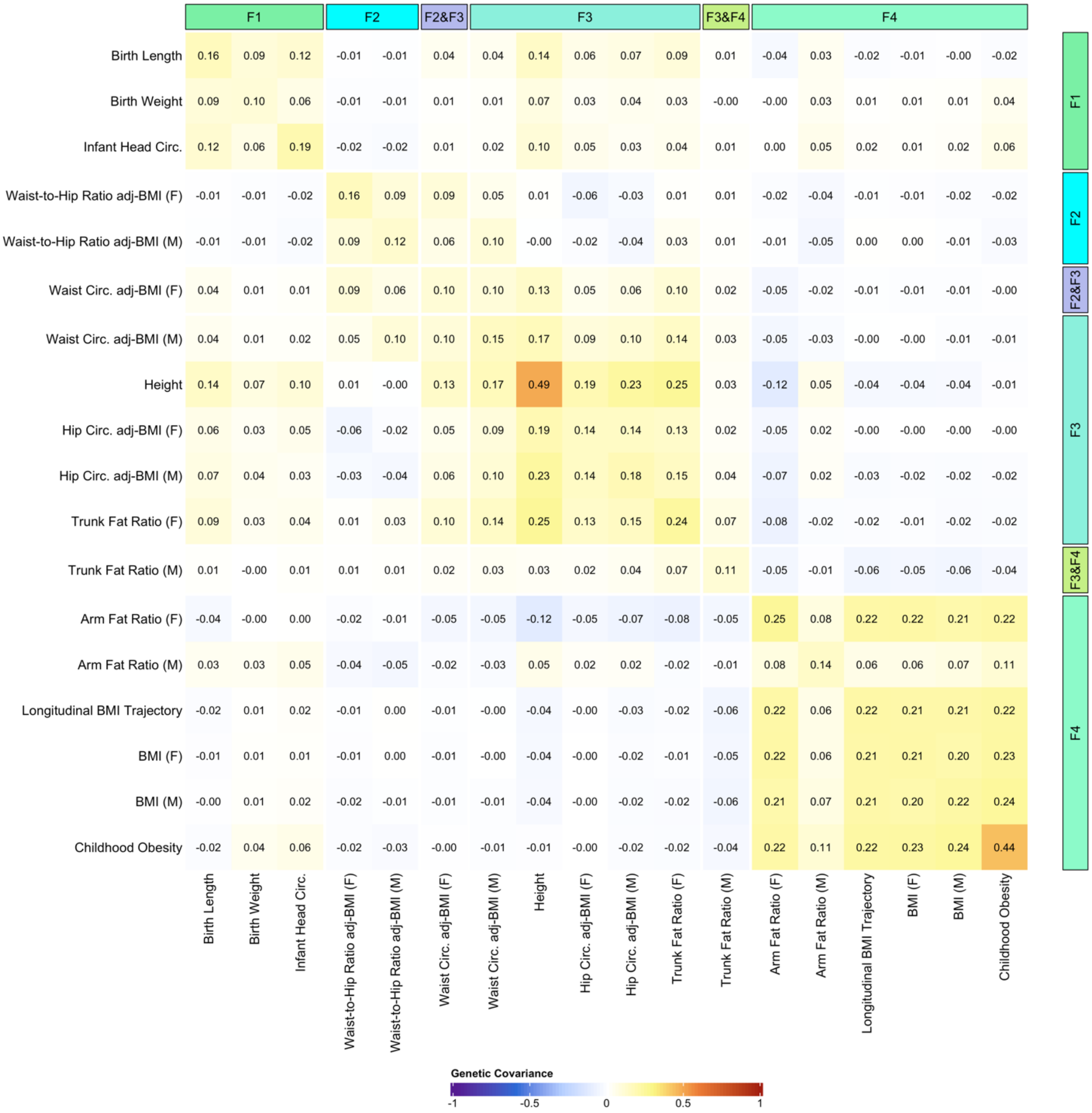


Supplementary Figure 1: The genetic covariance matrix of indicator variables included in the genomic structural equation model. SNP-based heritability estimates ($h^2$) are represented along the diagonal. Symmetrical annotations along the top and right sides of the matrix describe the specified loadings onto F1, F2, F3, and F4 for the measurement model. Among the 18 indicator variables, 6 of the included traits were stratified by sex with (M) denoting male and (F) denoting female.

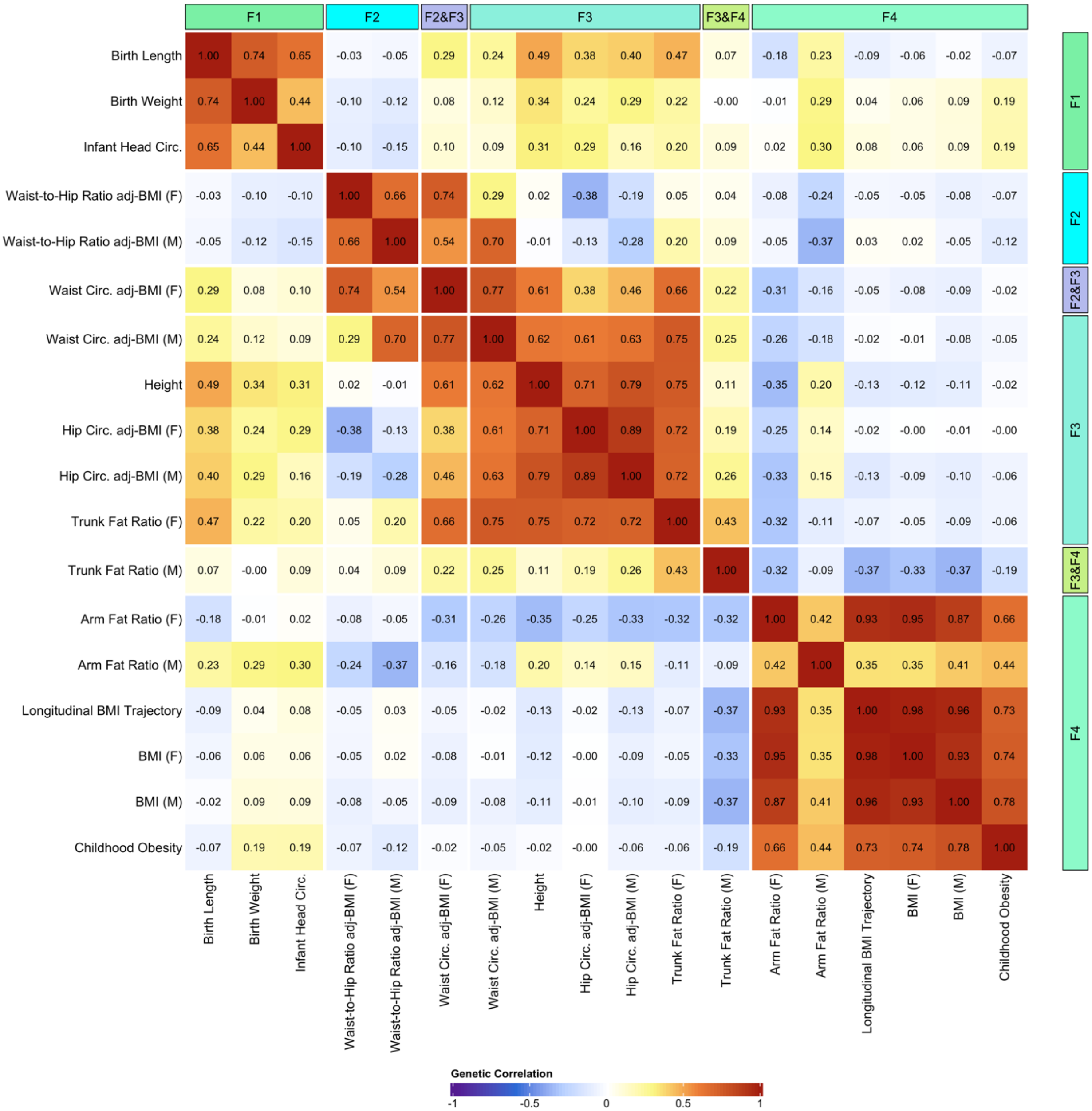


Supplementary Figure 2: The genetic correlation matrix of indicator variables included in the genomic structural equation model. Symmetrical annotations along the top and right sides of the matrix describe the specified loadings onto F1, F2, F3, and F4 for the measurement model. Among the 18 indicator variables, 6 of the included traits were stratified by sex with (M) denoting male and (F) denoting female.

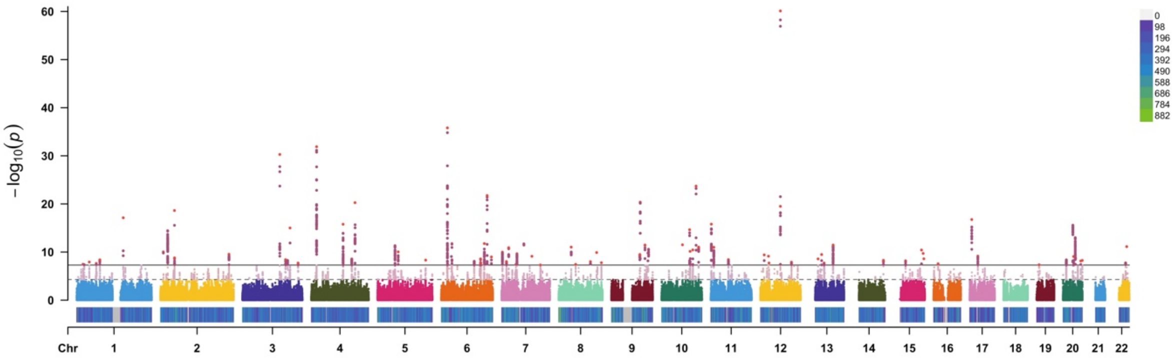

Supplementary Figure 3: GWAS Manhattan plot[11] for F1, the birth size factor, illustrating the 88 independent significant loci. The estimated effective sample size for F1 was 52,404. Each point represents a tested variant association, and SNPs above the solid line were genome-wide significant ($p < 5 \times 10^{-8}$) while SNPs above the dashed line were genome-wide suggestive ($p < 5 \times 10^{-5}$). SNP density across the genome is shown with a purple-green color scale above the x-axis and a legend in the top right.

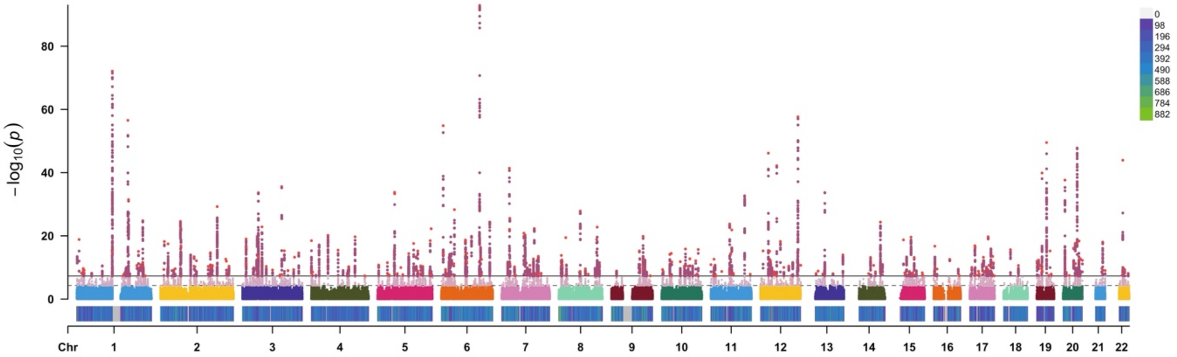


Supplementary Figure 4: GWAS Manhattan plot for F2, the abdominal size factor, illustrating the 344 independent significant loci. The estimated effective sample size for F2 was 176,820. Each point represents a tested variant association, and SNPs above the solid line were genome-wide significant ($p < 5 \times 10^{-8}$) while SNPs above the dashed line were genome-wide suggestive ($p < 5 \times 10^{-5}$). SNP density across the genome is shown with a purple-green color scale above the x-axis and a legend in the top right.

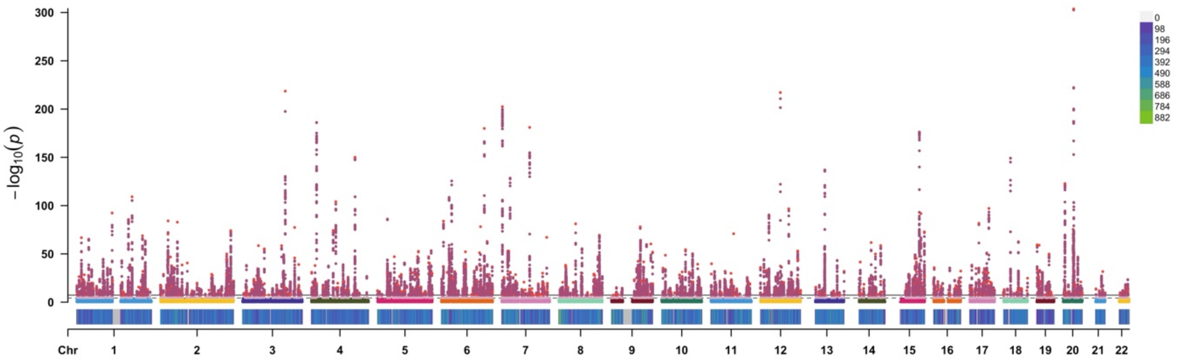


Supplementary Figure 5: GWAS Manhattan plot for F3, the body size and adipose distribution factor, illustrating the 1,173 independent significant loci. The estimated effective sample size for F3 was 690,110. Each point represents a tested variant association, and SNPs above the solid line were genome-wide significant ($p < 5 \times 10^{-8}$) while SNPs above the dashed line were genome-wide suggestive ($p < 5 \times 10^{-5}$). SNP density across the genome is shown with a purple-green color scale above the x-axis and a legend in the top right.

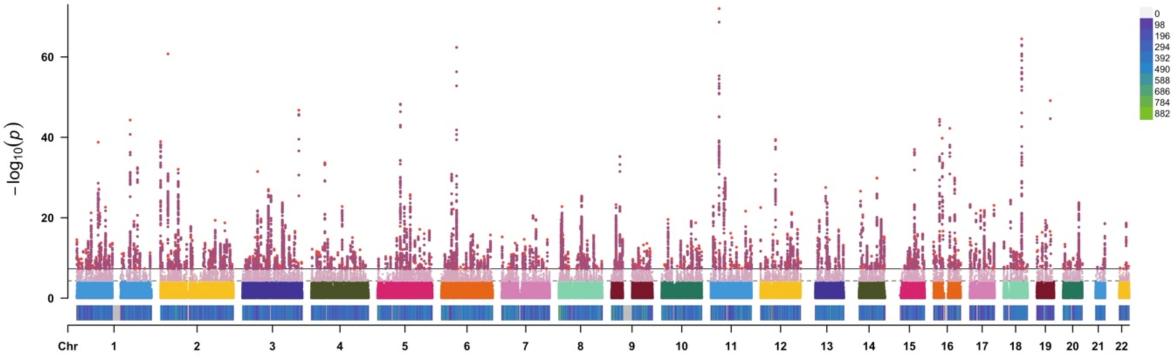


Supplementary Figure 6: GWAS Manhattan plot for F4, the adiposity factor, illustrating the 675 independent significant loci. The estimated effective sample size for F4 was 393,268. Each point represents a tested variant association, and SNPs above the solid line were genome-wide significant ($p < 5 \times 10^{-8}$) while SNPs above the dashed line were genome-wide suggestive ($p < 5 \times 10^{-5}$). SNP density across the genome is shown with a purple-green color scale above the x-axis and a legend in the top right.

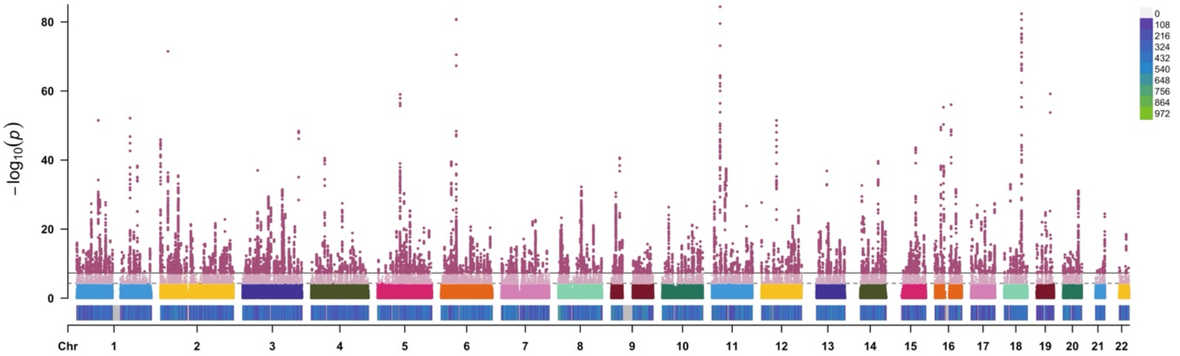


Supplementary Figure 7: Manhattan plot for the BMI GWAS illustrating the 1,035 independent significant loci. The median sample size for the BMI GWAS was 484,680 (combined males and females). Each point represents a tested variant association, and SNPs above the solid line were genome-wide significant ($p < 5 \times 10^{-8}$) while SNPs above the dashed line were genome-wide suggestive ($p < 5 \times 10^{-5}$). SNP density across the genome is shown with a purple-green color scale above the x-axis and a legend in the top right.

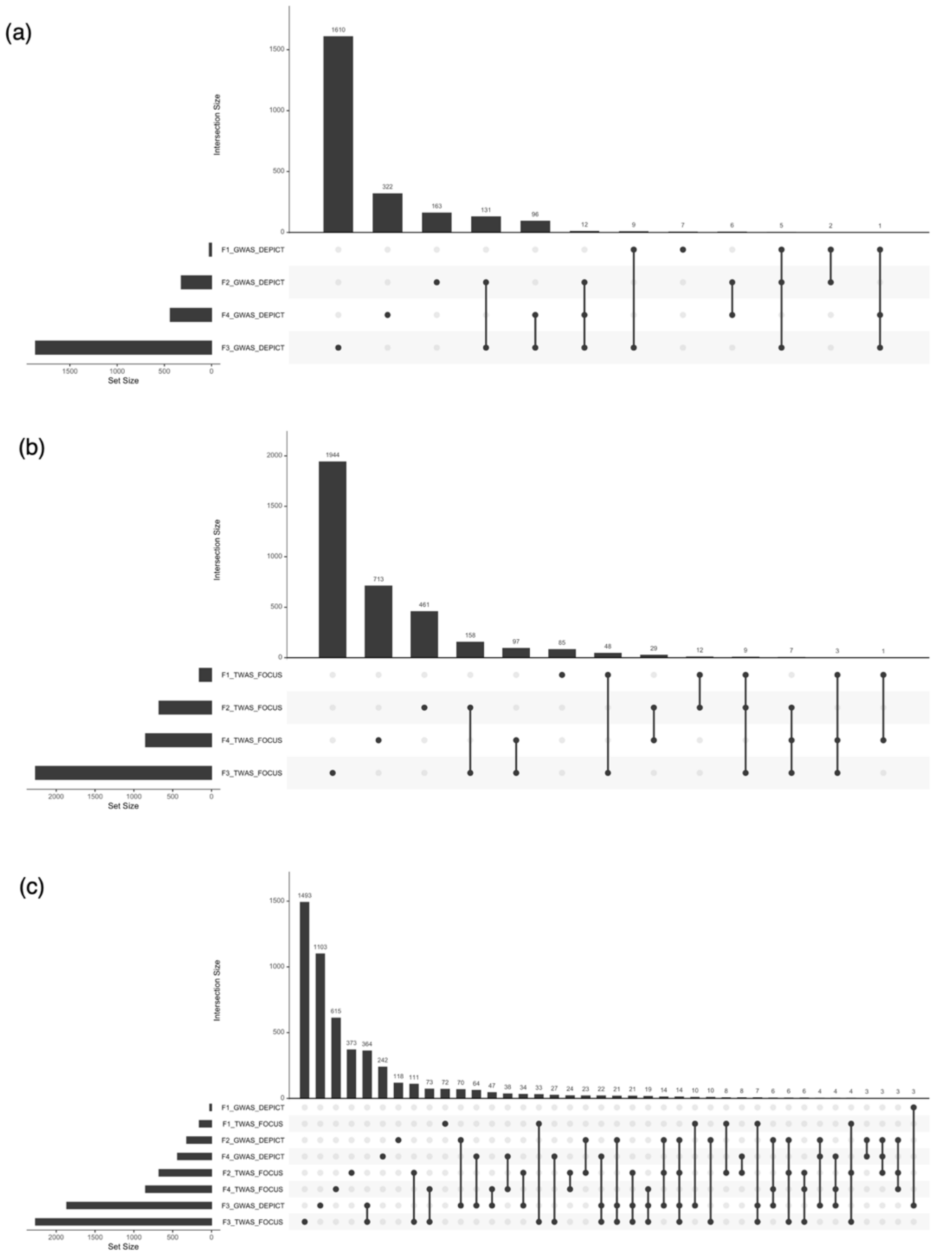


Supplementary Figure 8: UpSet plot visualization of the set membership for identified genes in the DEPICT and FOCUS analyses of F1, F2, F3, and F4. Panel (a) shows gene sets identified by DEPICT (significantly prioritized genes from DEPICT with FDR < 0.05 ), panel (b) shows gene sets identified by FOCUS (finemapped genes with PIP > 0.1 across non-null 90% credibles sets), and panel (c) shows gene sets identified by DEPICT and FOCUS. The implicated genes from DEPICT describe variant-to-gene mapping from the GWAS, and the implicated genes from FOCUS describe genes with predicted expression that are putatively causal for the phenotype in a brain-tissue-prioritized transcriptome wide association study (TWAS). Bars are ordered from left to right based on decreasing intersection size.

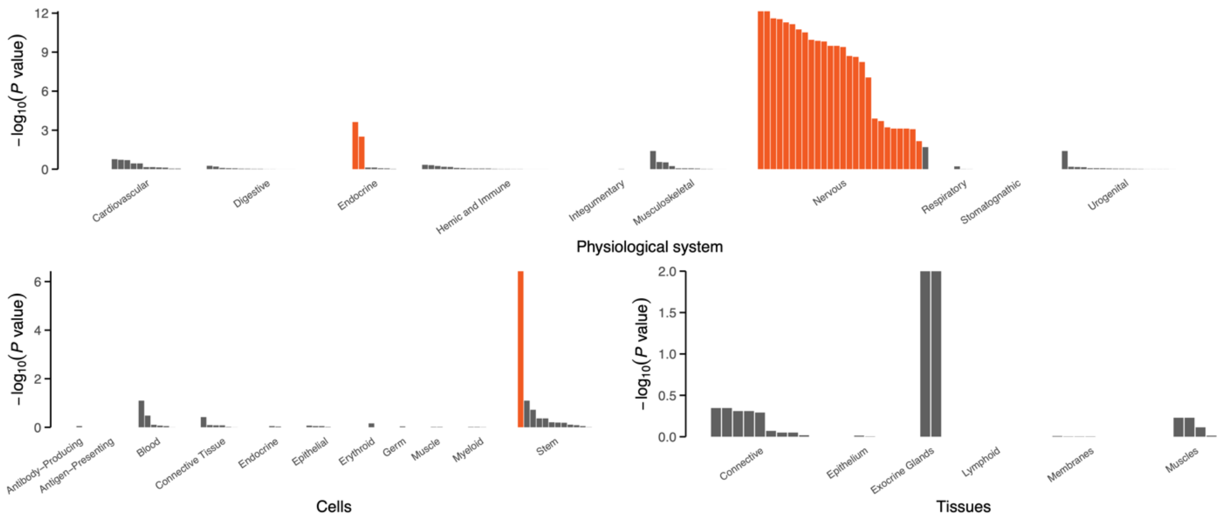


Supplementary Figure 9: The BMI DEPICT analysis showed gene expression enrichment in nervous and endocrine physiological systems (orange bars denote FDR < 0.05). Enrichment in the hypothalamus and adrenal glands were the differentiating features for BMI compared to F4 (Figure 5b).

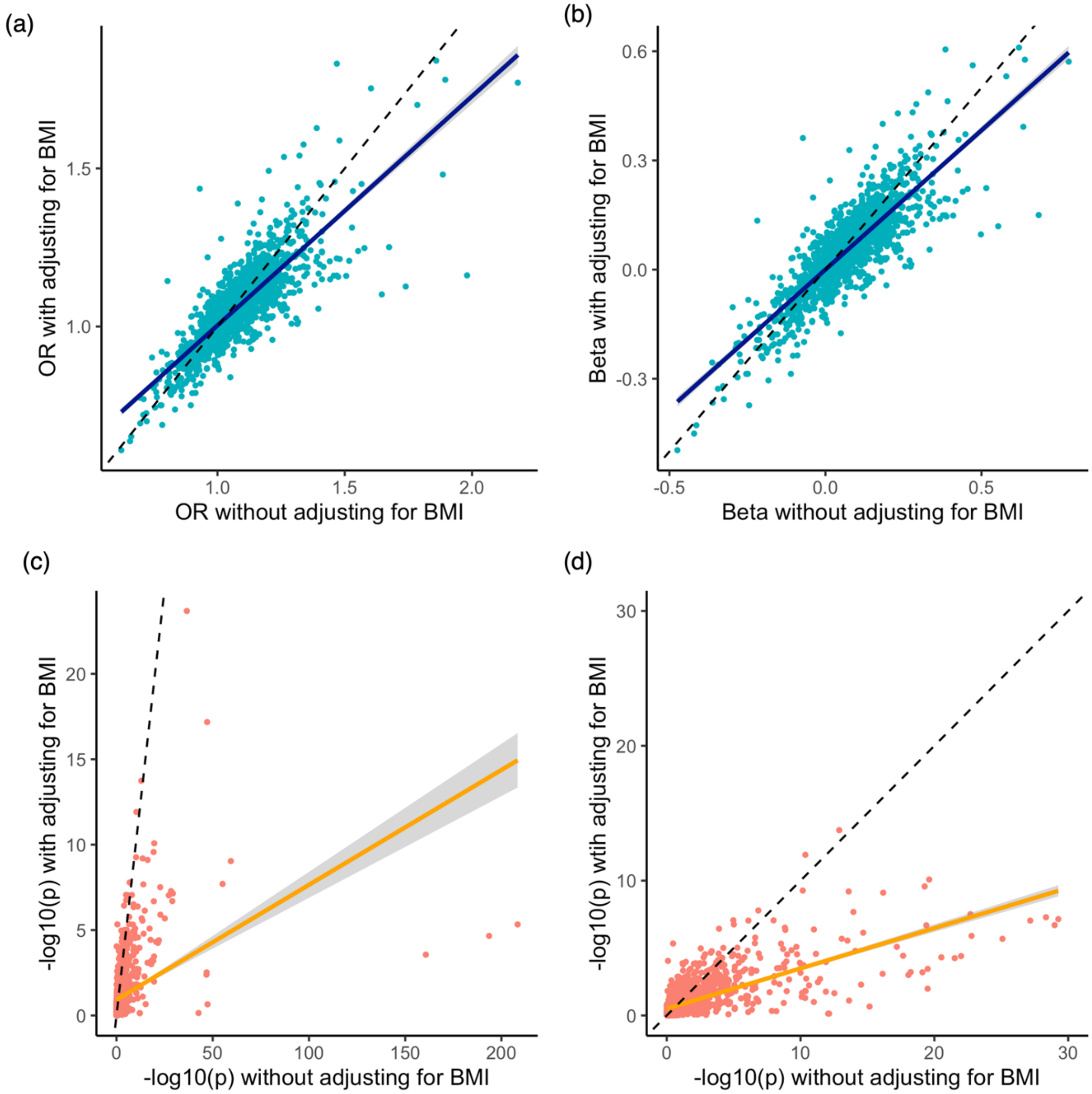


Supplementary Figure 10: A comparison of the pheWAS association model with and without adjusting for participant BMI. Attenuation of the odds ratios (a), effect sizes (b), and p-values (c-d) was generally observed after adjusting for BMI. The dotted black line in each plot illustrates a 1:1 relationship (slope of 1), and the colored lines depict the ordinary least squares regression through the data. Panel (c) shows the full range of -log10(p), and panel (d) shows the same scatter plot zoomed in (x and y axes limited from 0 to 30).

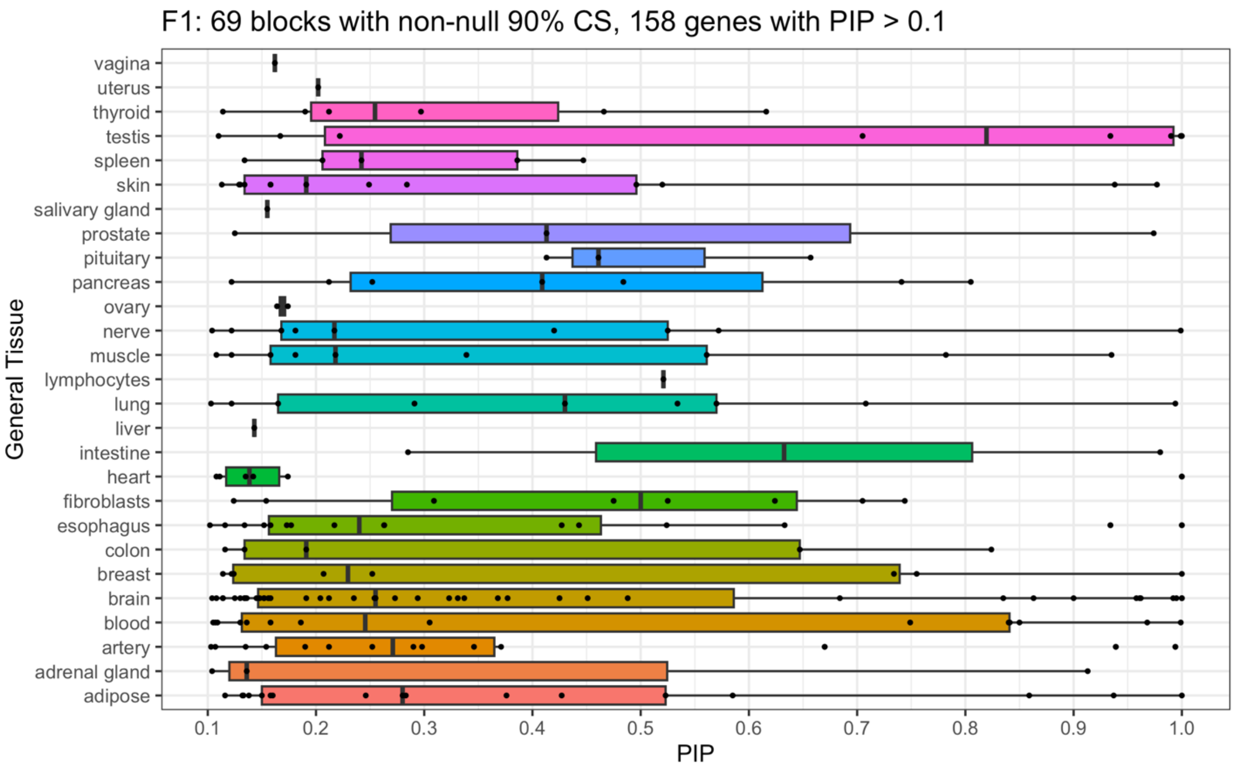


Supplementary Figure 11: The tissue-agnostic transcriptome-wide association study (TWAS) posterior inclusion probabilities (PIPs) for the F1 FOCUS analysis. Boxplots are shown for 215 genes (158 unique) across 27 tissues that were finemapped with PIP > 0.1 in 69 non-null 90% credibles sets.

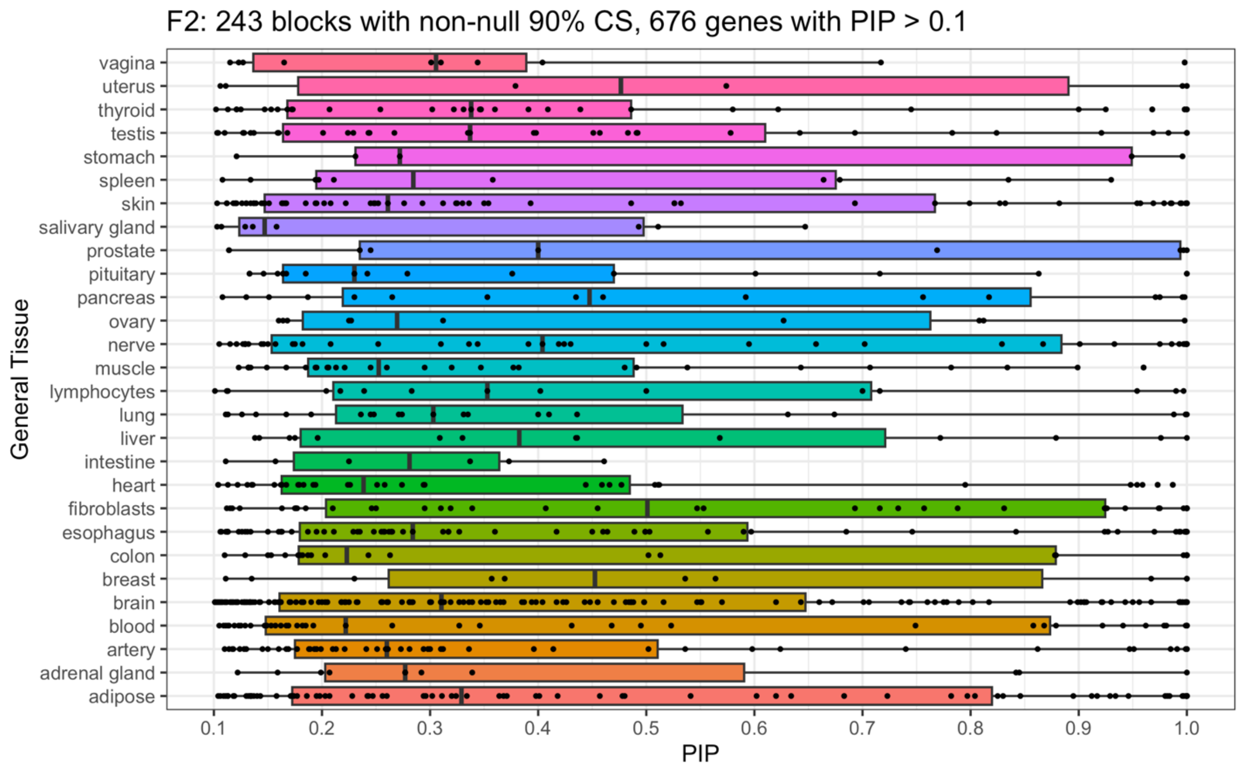


Supplementary Figure 12: The tissue-agnostic transcriptome-wide association study (TWAS) posterior inclusion probabilities (PIPs) for the F2 FOCUS analysis. Boxplots are shown for 862 genes (676 unique) across 28 tissues that were finemapped with PIP > 0.1 in 243 non-null 90% credibles sets.

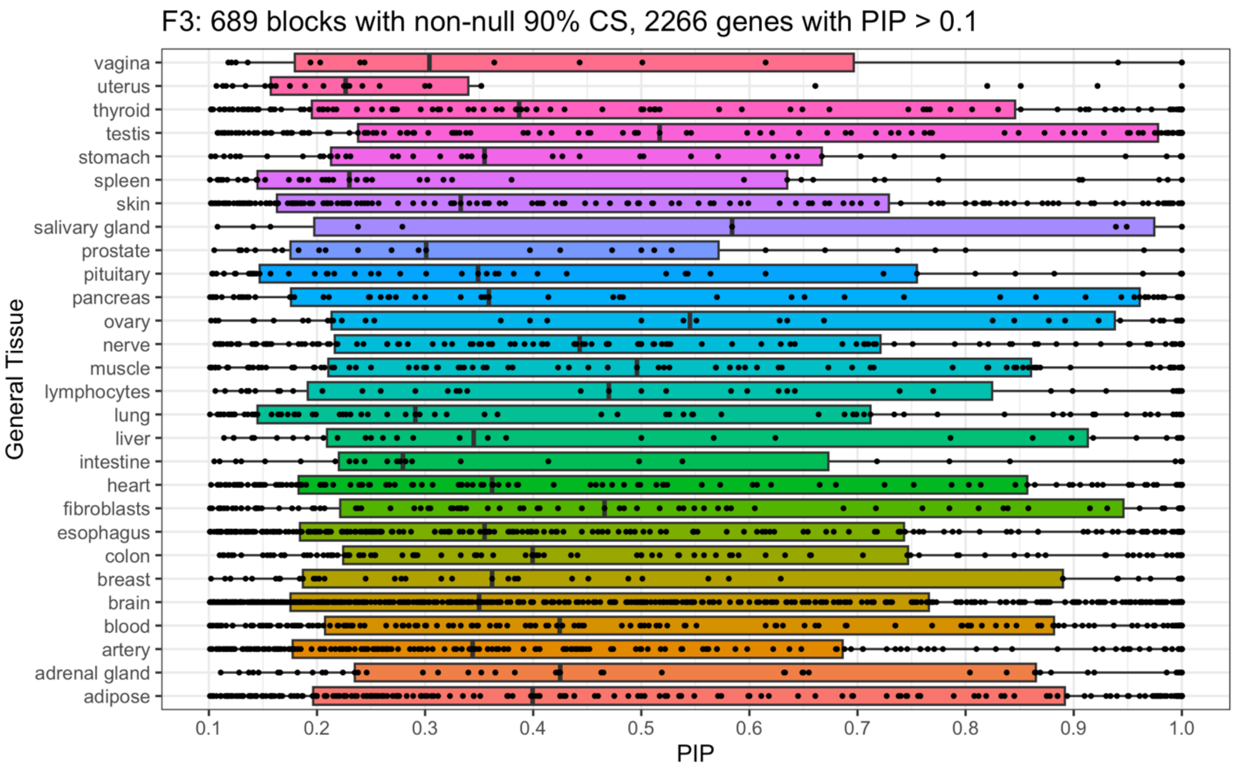


Supplementary Figure 13: The tissue-agnostic transcriptome-wide association study (TWAS) posterior inclusion probabilities (PIPs) for the F3 FOCUS analysis. Boxplots are shown for 2,944 genes (2,266 unique) across 28 tissues that were finemapped with PIP > 0.1 in 689 non-null 90% credibles sets.

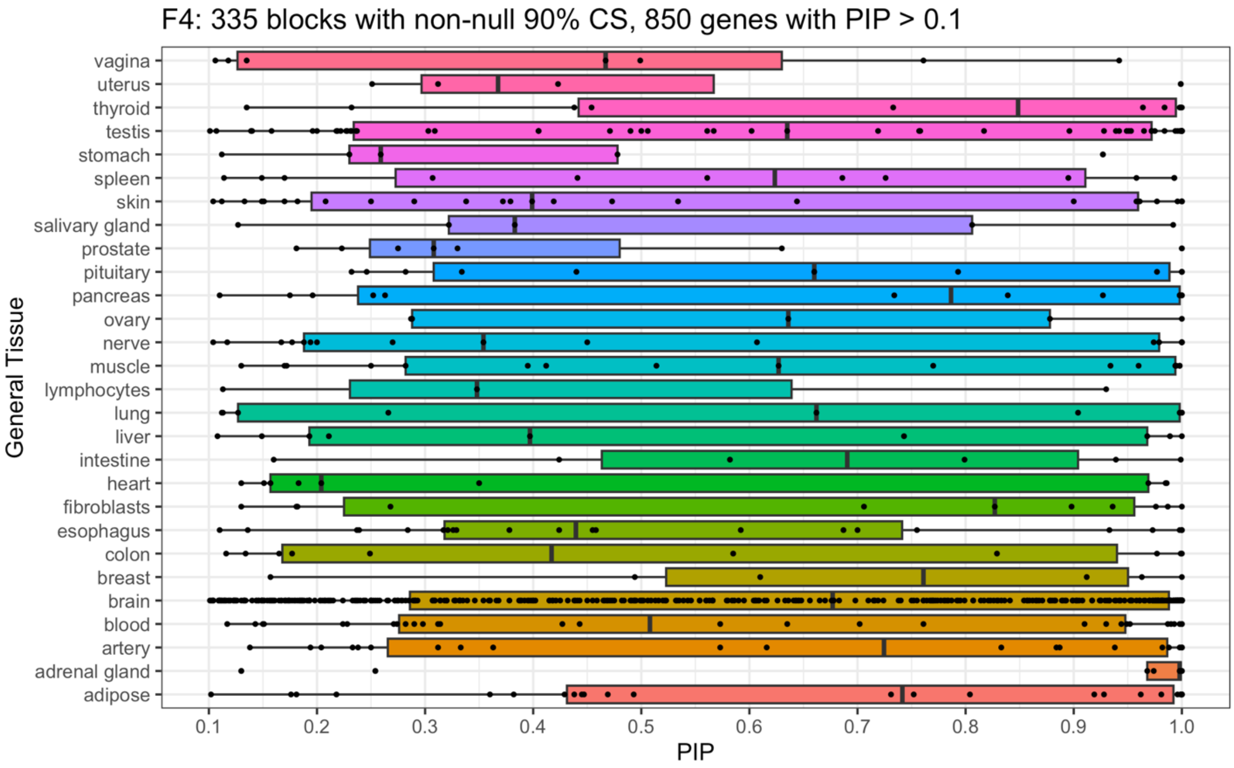


Supplementary Figure 14: The brain-tissue-prioritized transcriptome-wide association study (TWAS) posterior inclusion probabilities (PIPs) for the F4 FOCUS analysis. Boxplots are shown for 864 genes (850 unique) across 28 tissues that were finemapped with PIP > 0.1 in 335 non-null 90% credibles sets.

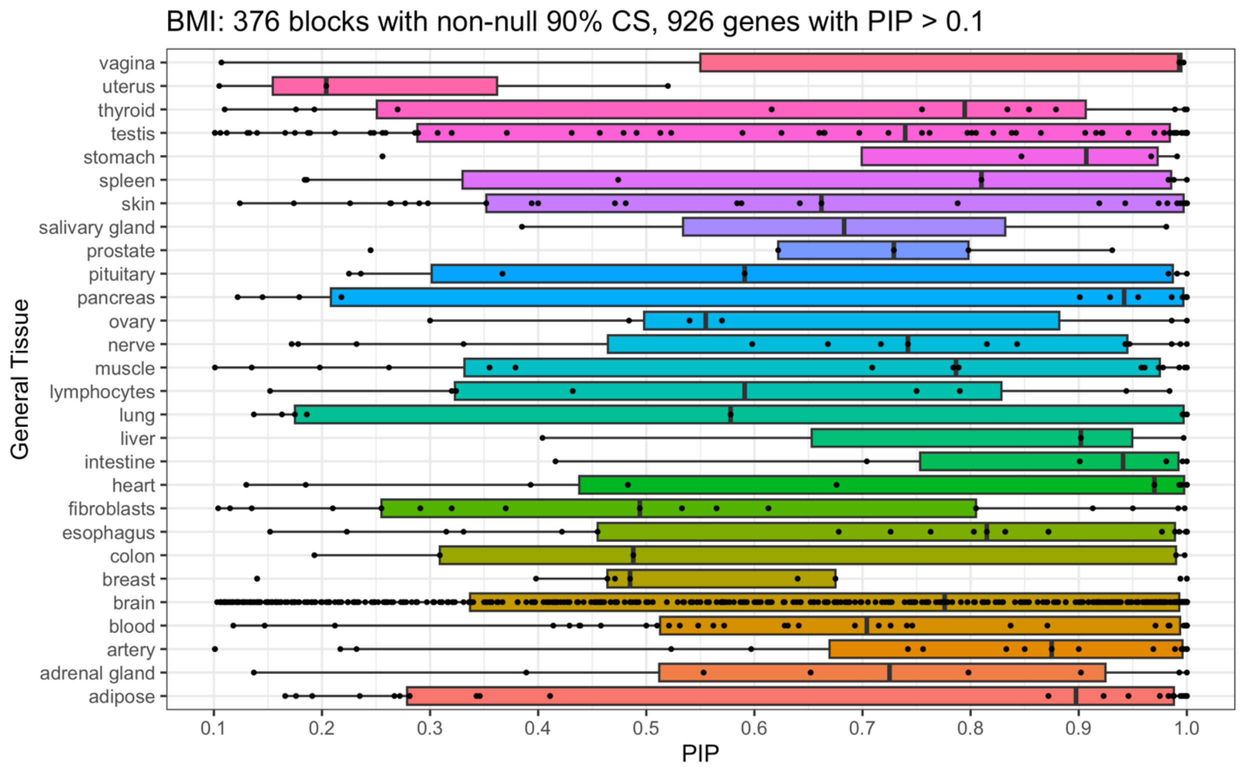


Supplementary Figure 15: The brain-tissue-prioritized transcriptome-wide association study (TWAS) posterior inclusion probabilities (PIPs) for the BMI FOCUS analysis. Boxplots are shown for 940 genes (926 unique) across 28 tissues that were finemapped with PIP > 0.1 in 376 non-null 90% credibles sets.

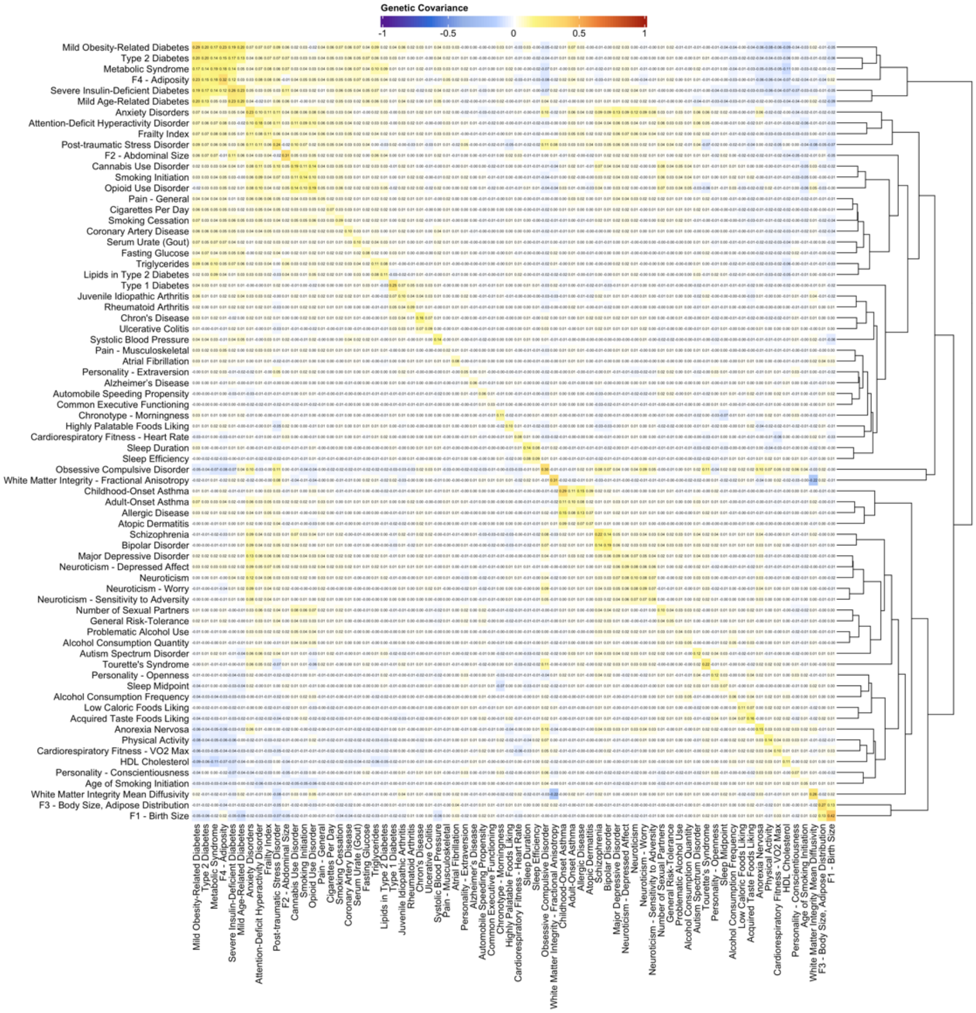


Supplementary Figure 16: The genetic covariance matrix of F1, F2, F3, F4, and 71 related traits. SNP-based heritability estimates ($h^2$) are represented along the diagonal. Rows and columns of the heatmap are ordered in correspondence to the dendrogram along the right to cluster traits with greater similarity. The full covariance matrix is shown in Supplementary Figure 51, standard errors are included in Supplementary Table 52, and information on the summary statistics is included in Supplemental Tables 3 and 48.

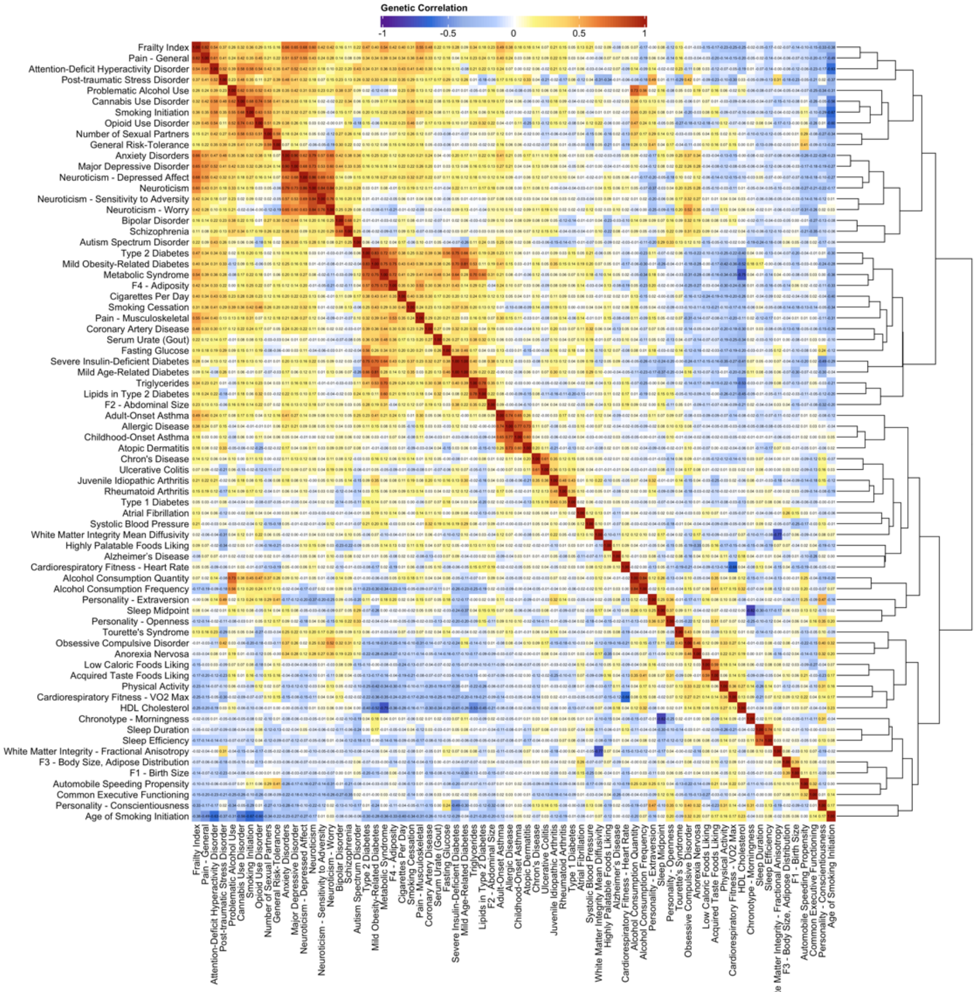


Supplementary Figure 17: The genetic correlation matrix of F1, F2, F3, F4, and 71 related traits. Rows and columns of the heatmap are ordered in correspondence to the dendrogram along the right to cluster traits with greater similarity. The full covariance matrix is shown in Supplementary Figure 49, standard errors are included in Supplementary Table 50, and information on the summary statistics is included in Supplemental Tables 3 and 48.

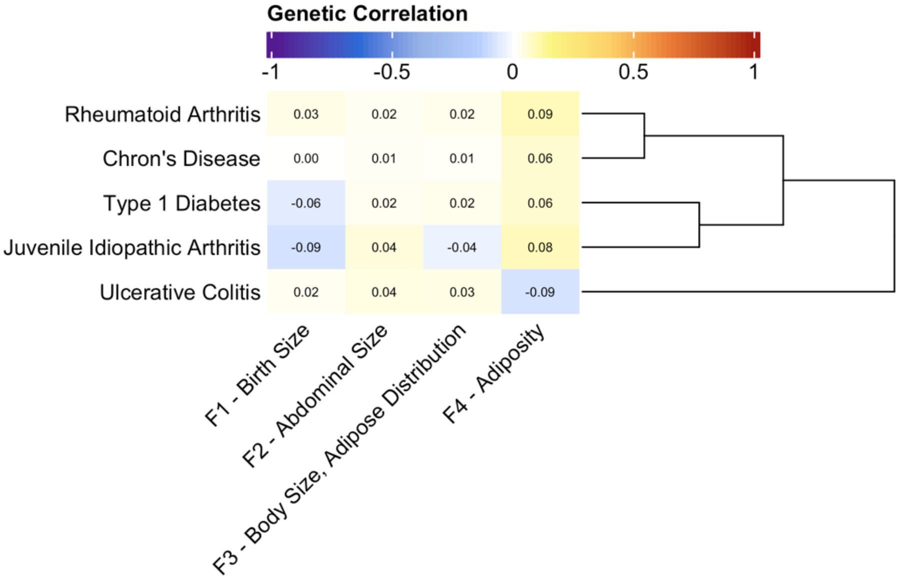


Supplementary Figure 18: The genetic correlations between F1, F2, F3, and F4 and autoimmune related traits. Rows of the heatmap are ordered in correspondence to the dendrogram along the right to cluster traits with similar relationships across the 4 factors.

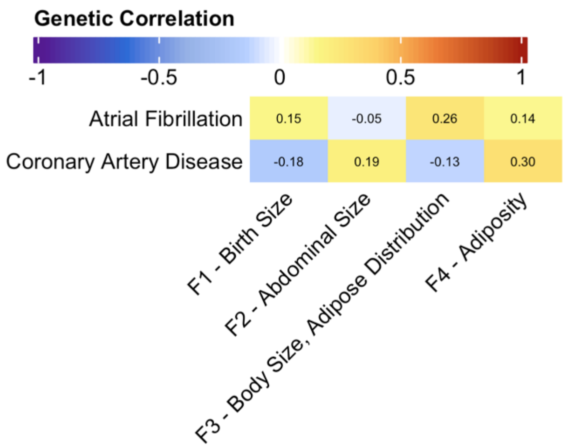


Supplementary Figure 19: The genetic correlations between F1, F2, F3, and F4 and cardiovascular related traits.

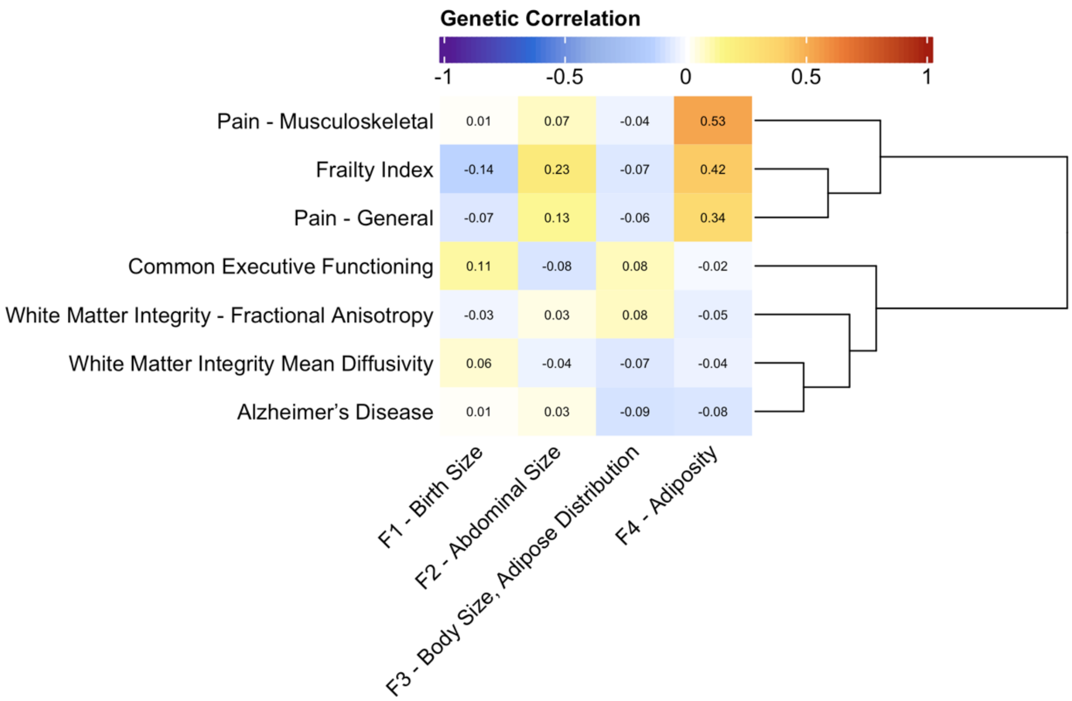


Supplementary Figure 20: The genetic correlations between F1, F2, F3, and F4 and phenotypes relating to pain, frailty, and Alzheimer's disease. Rows of the heatmap are ordered in correspondence to the dendrogram along the right to cluster traits with similar relationships across the 4 factors.

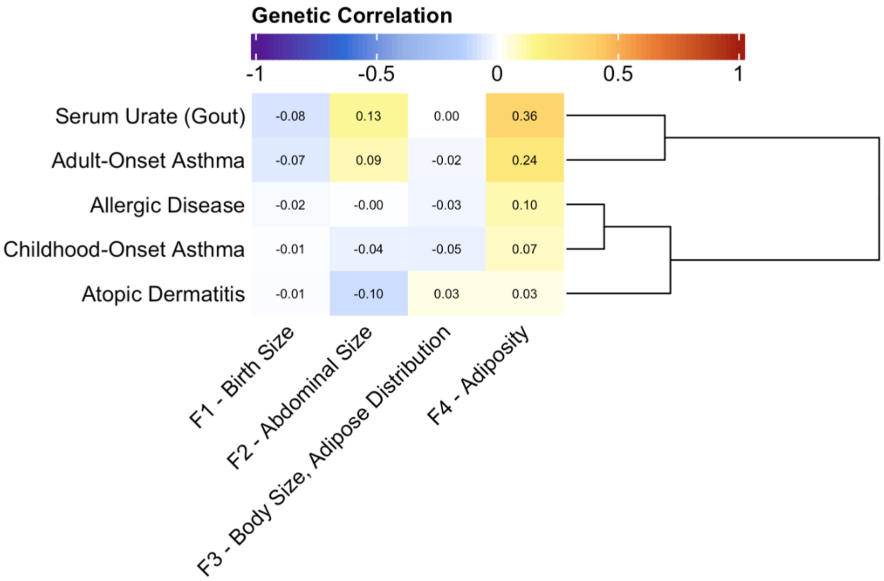


Supplementary Figure 21: The genetic correlations between F1, F2, F3, and F4 and inflammatory or allergic diseases. Rows of the heatmap are ordered in correspondence to the dendrogram along the right to cluster traits with similar relationships across the 4 factors.

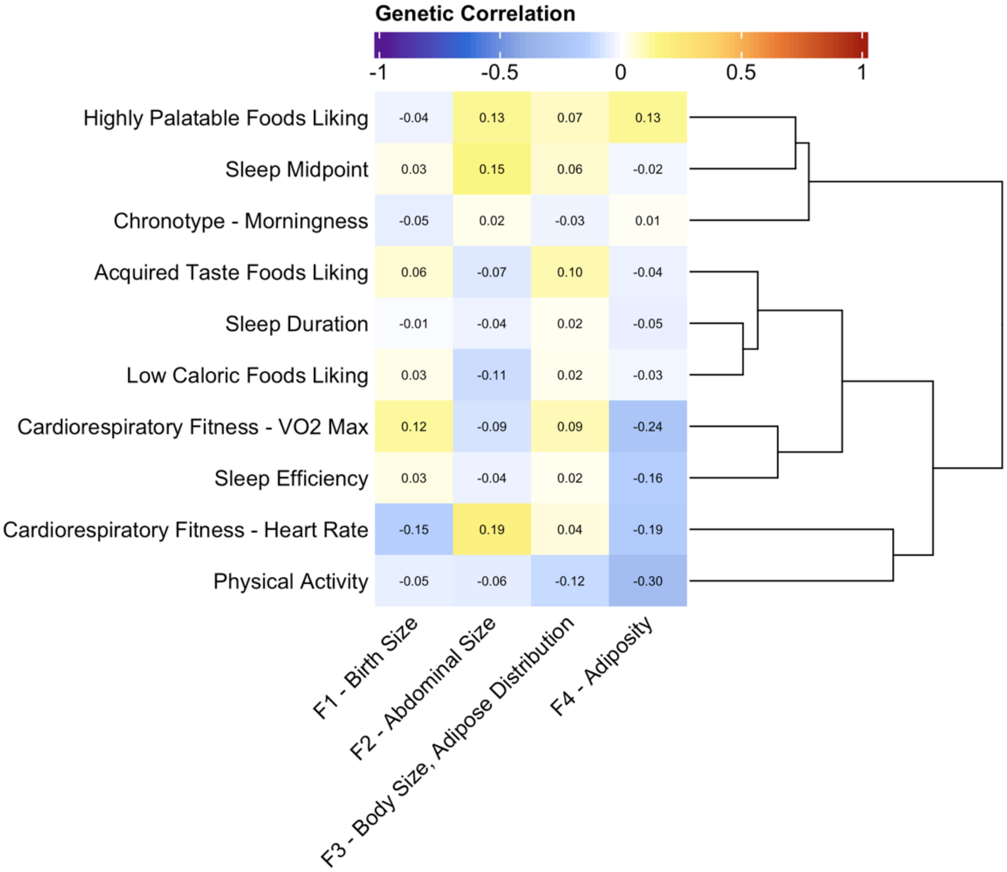


Supplementary Figure 22: The genetic correlations between F1, F2, F3, and F4 and phenotypes relating to diet, sleep, and exercise. Rows of the heatmap are ordered in correspondence to the dendrogram along the right to cluster traits with similar relationships across the 4 factors.

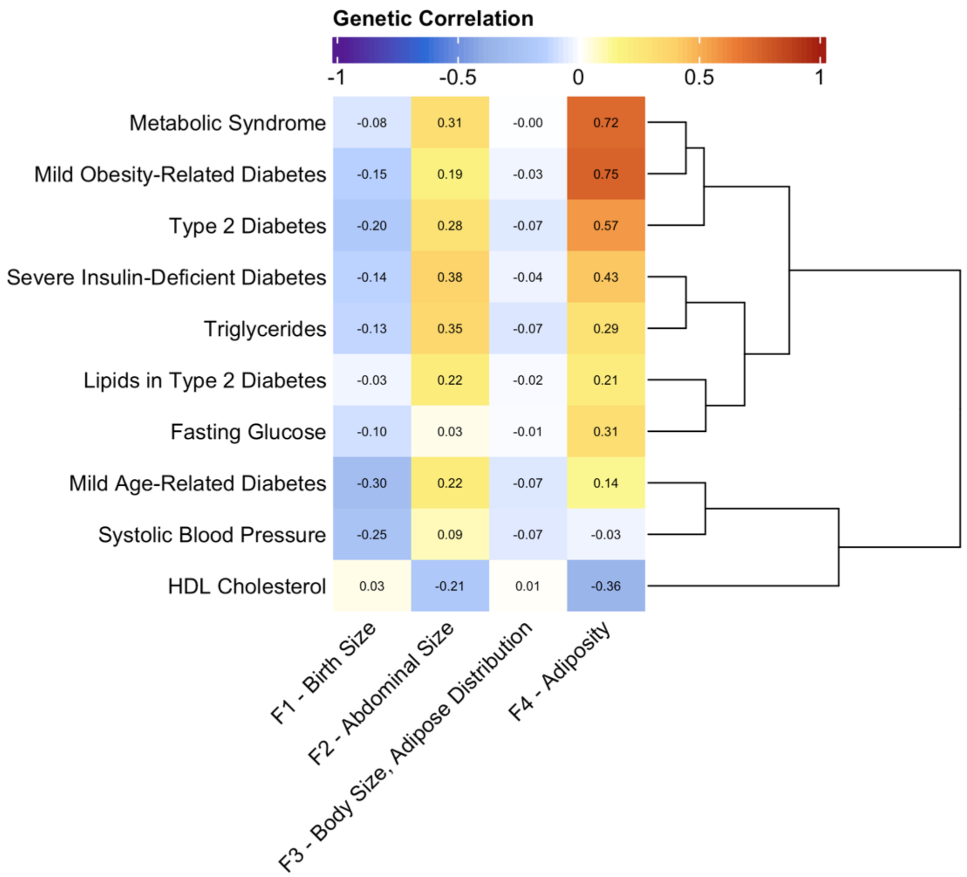


Supplementary Figure 23: The genetic correlations between F1, F2, F3, and F4 and metabolic traits. Rows of the heatmap are ordered in correspondence to the dendrogram along the right to cluster traits with similar relationships across the 4 factors.

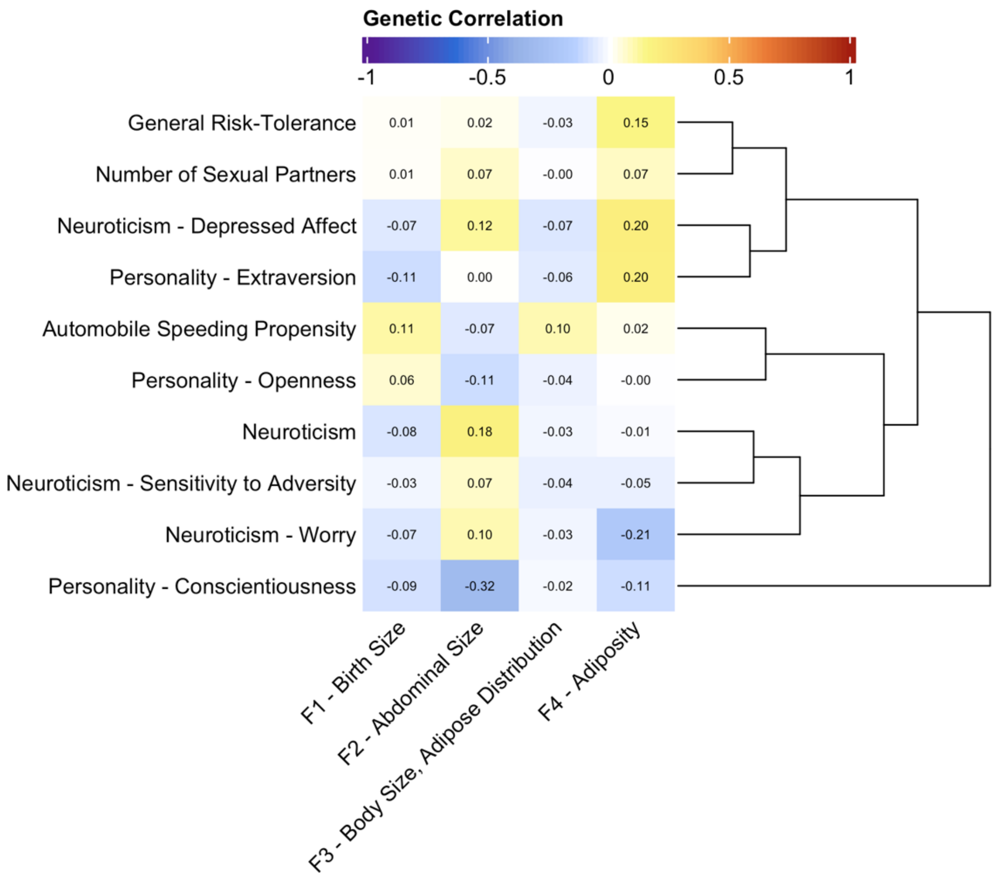


Supplementary Figure 24: The genetic correlations between F1, F2, F3, and F4 and phenotypes relating to risk tolerance, neuroticism, and personality. Rows of the heatmap are ordered in correspondence to the dendrogram along the right to cluster traits with similar relationships across the 4 factors.

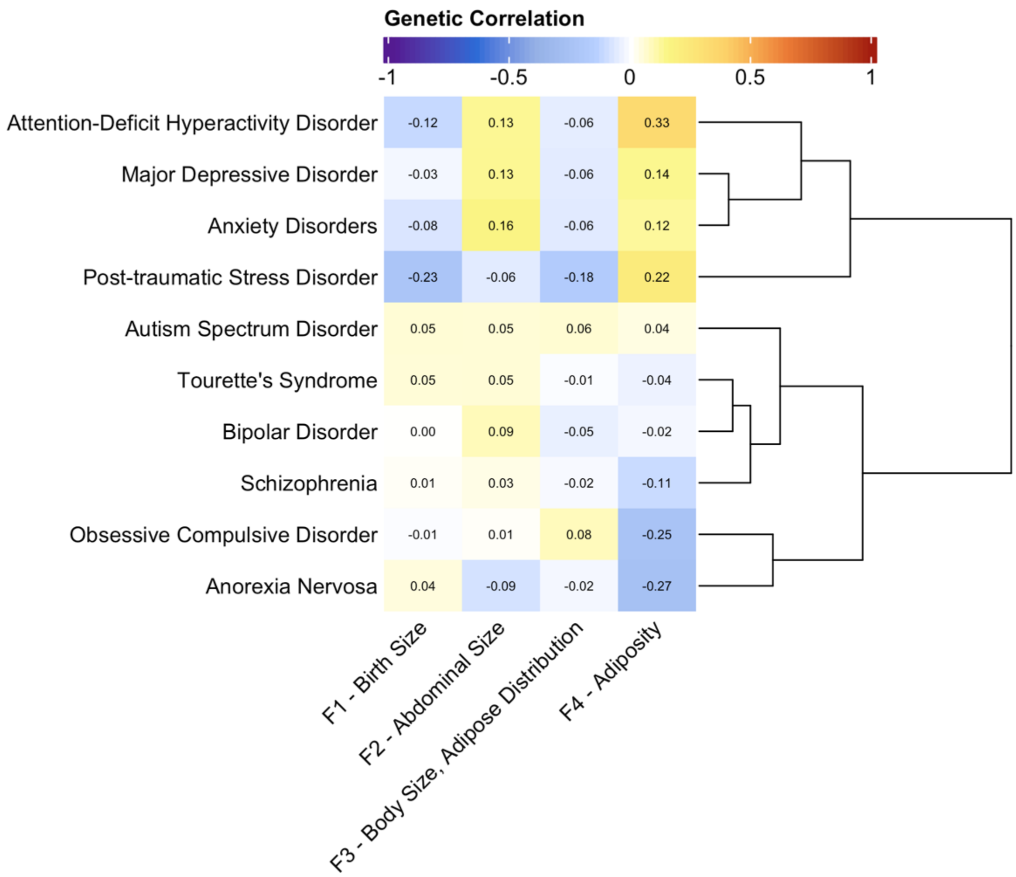


Supplementary Figure 25: The genetic correlations between F1, F2, F3, and F4 and psychopathology phenotypes. Rows of the heatmap are ordered in correspondence to the dendrogram along the right to cluster traits with similar relationships across the 4 factors.

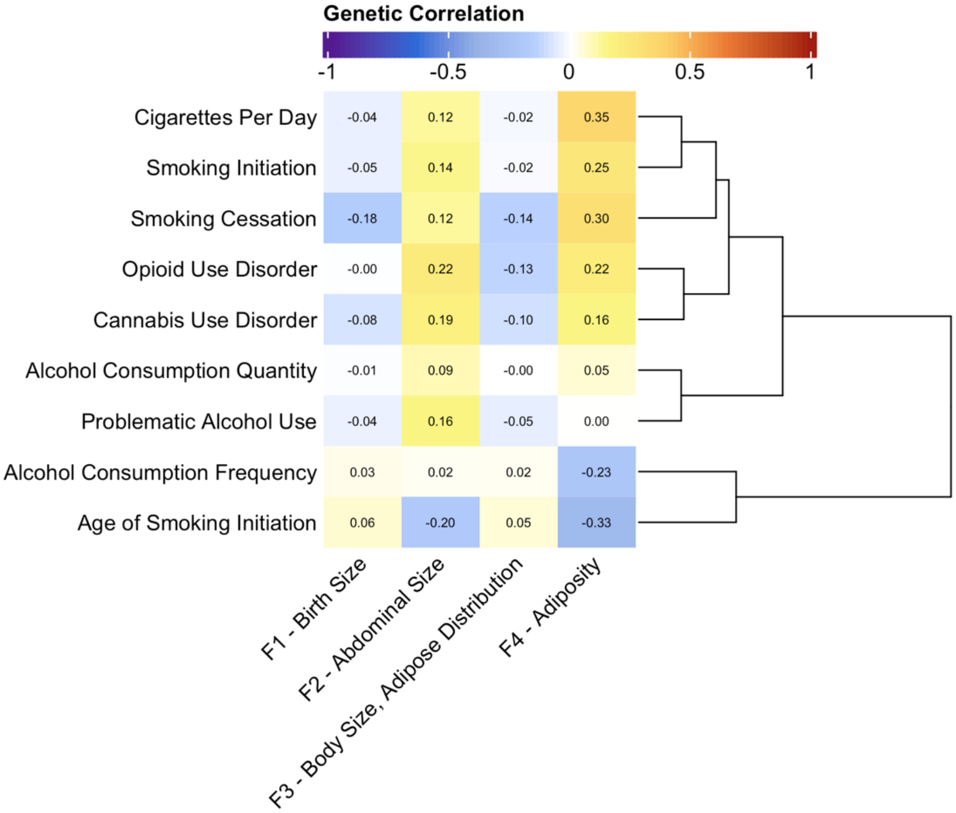


Supplementary Figure 26: The genetic correlations between F1, F2, F3, and F4 and phenotypes relating substance use. Rows of the heatmap are ordered in correspondence to the dendrogram along the right to cluster traits with similar relationships across the 4 factors.

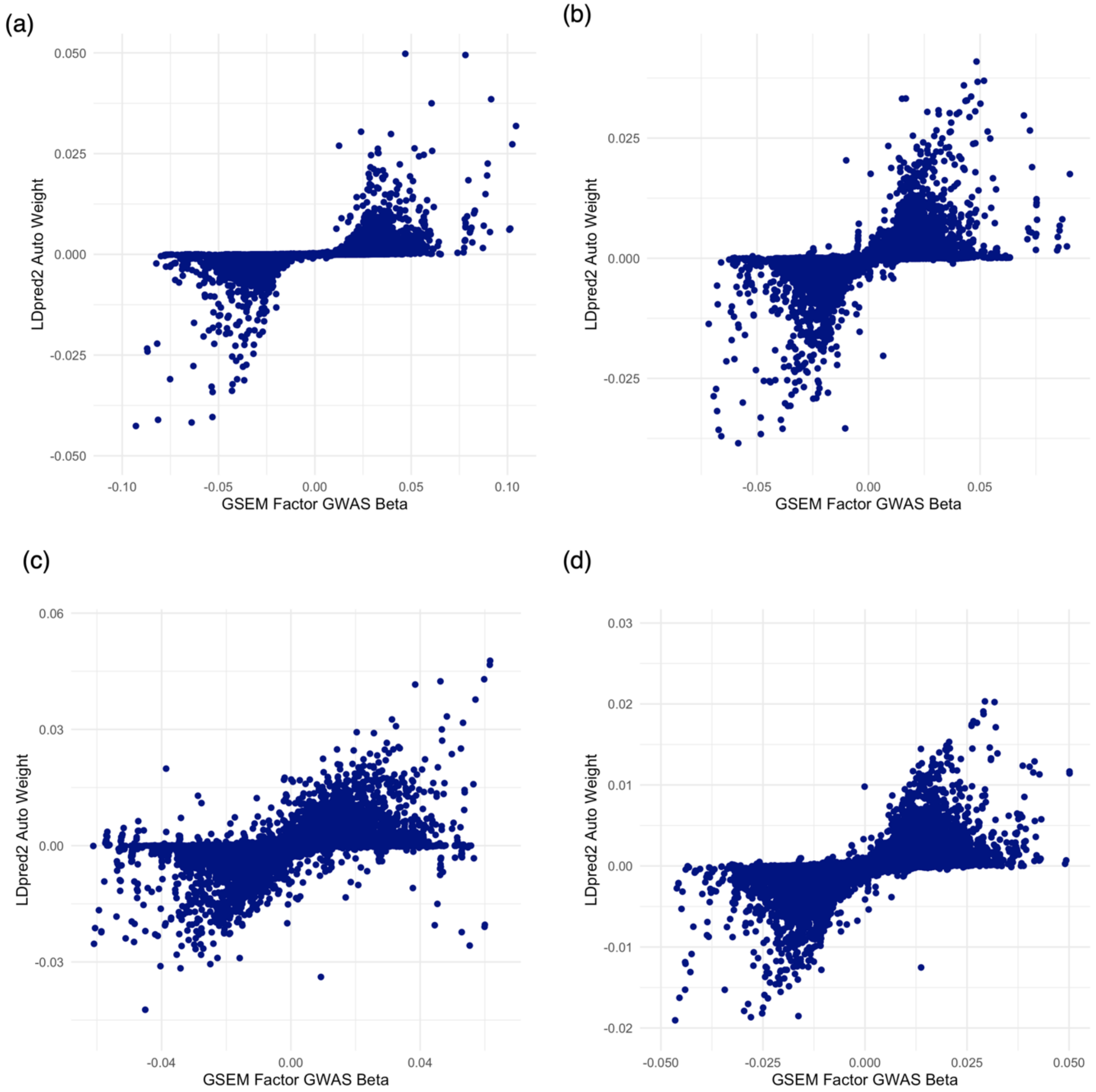


Supplementary Figure 27: The LDpred2-auto[12] estimated posterior mean causal effect sizes (y-axis) versus the raw GWAS effect sizes (x-axis) are shown for F1 (a), F2 (b), F3 (c), and F4 (d). The PRS weights tended to maintain consistent effect direction relative to the GWAS betas but showed attenuation due to linkage disequilibrium structure.

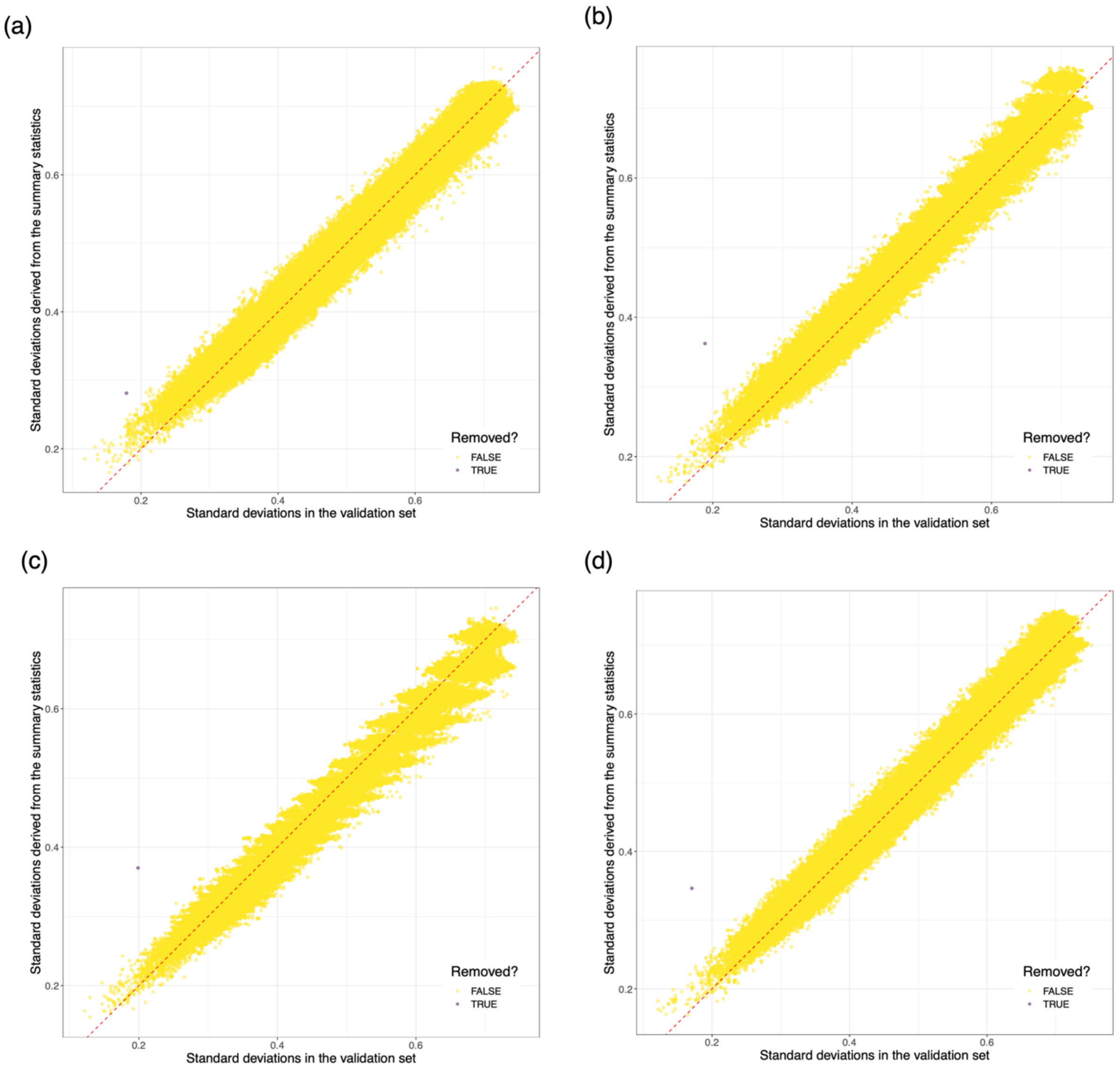


Supplementary Figure 28: Panels (a), (b), (c), and (d) illustrate the polygenic risk scoring (PRS) single nucleotide polymorphism (SNP) standard deviation quality control (QC) step from LDpred2[12] for F1, F2, F3, and F4, respectively. The European ancestry UK Biobank LD reference panel (N = 5,000; x-axis) had matching SNP variation compared to the European ancestry based multivariate GWASs (y-axis), and most SNPs fell diagonally along the dashed red line. Nearly all SNPs were retained in the PRS training datasets (shaded yellow) and only a few were removed (shaded purple).

Supplementary Table 3: GWAS summary statistics citations and sample sizes. These GWASs were used to evaluate genetic correlations with each of the 4 factors using LDSC. Additional information on these summary statistics is included in Supplementary Table 48.

| Trait and Reference | Abbreviated Name | N Cases | N Controls | N Total |
|---|---|---|---|---|
| Anorexia Nervosa[13] | AN | 16992 | 55525 | 72517 |
| Anxiety Disorders[14] | ANX | 31977 | 82114 | 114091 |
| Attention-Deficit Hyperactivity Disorder[15] | ADHD | 38691 | 186843 | 225534 |
| Autism Spectrum Disorder[16] | ASD | 18381 | 27969 | 46350 |
| Bipolar Disorder[17] | BIP | 41917 | 371549 | 413466 |
| Cannabis Use Disorder[18] | CUD | 14808 | 343726 | 358534 |
| Major Depressive Disorder[19] | MDD | 170756 | 329443 | 500199 |
| Obsessive Compulsive Disorder[20] | OCD | 2688 | 7037 | 9725 |
| Opioid Use Disorder[21] | OUD | 3211 | 11589 | 14800 |
| Post-traumatic Stress Disorder[22] | PTSD | 2424 | 7113 | 9537 |
| Schizophrenia[23] | SCZ | 53386 | 77258 | 130644 |
| Tourette's Syndrome[24] | TS | 4819 | 9488 | 14307 |
| Childhood-Onset Asthma[25] | AsCO | 9433 | 318237 | 327670 |
| Adult-Onset Asthma[25] | AsAO | 21564 | 318237 | 339801 |
| Allergic Disease[26] | Allerg | 96794 | 145775 | 242569 |
| Atopic Dermatitis[27] | ECZ | 22608 | 393165 | 415773 |
| Chronotype - Morningness[28] | Chrono | NA | NA | 449734 |
| Sleep Midpoint[29] | SleepM | NA | NA | 84810 |
| Sleep Duration[29] | SleepD | NA | NA | 85449 |
| Sleep Efficiency[29] | SleepE | NA | NA | 84810 |
| Metabolic Syndrome[30] | MetSyn | 59677 | 231430 | 291107 |
| Fasting Glucose[31] | FastGluc | NA | NA | 46186 |
| HDL Cholesterol[32] | HDL | NA | NA | 99900 |
| Triglycerides[32] | Triglyc | NA | NA | 96598 |
| Systolic Blood Pressure[33] | SBP | NA | NA | 745787 |
| Atrial Fibrillation[34] | AtrFib | 55114 | 482295 | 537409 |
| Coronary Artery Disease[35] | CAD | 181522 | 984168 | 1165690 |
| Type 2 Diabetes[36] | T2D | 62892 | 596424 | 659316 |
| Severe Autoimmune Diabetes[37] | T2DSAID | 450 | 2744 | 3194 |

| | | | | |
|---|---|---|---|---|
| Severe Insulin-Deficient Diabetes[37] | T2DSIDD | 1186 | 2744 | 3930 |
| Severe Insulin-Resistant Diabetes[37] | T2DSIRD | 1125 | 2744 | 3869 |
| Mild Obesity-Related Diabetes[37] | T2DMOD | 1372 | 2744 | 4116 |
| Mild Age-Related Diabetes[37] | T2DMARD | 2853 | 2744 | 5597 |
| Lipids in Type 2 Diabetes[38] | LipT2D | NA | NA | 21176 |
| Common Executive Functioning[39] | CEF | NA | NA | 1564847 |
| Problematic Alcohol Use[40] | PAU | NA | NA | 539828 |
| Alcohol Consumption Quantity[40] | ACQ | NA | NA | 543787 |
| Alcohol Consumption Frequency[40] | ACF | NA | NA | 445688 |
| Cigarettes Per Day[41] | CPD | NA | NA | 326497 |
| Smoking Initiation[41] | SI | 393680 | 411751 | 805431 |
| Smoking Cessation[41] | SC | 266596 | 121717 | 388313 |
| Age of Smoking Initiation[41] | AgeSI | NA | NA | 323386 |
| General Risk-Tolerance[42] | Risk | NA | NA | 466571 |
| Automobile Speeding Propensity[42] | Speed | NA | NA | 404291 |
| Number of Sexual Partners[42] | Nsex | NA | NA | 370711 |
| Neuroticism[43] | Neur | NA | NA | 168105 |
| Neuroticism - Depressed Affect[44] | NeurD | NA | NA | 357957 |
| Neuroticism - Worry[44] | NeurW | NA | NA | 348219 |
| Neuroticism - Sensitivity to Adversity[45] | NeurS | NA | NA | 449484 |
| Low Caloric Foods Liking[46] | FdLC | NA | NA | 154998 |
| Acquired Taste Foods Liking[46] | FdA | NA | NA | 156639 |
| Highly Palatable Foods Liking[46] | FdHP | NA | NA | 158666 |
| White Matter Integrity - Fractional Anisotropy[47] | FAwmi | NA | NA | 17663 |
| White Matter Integrity Mean Diffusivity[47] | MDwmi | NA | NA | 17467 |

| | | | | |
|---|---|---|---|---|
| Frailty Index[48] | Frail | NA | NA | 175226 |
| Personality - Extraversion[49] | Extrv | NA | NA | 63030 |
| Personality - Agreeableness[50] | Agr | NA | NA | 17375 |
| Personality - Conscientiousness[50] | Con | NA | NA | 17375 |
| Personality - Openness[50] | Open | NA | NA | 17375 |
| Alzheimer's Disease[51] | ALZ | 21982 | 41944 | 63926 |
| Pain - General[52] | PainG | NA | NA | 418910 |
| Pain - Musculoskeletal[52] | PainM | NA | NA | 464748 |
| Type 1 Diabetes[53] | T1D | 18942 | 501638 | 520580 |
| Rheumatoid Arthritis | RA | 14361 | 43923 | 58284 |
| Juvenile Idiopathic Arthritis[54] | JIA | 3305 | 9196 | 12501 |
| Ulcerative Colitis[55] | UC | 12366 | 33609 | 45975 |
| Chron's Disease[55] | CD | 12194 | 28072 | 40266 |
| Serum Urate (Gout)[56] | Urate | NA | NA | 288666 |
| F1 - Birth Size | F1 | NA | NA | 52404 |
| F2 - Abdominal Size | F2 | NA | NA | 176820 |
| F3 - Body Size, Adipose Distribution | F3 | NA | NA | 690110 |
| F4 - Adiposity | F4 | NA | NA | 393268 |
| Physical Activity[57] | PhysA | NA | NA | 89683 |
| Cardiorespiratory Fitness - VO2 Max[57] | FitVO2 | NA | NA | 70783 |
| Cardiorespiratory Fitness - Heart Rate[57] | FitHR | NA | NA | 70783 |

References


1. Bradfield, J. P. *et al.* A trans-ancestral meta-analysis of genome-wide association studies reveals loci associated with childhood obesity. *Hum. Mol. Genet.* **28**, 3327–3338 (2019).
2. Vogelezang, S. *et al.* Genetics of early-life head circumference and genetic correlations with neurological, psychiatric and cognitive outcomes. *BMC Med. Genomics* **15**, 124 (2022).
3. van der Valk, R. J. P. *et al.* A novel common variant in DCST2 is associated with length in early life and height in adulthood. *Hum. Mol. Genet.* **24**, 1155–1168 (2015).
4. EGG Consortium *et al.* Maternal and fetal genetic effects on birth weight and their relevance to cardio-metabolic risk factors. *Nat. Genet.* **51**, 804–814 (2019).
5. Pulit, S. L. *et al.* Meta-analysis of genome-wide association studies for body fat distribution in 694 649 individuals of European ancestry. *Hum. Mol. Genet.* **28**, 166–174 (2019).
6. The ADIPOGen Consortium *et al.* New genetic loci link adipose and insulin biology to body fat distribution. *Nature* **518**, 187–196 (2015).
7. Yengo, L. *et al.* A saturated map of common genetic variants associated with human height. *Nature* **610**, 704–712 (2022).
8. The Electronic Medical Records and Genomics (eMERGE) Consortium *et al.* Defining the role of common variation in the genomic and biological architecture of adult human height. *Nat. Genet.* **46**, 1173–1186 (2014).
9. Ko, S. *et al.* GWAS of longitudinal trajectories at biobank scale. *Am. J. Hum. Genet.* **109**, 433–445 (2022).
10. Rask-Andersen, M., Karlsson, T., Ek, W. E. & Johansson, Å. Genome-wide association study of body fat distribution identifies adiposity loci and sex-specific genetic effects. *Nat. Commun.* **10**, 339 (2019).
11. Yin, L. *et al.* rMVP: A Memory-efficient, Visualization-enhanced, and Parallel-accelerated Tool for Genome-wide Association Study. *Genomics Proteomics Bioinformatics* **19**, 619–628 (2021).
12. LDpred2: better, faster, stronger | Bioinformatics | Oxford Academic. https://academic.oup.com/bioinformatics/article/36/22-23/5424/6039173.
13. Watson, H. J. *et al.* Genome-wide association study identifies eight risk loci and implicates metabo-psychiatric origins for anorexia nervosa. *Nat. Genet.* **51**, 1207–1214 (2019).
14. Purves, K. L. *et al.* A major role for common genetic variation in anxiety disorders. *Mol. Psychiatry* **25**, 3292–3303 (2020).
15. Demontis, D. *et al.* Genome-wide analyses of ADHD identify 27 risk loci, refine the genetic architecture and implicate several cognitive domains. *Nat. Genet.* **55**, 198–208 (2023).
16. Anney, R. J. L. *et al.* Meta-analysis of GWAS of over 16,000 individuals with autism spectrum disorder highlights a novel locus at 10q24.32 and a significant overlap with schizophrenia. *Mol. Autism* **8**, 21 (2017).
17. Mullins, N. *et al.* Genome-wide association study of more than 40,000 bipolar disorder cases provides new insights into the underlying biology. *Nat. Genet.* **53**, 817–829 (2021).
18. Johnson, E. C. *et al.* A large-scale genome-wide association study meta-analysis of cannabis use disorder. *Lancet Psychiatry* **7**, 1032–1045 (2020).
19. Howard, D. M. *et al.* Genome-wide meta-analysis of depression identifies 102 independent variants and highlights the importance of the prefrontal brain regions. *Nat. Neurosci.* **22**, 343–352 (2019).
20. Arnold, P. D. *et al.* Revealing the complex genetic architecture of obsessive–compulsive disorder using meta-analysis. *Mol. Psychiatry* **23**, 1181–1188 (2018).

21. Polimanti, R. *et al.* Leveraging genome-wide data to investigate differences between opioid use vs. opioid dependence in 41,176 individuals from the Psychiatric Genomics Consortium. *Mol. Psychiatry* **25**, 1673–1687 (2020).
22. Duncan, L. E. *et al.* Largest GWAS of PTSD (N=20 070) yields genetic overlap with schizophrenia and sex differences in heritability. *Mol. Psychiatry* **23**, 666–673 (2018).
23. Trubetskoy, V. *et al.* Mapping genomic loci implicates genes and synaptic biology in schizophrenia. *Nature* **604**, 502–508 (2022).
24. Yu, D. *et al.* Interrogating the Genetic Determinants of Tourette's Syndrome and Other Tic Disorders Through Genome-Wide Association Studies. *Am. J. Psychiatry* **176**, 217–227 (2019).
25. Pividori, M., Schoettler, N., Nicolae, D. L., Ober, C. & Im, H. K. Shared and distinct genetic risk factors for childhood-onset and adult-onset asthma: genome-wide and transcriptome-wide studies. *Lancet Respir. Med.* **7**, 509–522 (2019).
26. Ferreira, M. A. *et al.* Shared genetic origin of asthma, hay fever and eczema elucidates allergic disease biology. *Nat. Genet.* **49**, 1752–1757 (2017).
27. Arehart, C. H. *et al.* Polygenic prediction of atopic dermatitis improves with atopic training and filaggrin factors. *J. Allergy Clin. Immunol.* S0091-6749(21)00895–2 (2021) doi:10.1016/j.jaci.2021.05.034.
28. Jones, S. E. *et al.* Genome-wide association analyses of chronotype in 697,828 individuals provides insights into circadian rhythms. *Nat. Commun.* **10**, 343 (2019).
29. Jones, S. E. *et al.* Genetic studies of accelerometer-based sleep measures yield new insights into human sleep behaviour. *Nat. Commun.* **10**, 1585 (2019).
30. Lind, L. Genome-Wide Association Study of the Metabolic Syndrome in UK Biobank. *Metab. Syndr. Relat. Disord.* **17**, 505–511 (2019).
31. Dupuis, J. *et al.* New genetic loci implicated in fasting glucose homeostasis and their impact on type 2 diabetes risk. *Nat. Genet.* **42**, 105–116 (2010).
32. Teslovich, T. M. *et al.* Biological, clinical and population relevance of 95 loci for blood lipids. *Nature* **466**, 707–713 (2010).
33. Evangelou, E. *et al.* Genetic analysis of over 1 million people identifies 535 new loci associated with blood pressure traits. *Nat. Genet.* **50**, 1412–1425 (2018).
34. Roselli, C. *et al.* Multi-ethnic genome-wide association study for atrial fibrillation. *Nat. Genet.* **50**, 1225–1233 (2018).
35. Aragam, K. G. *et al.* Discovery and systematic characterization of risk variants and genes for coronary artery disease in over a million participants. *Nat. Genet.* **54**, 1803–1815 (2022).
36. Xue, A. *et al.* Genome-wide association analyses identify 143 risk variants and putative regulatory mechanisms for type 2 diabetes. *Nat. Commun.* **9**, 2941 (2018).
37. Mansour Aly, D. *et al.* Genome-wide association analyses highlight etiological differences underlying newly defined subtypes of diabetes. *Nat. Genet.* **53**, 1534–1542 (2021).
38. Selvaraj, M. S. *et al.* Genome-wide discovery for diabetes-dependent triglycerides-associated loci. *PLOS ONE* **17**, e0275934 (2022).
39. Hatoum, A. S. *et al.* Genome-wide Association Study Shows That Executive Functioning Is Influenced by GABAergic Processes and Is a Neurocognitive Genetic Correlate of Psychiatric Disorders. *Biol. Psychiatry* **93**, 59–70 (2023).
40. Colbert, S. M. C. *et al.* Novel characterization of the multivariate genetic architecture of internalizing psychopathology and alcohol use. *Am. J. Med. Genet. B Neuropsychiatr. Genet.* **186**, 353–366 (2021).

41. Saunders, G. R. B. *et al.* Genetic diversity fuels gene discovery for tobacco and alcohol use. *Nature* **612**, 720–724 (2022).
42. Karlsson Linnér, R. *et al.* Genome-wide association analyses of risk tolerance and risky behaviors in over 1 million individuals identify hundreds of loci and shared genetic influences. *Nat. Genet.* **51**, 245–257 (2019).
43. Turley, P. *et al.* Multi-trait analysis of genome-wide association summary statistics using MTAG. *Nat. Genet.* **50**, 229–237 (2018).
44. Nagel, M., Watanabe, K., Stringer, S., Posthuma, D. & van der Sluis, S. Item-level analyses reveal genetic heterogeneity in neuroticism. *Nat. Commun.* **9**, 905 (2018).
45. Nagel, M., Speed, D., van der Sluis, S. & Østergaard, S. D. Genome-wide association study of the sensitivity to environmental stress and adversity neuroticism cluster. *Acta Psychiatr. Scand.* **141**, 476–478 (2020).
46. May-Wilson, S. *et al.* Large-scale GWAS of food liking reveals genetic determinants and genetic correlations with distinct neurophysiological traits. *Nat. Commun.* **13**, 2743 (2022).
47. Persyn, E. *et al.* Genome-wide association study of MRI markers of cerebral small vessel disease in 42,310 participants. *Nat. Commun.* **11**, 2175 (2020).
48. Atkins, J. L. *et al.* A genome-wide association study of the frailty index highlights brain pathways in ageing. *Aging Cell* **20**, e13459 (2021).
49. van den Berg, S. M. *et al.* Meta-analysis of Genome-Wide Association Studies for Extraversion: Findings from the Genetics of Personality Consortium. *Behav. Genet.* **46**, 170–182 (2016).
50. de Moor, M. H. M. *et al.* Meta-analysis of genome-wide association studies for personality. *Mol. Psychiatry* **17**, 337–349 (2012).
51. Kunkle, B. W. *et al.* Genetic meta-analysis of diagnosed Alzheimer's disease identifies new risk loci and implicates Aβ, tau, immunity and lipid processing. *Nat. Genet.* **51**, 414–430 (2019).
52. Zorina-Lichtenwalter, K. *et al.* Genetic risk shared across 24 chronic pain conditions: identification and characterization with genomic structural equation modeling. *PAIN* **164**, 2239 (2023).
53. Chiou, J. *et al.* Interpreting type 1 diabetes risk with genetics and single-cell epigenomics. *Nature* **594**, 398–402 (2021).
54. Okada, Y. *et al.* Genetics of rheumatoid arthritis contributes to biology and drug discovery. *Nature* **506**, 376–381 (2014).
55. de Lange, K. M. *et al.* Genome-wide association study implicates immune activation of multiple integrin genes in inflammatory bowel disease. *Nat. Genet.* **49**, 256–261 (2017).
56. Tin, A. *et al.* Target genes, variants, tissues and transcriptional pathways influencing human serum urate levels. *Nat. Genet.* **51**, 1459–1474 (2019).
57. Hanscombe, K. B. *et al.* The genetic case for cardiorespiratory fitness as a clinical vital sign and the routine prescription of physical activity in healthcare. *Genome Med.* **13**, 180 (2021).
